# Azithromycin in Hospitalised Patients with COVID-19 (RECOVERY): a randomised, controlled, open-label, platform trial

**DOI:** 10.1101/2020.12.10.20245944

**Authors:** Peter W Horby, Alistair Roddick, Enti Spata, Natalie Staplin, Jonathan R Emberson, Guilherme Pessoa-Amorim, Leon Peto, Mark Campbell, Christopher Brightling, Ben Prudon, David Chadwick, Andrew Ustianowski, Abdul Ashish, Stacy Todd, Bryan Yates, Robert Buttery, Stephen Scott, Diego Maseda, J Kenneth Baillie, Maya H Buch, Lucy C Chappell, Jeremy N Day, Saul N Faust, Thomas Jaki, Katie Jeffery, Edmund Juszczak, Wei Shen Lim, Alan Montgomery, Andrew Mumford, Kathryn Rowan, Guy Thwaites, Marion Mafham, Richard Haynes, Martin J Landray, RECOVERY Collaborative Group

## Abstract

**Background:** Azithromycin has been proposed as a treatment for COVID-19 on the basis of its immunomodulatory actions. We evaluated the efficacy and safety of azithromycin in hospitalised patients with COVID-19.

**Methods:** In this randomised, controlled, open-label, adaptive platform trial, several possible treatments were compared with usual care in patients hospitalised with COVID-19 in the UK. Eligible and consenting patients were randomly allocated to either usual standard of care alone or usual standard of care plus azithromycin 500 mg once daily by mouth or intravenously for 10 days or until discharge (or one of the other treatment arms). Patients were twice as likely to be randomised to usual care as to any of the active treatment groups. The primary outcome was 28-day mortality. The trial is registered with ISRCTN (50189673) and clinicaltrials.gov (NCT04381936).

**Findings:** Between 7 April and 27 November 2020, 2582 patients were randomly allocated to receive azithromycin and 5182 patients to receive usual care alone. Overall, 496 (19%) patients allocated to azithromycin and 997 (19%) patients allocated to usual care died within 28 days (rate ratio 1·00; 95% confidence interval [CI] 0·90-1·12; p=0·99). Consistent results were seen in all pre-specified subgroups of patients. There was no difference in duration of hospitalisation (median 12 days vs. 13 days) or the proportion of patients discharged from hospital alive within 28 days (60% vs. 59%; rate ratio 1·03; 95% CI 0·97-1·10; p=0·29). Among those not on invasive mechanical ventilation at baseline, there was no difference in the proportion meeting the composite endpoint of invasive mechanical ventilation or death (21% vs. 22%; risk ratio 0·97; 95% CI 0·89-1·07; p=0·54).

**Interpretation:** In patients hospitalised with COVID-19, azithromycin did not provide any clinical benefit. Azithromycin use in patients hospitalised with COVID-19 should be restricted to patients where there is a clear antimicrobial indication.

**Funding:** UK Research and Innovation (Medical Research Council) and National Institute of Health Research (Grant ref: MC_PC_19056).

## INTRODUCTION

A substantial proportion of individuals infected with severe acute respiratory syndrome coronavirus 2 (SARS-CoV-2) develop a respiratory illness requiring hospital care, which can progress to critical illness with hypoxic respiratory failure requiring prolonged ventilatory support. Among COVID-19 patients admitted to UK hospitals in the first wave of the epidemic, the case fatality rate was over 26%, and in excess of 37% in patients requiring invasive mechanical ventilation.^1^

Among patients with severe COVID-19, the host immune response is thought to play a key role in driving an acute pneumonic process with diffuse alveolar damage, inflammatory infiltrates, and microvascular thrombosis.^2^ The beneficial effects of dexamethasone and other corticosteroids in patients with hypoxic lung damage suggest that other drugs that suppress or modulate the immune system may provide additional improvements in clinical outcomes.^3,4^

Macrolide antibiotics, such as azithromycin, clarithromycin and erythromycin, are widely available and their safety is well established. In addition to antibacterial properties, they are known to have immunomodulatory activity, decreasing production of pro-inflammatory cytokines and inhibiting neutrophil activation.^5-7^ They are widely used both in bacterial pneumonia due to their antimicrobial activity and in chronic inflammatory lung disease due to their immunomodulatory effects.^8,9^ In addition, azithromycin has *in vitro* antiviral activity against a range of viruses including SARS-CoV-2.^10,11^

The use of macrolides in influenza-associated pneumonia has been associated with a faster reduction in inflammatory cytokines and, in combination with naproxen, decreased mortality.^12-14^ However, randomised trials have so far failed to demonstrate convincing clinical benefit of macrolides in COVID-19.^15-17^ Here we report the preliminary results of a randomised controlled trial of azithromycin in patients hospitalised with COVID-19.

## METHODS

### Study design and participants

The Randomised Evaluation of COVID-19 therapy (RECOVERY) trial is an investigator-initiated, individually randomised, controlled, open-label, adaptive platform trial to evaluate the effects of potential treatments in patients hospitalised with COVID-19. Details of the trial design and results for other possible treatments (dexamethasone, hydroxychloroquine, and lopinavir-ritonavir) have been published previously.^3,18,19^ The trial is underway at 176 hospitals in the United Kingdom (appendix pp 2-22), supported by the National Institute for Health Research Clinical Research Network. The trial is coordinated by the Nuffield Department of Population Health at the University of Oxford (Oxford, UK), the trial sponsor. The trial is conducted in accordance with the principles of the International Conference on Harmonisation–Good Clinical Practice guidelines and approved by the UK Medicines and Healthcare products Regulatory Agency (MHRA) and the Cambridge East Research Ethics Committee (ref: 20/EE/0101). The protocol, statistical analysis plan, and additional information are available on the study website www.recoverytrial.net.

Although the azithromycin, dexamethasone, hydroxychloroquine, and lopinavir-ritonavir arms have now been stopped, the trial continues to study the effects of tocilizumab, convalescent plasma, REGEN-COV2 (a combination of two monoclonal antibodies directed against SARS-CoV-2 spike glycoprotein), aspirin, and colchicine. Other treatments may be studied in future.

Patients admitted to hospital were eligible for the study if they had clinically suspected or laboratory confirmed SARS-CoV-2 infection and no medical history that might, in the opinion of the attending clinician, put the patient at significant risk if they were to participate in the trial. Initially, recruitment was limited to patients aged at least 18 years but from 9 May 2020, the age limit was removed. Patients with known prolonged QTc interval or hypersensitivity to a macrolide antibiotic and those already receiving chloroquine or hydroxychloroquine were excluded from being randomised between azithromycin and usual care. Written informed consent was obtained from all patients, or a legal representative if they were too unwell or unable to provide consent.

### Randomisation and masking

Baseline data were collected using a web-based case report form that included demographics, level of respiratory support, major comorbidities, suitability of the study treatment for a particular patient, and treatment availability at the study site (appendix pp 26-28). Eligible and consenting patients were assigned to either usual standard of care or usual standard of care plus azithromycin or one of the other available RECOVERY treatment arms using web-based simple (unstratified) randomisation with allocation concealed until after randomisation (appendix pp 23-25). Randomisation to usual care was twice that of any of the active arms the patient was eligible for (e.g. 2:1 in favour of usual care if the patient was eligible for only one active arm, 2:1:1 if the patient was eligible for two active arms). For some patients, azithromycin was unavailable at the hospital at the time of enrolment or if a macrolide antibiotic was considered by the managing physician to be either definitely indicated or definitely contraindicated. These patients were excluded from the randomised comparison between azithromycin and usual care. Patients allocated to azithromycin were to receive azithromycin 500 mg by mouth, nasogastric tube, or intravenous injection once daily for 10 days or until discharge, if sooner. Allocated treatment was prescribed by the managing doctor. Participants and local study staff were not masked to the allocated treatment. The Steering Committee, investigators, and all others involved in the trial were masked to the outcome data during the trial.

### Procedures

A single online follow-up form was completed when participants were discharged, had died or at 28 days after randomisation, whichever occurred earliest (appendix p 29-35). Information was recorded on adherence to allocated study treatment, receipt of other COVID-19 treatments, duration of admission, receipt of respiratory or renal support, and vital status (including cause of death). In addition, routine healthcare and registry data were obtained including information on vital status (with date and cause of death), discharge from hospital, receipt of respiratory support, or renal replacement therapy.

### Outcomes

Outcomes were assessed at 28 days after randomisation, with further analyses specified at 6 months. The primary outcome was all-cause mortality. Secondary outcomes were time to discharge from hospital, and, among patients not on invasive mechanical ventilation at randomisation, invasive mechanical ventilation (including extra-corporal membrane oxygenation) or death. Prespecified subsidiary clinical outcomes were cause-specific mortality, use of haemodialysis or haemofiltration, major cardiac arrhythmia (recorded in a subset), and receipt and duration of ventilation. Among those on invasive mechanical ventilation at randomisation, a subsidiary clinical outcome of successful cessation of invasive mechanical ventilation was defined as cessation within (and survival to) 28 days. Information on suspected serious adverse reactions was collected in an expedited fashion to comply with regulatory requirements.

### Statistical Analysis

An intention-to-treat comparison was conducted between patients randomised to azithromycin and patients randomised to usual care but for whom azithromycin was both available and suitable as a treatment. For the primary outcome of 28-day mortality, the log-rank observed minus expected statistic and its variance were used to both test the null hypothesis of equal survival curves (i.e., the log-rank test) and to calculate the one-step estimate of the average mortality rate ratio. We constructed Kaplan-Meier survival curves to display cumulative mortality over the 28-day period. The 2059 patients (27%) who had not been followed for 28 days and were not known to have died by the time of the data cut for this <u>preliminary</u> analysis (30 November 2020) were either censored on 30 November 2020 or, if they had already been discharged alive, were right-censored for mortality at day 29 (that is, in the absence of any information to the contrary they were assumed to have survived 28 days). [Note: This censoring rule will not be necessary for the final report.] We used similar methods to analyse time to hospital discharge and successful cessation of invasive mechanical ventilation, with patients who died in hospital right-censored on day 29. Median time to discharge was derived from Kaplan-Meier estimates. For the pre-specified composite secondary outcome of invasive mechanical ventilation or death within 28 days (among those not receiving invasive mechanical ventilation at randomisation) and the subsidiary clinical outcomes of receipt of ventilation and use of haemodialysis or haemofiltration, the precise dates were not available and so the risk ratio was estimated instead.

Prespecified analyses of the primary outcome were performed separately in seven subgroups defined by characteristics at randomisation: age, sex, ethnicity, level of respiratory support, days since symptom onset, use of corticosteroids, and predicted 28-day mortality risk (appendix p 26). Observed effects within subgroup categories were compared using a chi-squared test for heterogeneity or trend, in accordance with the prespecified analysis plan.

Estimates of rate and risk ratios are shown with 95% confidence intervals. All p-values are 2-sided and are shown without adjustment for multiple testing. The full database is held by the study team which collected the data from study sites and performed the analyses at the Nuffield Department of Population Health, University of Oxford (Oxford, UK).

As stated in the protocol, appropriate sample sizes could not be estimated when the trial was being planned at the start of the COVID-19 pandemic (appendix p 26). As the trial progressed, the trial steering committee, whose members were unaware of the results of the trial comparisons, determined that sufficient patients should be enrolled to provide at least 90% power at a two-sided p-value of 0.01 to detect a clinically relevant proportional reduction in the primary outcome of 20% between the two groups. Consequently, on 27 November 2020, the steering committee, blinded to the results, closed recruitment to the azithromycin comparison as sufficient patients had been enrolled.

Analyses were performed using SAS version 9.4 and R version 3.4.0. The trial is registered with ISRCTN (50189673) and clinicaltrials.gov (NCT04381936).

### Role of the funding source

The funder of the study had no role in study design, data collection, data analysis, data interpretation, or writing of the report. The corresponding authors had full access to all the data in the study and had final responsibility for the decision to submit for publication.

## RESULTS

Between 7 April 2020 and 27 November 2020, 9434 (57%) of 16443 patients enrolled into the RECOVERY trial were eligible to be randomly allocated to azithromycin (that is azithromycin was available in the hospital at the time and the attending clinician was of the opinion that the patient had no known indication for or contraindication to azithromycin, figure 1; appendix p 38). 2582 patients were randomly allocated to azithromycin and 5182 were randomly allocated to usual care, with the remainder being randomly allocated to one of the other treatment arms. The mean age of study participants in this comparison was 65·3 years (SD 15·7) and the median time since symptom onset was 8 days (IQR 5 to 11 days) (table 1; appendix p 38).

**Table 1:**
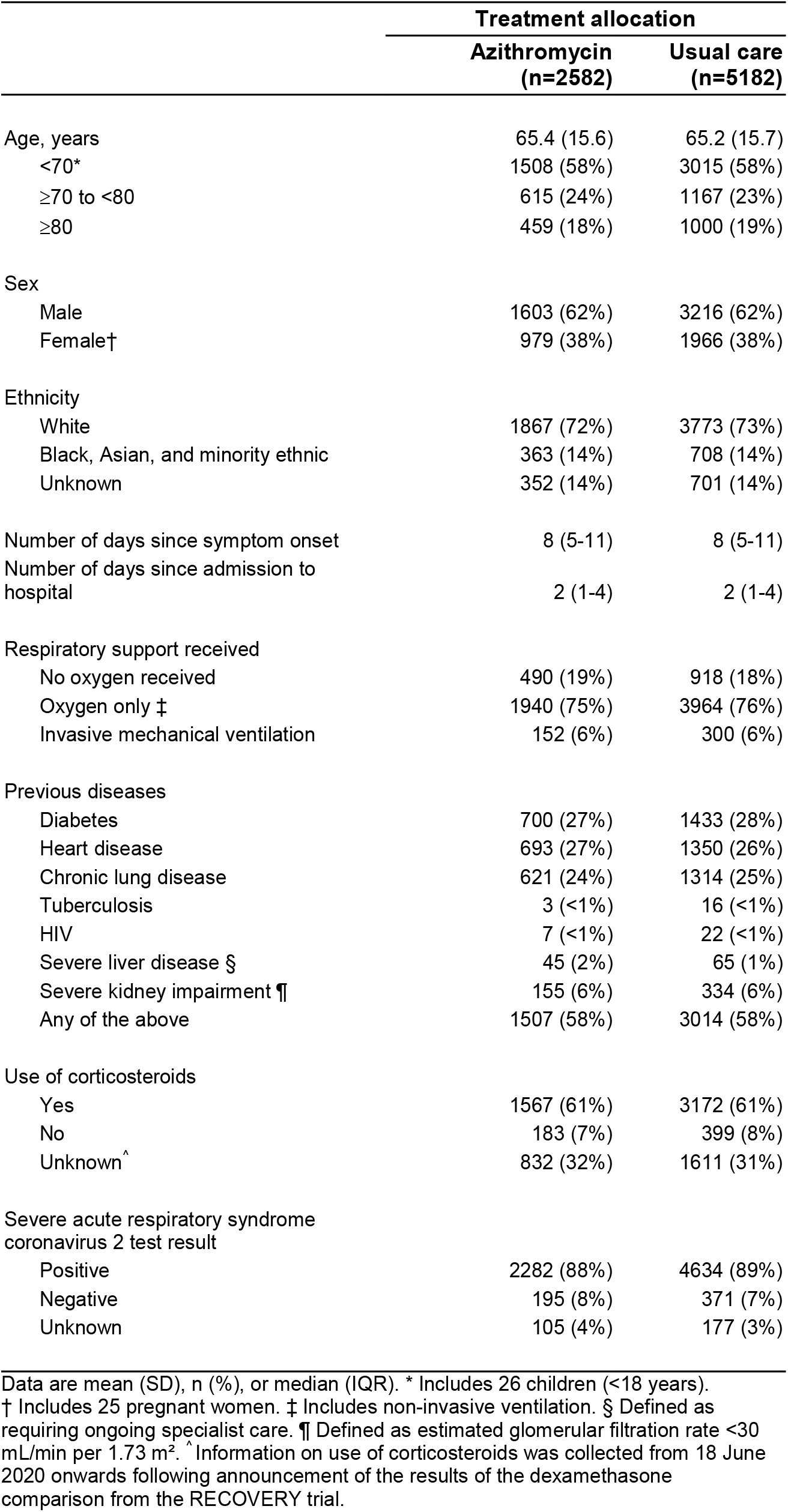
Baseline characteristics.

|  | Treatment allocation |  |
| --- | --- | --- |
|  | Azithromycin<br>(n=2582) | Usual care<br>(n=5182) |
| Age, years | 65.4 (15.6) | 65.2 (15.7) |
| <70* | 1508 (58%) | 3015 (58%) |
| ≥70 to <80 | 615 (24%) | 1167 (23%) |
| ≥80 | 459 (18%) | 1000 (19%) |
| Sex |  |  |
| Male | 1603 (62%) | 3216 (62%) |
| Female† | 979 (38%) | 1966 (38%) |
| Ethnicity |  |  |
| White | 1867 (72%) | 3773 (73%) |
| Black, Asian, and minority ethnic | 363 (14%) | 708 (14%) |
| Unknown | 352 (14%) | 701 (14%) |
| Number of days since symptom onset | 8 (5-11) | 8 (5-11) |
| Number of days since admission to hospital | 2 (1-4) | 2 (1-4) |
| Respiratory support received |  |  |
| No oxygen received | 490 (19%) | 918 (18%) |
| Oxygen only ‡ | 1940 (75%) | 3964 (76%) |
| Invasive mechanical ventilation | 152 (6%) | 300 (6%) |
| Previous diseases |  |  |
| Diabetes | 700 (27%) | 1433 (28%) |
| Heart disease | 693 (27%) | 1350 (26%) |
| Chronic lung disease | 621 (24%) | 1314 (25%) |
| Tuberculosis | 3 (<1%) | 16 (<1%) |
| HIV | 7 (<1%) | 22 (<1%) |
| Severe liver disease § | 45 (2%) | 65 (1%) |
| Severe kidney impairment ¶ | 155 (6%) | 334 (6%) |
| Any of the above | 1507 (58%) | 3014 (58%) |
| Use of corticosteroids |  |  |
| Yes | 1567 (61%) | 3172 (61%) |
| No | 183 (7%) | 399 (8%) |
| Unknown^ | 832 (32%) | 1611 (31%) |
| Severe acute respiratory syndrome coronavirus 2 test result |  |  |
| Positive | 2282 (88%) | 4634 (89%) |
| Negative | 195 (8%) | 371 (7%) |
| Unknown | 105 (4%) | 177 (3%) |

Data are mean (SD), n (%), or median (IQR). \* Includes 26 children (<18 years).
† Includes 25 pregnant women. ‡ Includes non-invasive ventilation. § Defined as requiring ongoing specialist care. ¶ Defined as estimated glomerular filtration rate <30 mL/min per 1.73 m<sup>2</sup>. ^ Information on use of corticosteroids was collected from 18 June 2020 onwards following announcement of the results of the dexamethasone comparison from the RECOVERY trial.

**Figure 1:**
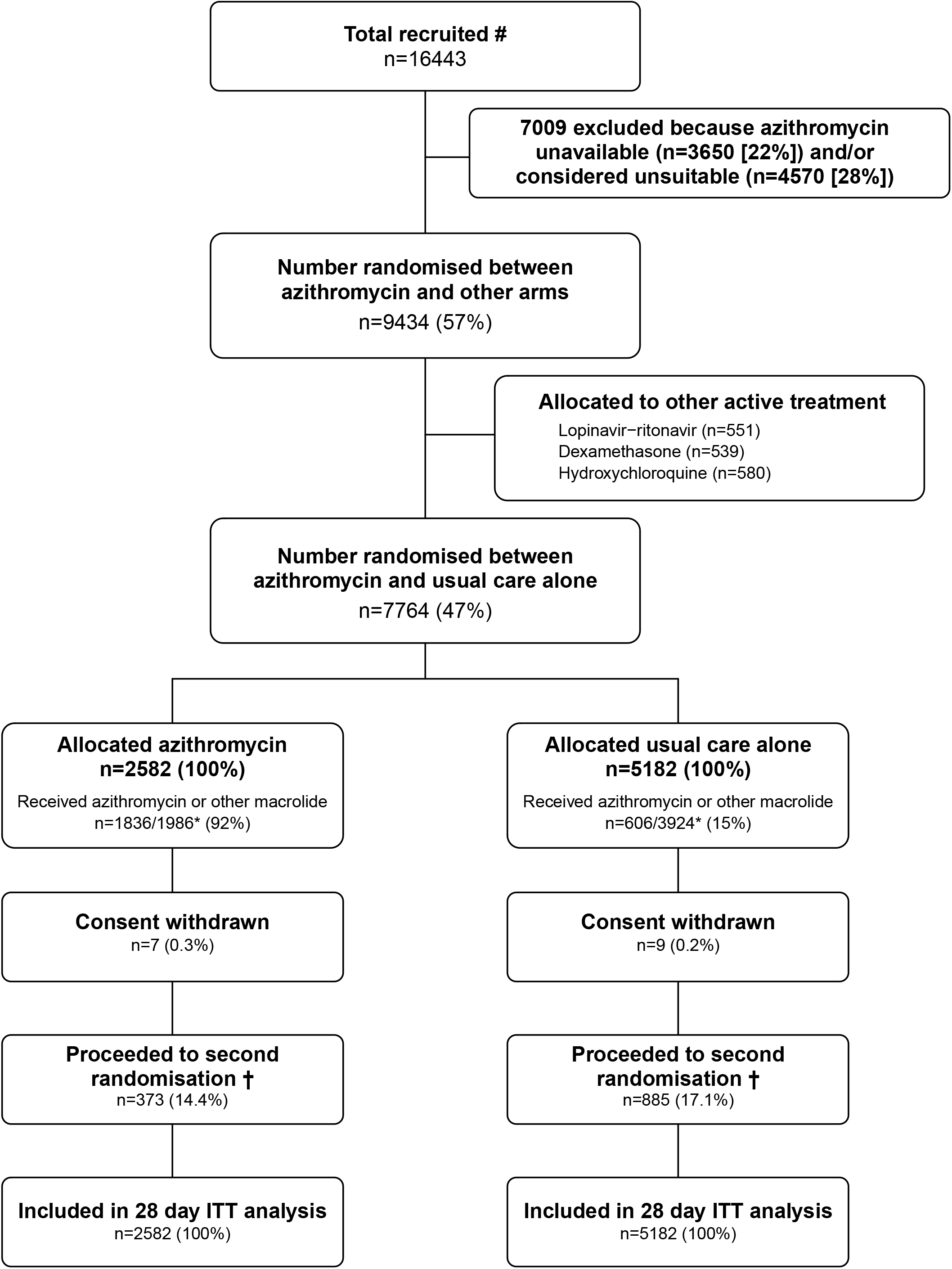
Trial profile. ITT=intention to treat. # Number recruited overall during period that adult participants could be recruited into azithromycin comparison. * 1986/2582 (77%) and 3924/5182 (76%) patients have a completed follow-up form at time of analysis. 3994 patients were additionally randomised to convalescent plasma vs REGN-COV2 vs usual care (1320 [51%] patients allocated to azithromycin vs 2674 [52%] patients allocated usual care) and 975 patients were additionally randomised to aspirin vs usual care (323 [13%] patients allocated to azithromycin vs 652 [13%] patients allocated usual care. † Includes 197/2582 (7.6%) patients in the azithromycin arm and 446/5182 (8.6%) patients in the usual care arm allocated to tocilizumab.

Among the 5910 (76%) patients for whom a follow-up form has been completed to date, 1760 (89%) allocated to azithromycin vs. 55 (1%) allocated to usual care received at least one dose, and 1836 (92%) vs. 606 (15%) received any macrolide antibiotic (figure 1; appendix p 39). The median duration of treatment with azithromycin was 6 days (IQR 3-9 days). Use of other treatments for COVID-19 was similar among patients allocated azithromycin and among those allocated usual care, with nearly one half receiving a corticosteroid, about one-fifth receiving remdesivir, and one-fifth receiving convalescent plasma.

We observed no significant difference in the proportion of patients who met the primary outcome of 28-day mortality between the two randomised groups (496 [19%] patients in the azithromycin group vs. 997 (19%) patients in the usual care group; rate ratio 1·00; 95% confidence interval [CI], 0·90 to 1·12; p=0·99; figure 2). We observed similar results across all pre-specified subgroups (figure 3). In an exploratory analysis restricted to the 6916 (89%) patients with a positive SARS-CoV-2 test result, the result was similar (rate ratio 0·99, 95% CI 0·88 to 1·10; p=0·81).

**Figure 2:**
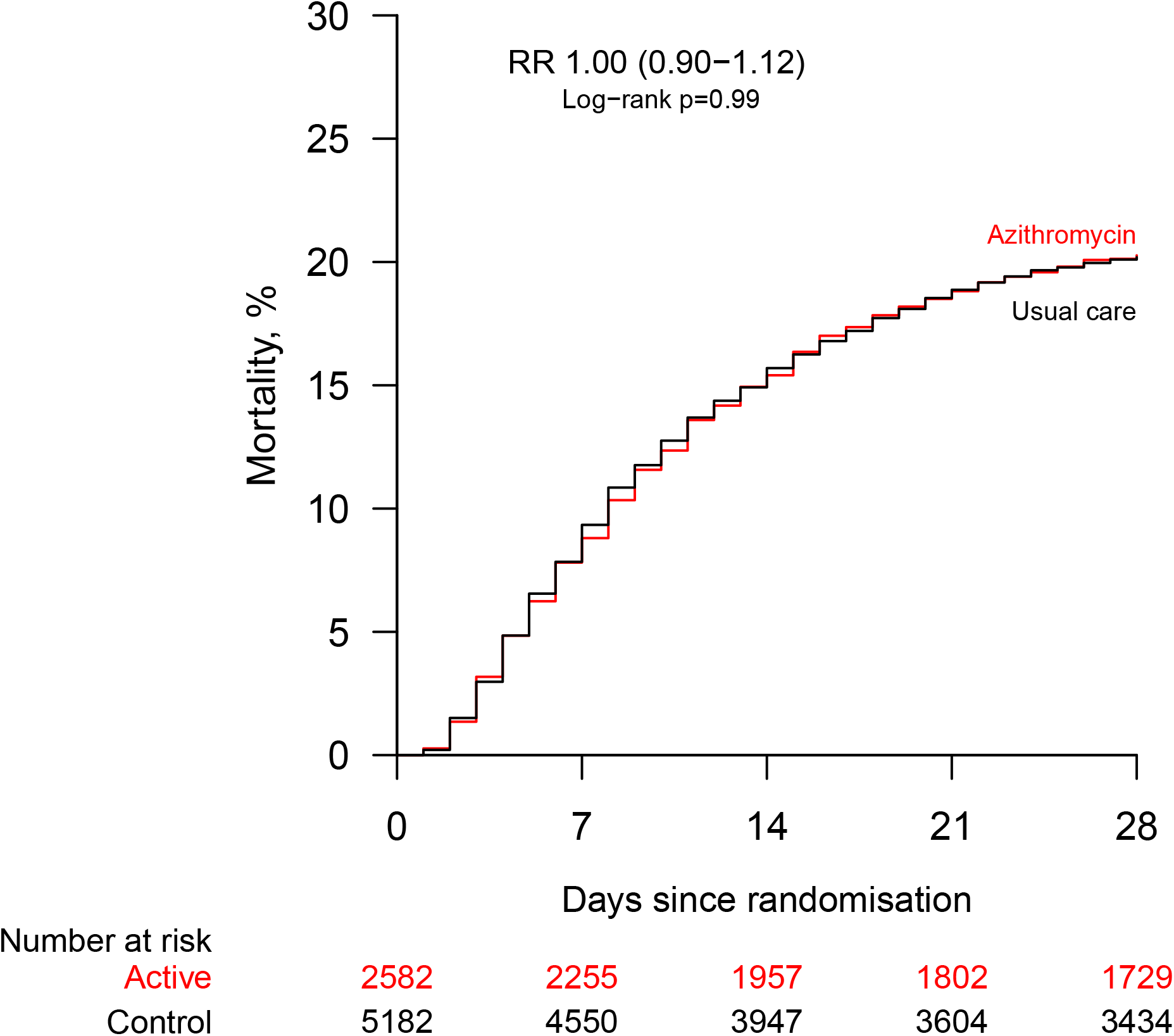
Effect of allocation to azithromycin on 28-day mortality.

**Figure 3:**
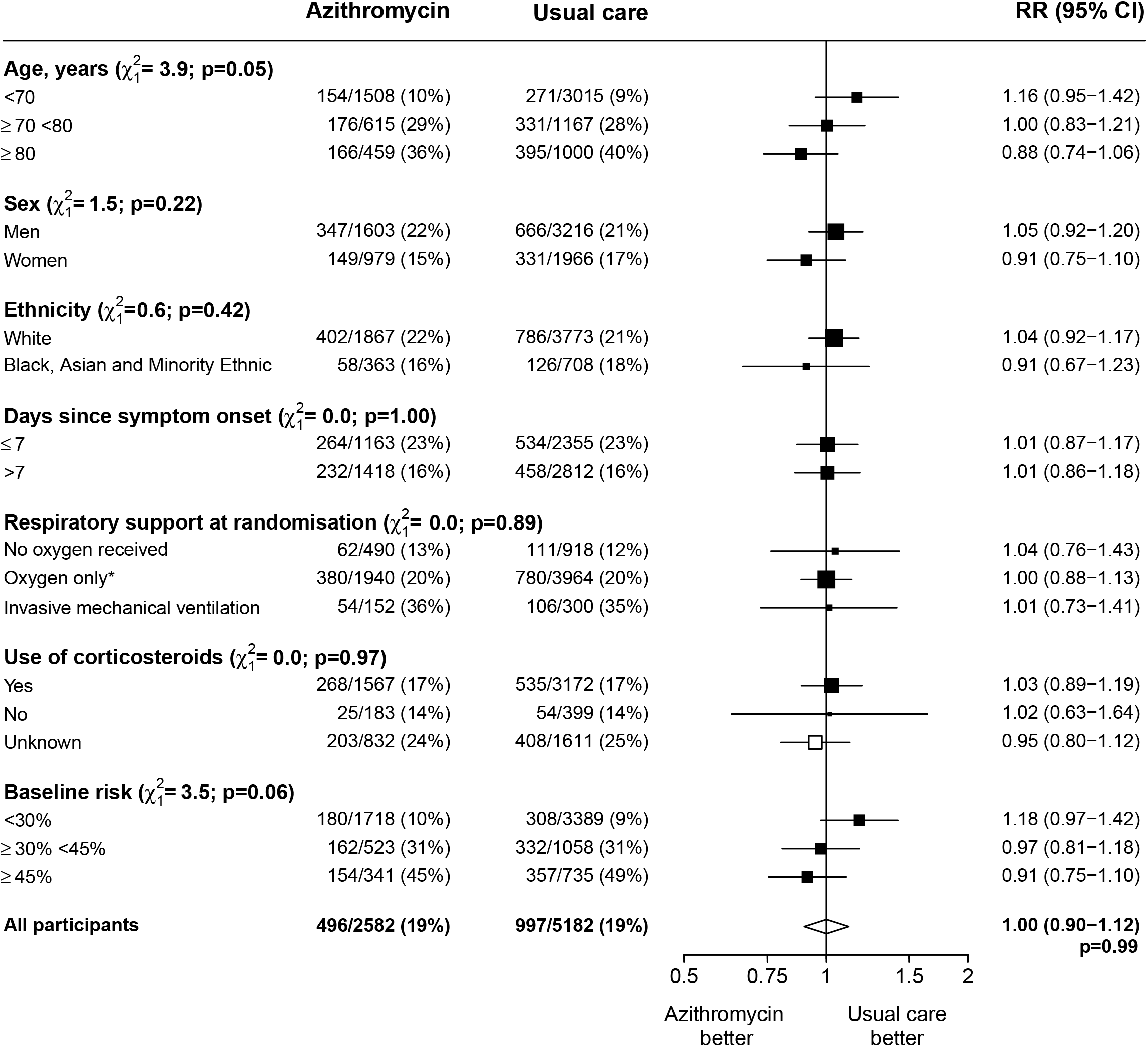
Effect of allocation to azithromycin on 28-day mortality by baseline characteristics. Subgroup-specific rate ratio estimates are represented by squares (with areas of the squares proportional to the amount of statistical information) and the lines through them correspond to the 95% CIs. The ethnicity and days since onset subgroups exclude those with missing data, but these patients are included in the overall summary diamond. * Includes patients receiving non-invasive ventilation. Information on use of corticosteroids was collected from 18 June 2020 onwards following announcement of the results of the dexamethasone comparison from the RECOVERY trial.

Allocation to azithromycin was associated with a similar time until discharge from hospital alive as usual care (median 12 days vs. 13 days) and a similar probability of discharge alive within 28 days (60% vs. 59%, rate ratio 1·03, 95% CI 0·97 to 1·10, p=0·29) (Table 2). Among those not on invasive mechanical ventilation at baseline, the number of patients progressing to the pre-specified composite secondary outcome of invasive mechanical ventilation or death among those allocated to azithromycin was similar to that among those allocated to usual care (21% vs. 22%, risk ratio 0·97, 95% CI 0·89 to 1·07, p=0·54). Allowing for multiple testing in interpretation of the results, there was no evidence that the effect of allocation to azithromycin vs. usual care on time until discharge from hospital alive or on invasive mechanical ventilation or death differed between pre-specified subgroups of patients (appendix p 43-44).

**Table 2:** Effect of allocation to azithromycin on key study outcomes.

|  | Treatment allocation |  | RR (95% CI) | p-value |
| --- | --- | --- | --- | --- |
|  | Azithromycin<br>(n=2582) | Usual care<br>(n=5182) |  |  |
| Primary outcome: |  |  |  |  |
| 28-day mortality | 496 (19%) | 997 (19%) | 1.00 (0.90-1.12) | 0.99 |
| Secondary outcomes: |  |  |  |  |
| Median time to being discharged alive, days | 12 | 13 |  |  |
| Discharged from hospital within 28 days | 1554 (60%) | 3066 (59%) | 1.03 (0.97-1.10) | 0.29 |
| Receipt of invasive mechanical ventilation or death* | 517/2430 (21%) | 1069/4882 (22%) | 0.97 (0.89-1.07) | 0.54 |
| Invasive mechanical ventilation | 154/2430 (6%) | 325/4882 (7%) | 0.95 (0.79-1.15) | 0.60 |
| Death | 442/2430 (18%) | 891/4882 (18%) | 1.00 (0.90-1.10) | 0.95 |
| Subsidiary clinical outcomes |  |  |  |  |
| Receipt of ventilation † | 27/490 (6%) | 50/918 (5%) | 1.01 (0.64-1.59) | 0.96 |
| Non-invasive ventilation | 24/490 (5%) | 43/918 (5%) | 1.05 (0.64-1.70) | 0.86 |
| Invasive mechanical ventilation | 8/490 (2%) | 11/918 (1%) | 1.36 (0.55-3.37) | 0.50 |
| Successful cessation of invasive mechanical ventilation ‡ | 42/152 (28%) | 95/300 (32%) | 0.84 (0.59-1.20) | 0.33 |
| Use of haemodialysis or haemofiltration § | 79/2548 (3%) | 158/5125 (3%) | 1.01 (0.77-1.31) | 0.97 |

Data are n (%) or n/N (%), unless otherwise indicated. RR=rate ratio for the outcomes of 28-day mortality, hospital discharge and successful cessation of invasive mechanical ventilation, and risk ratio for other outcomes. \* Analyses exclude those on invasive mechanical ventilation at randomisation. † Analyses exclude those on any form of ventilation at randomisation. ‡ Analyses restricted to those on invasive mechanical ventilation at randomisation. § Analyses exclude those on haemodialysis or haemofiltration at randomisation.

We found no significant differences in the prespecified subsidiary clinical outcomes of cause-specific mortality (appendix p 40), use of ventilation, successful cessation of invasive mechanical ventilation, or need for renal dialysis or haemofiltration (Table 2). We observed no significant differences in the frequency of new cardiac arrhythmias (appendix p 41). There was one report of a serious adverse reaction believed related to azithromycin: a case of pseudomembranous colitis from which the patient recovered with standard treatment.

## DISCUSSION

The results of this large randomised trial show that azithromycin is not an effective treatment for patients hospitalised with COVID-19. Allocation to azithromycin was not associated with reductions in mortality, duration of hospitalisation or the risk of being ventilated or dying for those not on ventilation at baseline. These results were consistent across the prespecified subgroups of age, sex, ethnicity, duration of symptoms prior to randomisation, level of respiratory support at randomisation, use of corticosteroids, and baseline predicted risk of death at randomisation.

Azithromycin was proposed as a treatment for COVID-19 based on its immunomodulatory activity.^7^ Although no major organization or professional society has recommended the routine use of azithromycin in COVID-19 unless there is evidence of bacterial super-infection, it has nevertheless been used widely in COVID-19 patients, particularly in combination with hydroxychloroquine.^20-22^ Macrolides have long been suggested as potential therapies for inflammatory viral pneumonias but this has been based on in vitro, animal and observational data, with very little clinical trial evidence of benefit.^11-13^ The benefit of dexamethasone in COVID-19 patients requiring respiratory support suggests that inflammation has a causal role in mortality.^3^ Noting that the results were consistent regardless of whether patients were also being treated with a corticosteroid or not, we conclude that the immunomodulatory properties of azithromycin are either insufficient or off-target in COVID-19.

Macrolides are commonly used to treat bacterial infections of the lower respiratory tract because of their good activity against Gram positive bacteria and atypical pathogens such as *Mycoplasma pneumoniae* and *Legionella* species, and their excellent tissue penetration. More than 75% of hospitalised COVID-19 patients are prescribed antibiotics and the widespread clinical use of macrolides in COVID-19 is likely to be driven largely by concerns of bacterial superinfection rather than purported immunomodulatory activity.^23^ It is therefore important to highlight that in patients with moderate or severe COVID-19, who might be expected to experience some burden of secondary bacterial lung infection, there was no observed clinical benefit of azithromycin use. This lack of effect may either reflect the relatively low rate of secondary bacterial infection in COVID-19 or the widespread use of other antibiotics such as β-lactam antibiotics, which may have abrogated any anti-bacterial benefit of allocation to azithromycin in this trial.^24,25^ Our results show that azithromycin confers no clinical benefit in hospitalised COVID-19 patients, whether that be anti-inflammatory or antimicrobial. Although we detected no harm to individual patients treated with azithromycin, there is a risk of harm at a societal level from widespread use of antimicrobial agents. Azithromycin is classified within the WHO Watch Group of Antibiotics: antibiotics that have higher resistance potential and should be prioritized as key targets of antimicrobial stewardship programs. In light of the new evidence from the RECOVERY trial, the widespread use in COVID-19 patients of macrolides in particular and antibiotics in general must be questioned.^26^

Our trial has some limitations: Detailed information on laboratory markers of inflammatory status, co-existent bacterial infection, or use of non-macrolide antibiotics was not collected, nor was information on radiological or physiological outcomes. This initial report is based on complete follow-up for the primary outcome in 73% of patients (and partial follow-up for the remaining 27%). However, collection of outcome information both through case report forms and linkage to routine NHS records is ongoing and, based on previous reports from this trial, will deliver complete follow-up information for over 99% of patients by early January 2021. However, additional follow-up is unlikely to change the conclusion that azithromycin has no meaningful benefit for hospitalised patients with COVID-19.

Three other randomised controlled trials have assessed the efficacy of azithromycin for the treatment of COVID-19 in hospitalised patients, all of which additionally treated patients with hydroxychloroquine.^15-17^ The COALITION I and COALITION II trials found that allocation of hospitalized patients with COVID-19 to azithromycin and hydroxychloroquine, was not associated with any improvement in mortality, duration of hospital stay, or clinical status as assessed using an ordinal outcome scale.^15,16^ A small trial in Iran that randomised patients to hydroxychloroquine and lopinavir-ritonavir with or without azithromycin also found no significant difference in mortality or intensive care unit admission, but suggested a reduction in duration of hospital stay.^17^ The total number of patients in all three prior trials combined was 1223, with 130 deaths. The RECOVERY trial, with 7764 participants and 1483 deaths in this assessment of azithromycin, is far better powered to detect modest treatment benefits; none were observed.

At the time of writing, 24 trials evaluating the use of macrolides in COVID-19 patients were registered in the WHO International Clinical Trials Registry Platform, of which two (COALITION I and COALITION II, described above) have published results. Of the remaining 22, 16 are studying macrolides in inpatients either alone or in combination with other putative treatments, whilst 6 are studying non-hospitalised patients with suspected or confirmed COVID-19.

Whilst our findings do not address the use of macrolides for the treatment of non-hospitalised COVID-19 patients with early, mild disease, the results do show that azithromycin is not an effective treatment for hospitalised COVID-19 patients.

## Supporting information

Protocol and Statistical Analysis Plan

## Data Availability

The protocol, consent form, statistical analysis plan, definition & derivation of clinical characteristics & outcomes, training materials, regulatory documents, and other relevant study materials are available online at www.recoverytrial.net. As described in the protocol, the trial Steering Committee will facilitate the use of the study data and approval will not be unreasonably withheld. Deidentified participant data will be made available to bona fide researchers registered with an appropriate institution within 3 months of publication. However, the Steering Committee will need to be satisfied that any proposed publication is of high quality, honours the commitments made to the study participants in the consent documentation and ethical approvals, and is compliant with relevant legal and regulatory requirements (e.g. relating to data protection and privacy). The Steering Committee will have the right to review and comment on any draft manuscripts prior to publication.

https://www.ndph.ox.ac.uk/data-access

https://www.recoverytrial.net

## NOTE

Enrolment to the azithromycin arm of the RECOVERY trial was closed on 27 November 2020. Here we report the preliminary findings based on a data cut on 30 November 2020. Final results will be made available after the last patient has completed the 28-day follow-up period for the primary outcome on 25 December 2020. As with previous reports, we anticipate >99% follow-up of all patients due to the linkage with routine NHS data. With 1483 deaths among a total of 7764 patients included in the current report, the findings are unlikely to change in any material way.

## Contributors

This manuscript was initially drafted by PWH and MJL, further developed by the Writing Committee, and approved by all members of the trial steering committee. PWH and MJL vouch for the data and analyses, and for the fidelity of this report to the study protocol and data analysis plan. PWH, MM, JKB, LCC, SNF, TJ, KJ, WSL, AM, KR, EJ, RH, and MJL designed the trial and study protocol. MM, AR, G P-A, CB, BP, DC, AU, AA, ST, BY, RB, SS, DM, RH, the Data Linkage team at the RECOVERY Coordinating Centre, and the Health Records and Local Clinical Centre staff listed in the appendix collected the data. ES, NS, and JRE did the statistical analysis. All authors contributed to data interpretation and critical review and revision of the manuscript. PWH and JL had access to the study data and had final responsibility for the decision to submit for publication.

## Writing Committee (on behalf of the RECOVERY Collaborative Group)

Professor Peter W Horby PhD FRCP,^a,^* Alistair Roddick MBBS,^b,c,*^ Enti Spata,^b,d,*^ Natalie Staplin PhD,^b,d^ Professor Jonathan R Emberson PhD,^b,d^ Guilherme Pessoa-Amorim MD,^b,e^ Leon Peto PhD,^a,c^ Mark Campbell FRCPath,^b,c^ Professor Christopher Brightling FRCP,^f^ Benjamin Prudon FRCP,^f^ David Chadwick PhD,^h^ Andrew Ustianowski PhD,^i^ Abdul Ashish MD,j Stacy Todd PhD,^k^ Bryan Yates MRCP,^l^ Robert Buttery PhD,^m^ Stephen Scott PhD,^n^ Diego Maseda MD,^o^ J Kenneth Baillie MD PhD,^p^ Professor Maya H Buch PhD FRCP,^q^ Professor Lucy C Chappell PhD,^r^ Professor Jeremy Day PhD FRCP,^a,s^ Professor Saul N Faust PhD FRCPCH,^t^ Professor Thomas Jaki PhD,^u,v^ Katie Jeffery PhD FRCP FRCPath,^c^ Professor Edmund Juszczak MSc,^w^ Professor Wei Shen Lim FRCP,^w,x^ Professor Alan Montgomery PhD,^w^ Professor Andrew Mumford PhD,^y^ Kathryn Rowan PhD,^z^ Professor Guy Thwaites PhD FRCP,^a,s^ Marion Mafham MD,^b,†^ Professor Richard Haynes DM,^b,d,†^ Professor Martin J Landray PhD FRCP.^b,d,e,†^

^a^ Nuffield Department of Medicine, University of Oxford, Oxford, United Kingdom.

^b^ Nuffield Department of Population Health, University of Oxford, Oxford, United Kingdom

^c^ Oxford University Hospitals NHS Foundation Trust, Oxford, United Kingdom

^d^ MRC Population Health Research Unit, University of Oxford, Oxford, United Kingdom

^e^ NIHR Oxford Biomedical Research Centre, Oxford University Hospitals NHS Foundation Trust, Oxford, United Kingdom

^f^ Institute for Lung Health, Leicester NIHR Biomedical Research Centre, University of Leicester, Leicester, United Kingdom

^g^ Department of Respiratory Medicine, North Tees & Hartlepool NHS Foundation Trust, Stockton-on-Tees, United Kingdom

^h^ Centre for Clinical Infection, James Cook University Hospital, Middlesbrough, United Kingdom

^i^ North Manchester General Hospital, Pennine Acute Hospitals NHS Trust, Bury, United Kingdom

^j^ Wrightington Wigan and Leigh NHS Foundation Trust, Wigan, United Kingdom

^k^ Liverpool University Hospitals NHS Foundation Trust, Liverpool, United Kingdom

^l^ Northumbria Healthcare NHS Foundation Trust, North Tyneside, United Kingdom

^m^ North West Anglia NHS Foundation Trust, Peterborough, United Kingdom

^n^ The Countess of Chester Hospital NHS Foundation Trust, Chester, United Kingdom

^o^ Mid Cheshire Hospitals NHS Foundation Trust, Crewe, United Kingdom

^p^ Roslin Institute, University of Edinburgh, Edinburgh, United Kingdom

^q^ Centre for Musculoskeletal Research, University of Manchester, Manchester, and NIHR Manchester Biomedical Research Centre, United Kingdom.

^r^ School of Life Course Sciences, King’s College London, London, United Kingdom

^s^ Oxford University Clinical Research Unit, Ho Chi Minh City, Viet Nam

^t^ NIHR Southampton Clinical Research Facility and Biomedical Research Centre, University Hospital Southampton NHS Foundation Trust and University of Southampton, Southampton, United Kingdom

^u^ Department of Mathematics and Statistics, Lancaster University, Lancaster, United Kingdom

^v^ MRC Biostatistics Unit, University of Cambridge, Cambridge, United Kingdom

^w^ School of Medicine, University of Nottingham, Nottingham, United Kingdom

^x^ Respiratory Medicine Department, Nottingham University Hospitals NHS Trust, Nottingham, United Kingdom

^y^ School of Cellular and Molecular Medicine, University of Bristol, Bristol, United kingdom

^z^ Intensive Care National Audit & Research Centre, London, United Kingdom

*,^†^ equal contribution

## Data Monitoring Committee

Peter Sandercock, Janet Darbyshire, David DeMets, Robert Fowler, David Lalloo, Ian Roberts, Janet Wittes.

## Declaration of interests

The authors have no conflict of interest or financial relationships relevant to the submitted work to disclose. No form of payment was given to anyone to produce the manuscript. All authors have completed and submitted the ICMJE Form for Disclosure of Potential Conflicts of Interest. The Nuffield Department of Population Health at the University of Oxford has a staff policy of not accepting honoraria or consultancy fees directly or indirectly from industry (see https://www.ndph.ox.ac.uk/files/about/ndph-independence-of-research-policy-jun-20.pdf).

## Data sharing

The protocol, consent form, statistical analysis plan, definition & derivation of clinical characteristics & outcomes, training materials, regulatory documents, and other relevant study materials are available online at www.recoverytrial.net. As described in the protocol, the trial Steering Committee will facilitate the use of the study data and approval will not be unreasonably withheld. Deidentified participant data will be made available to bona fide researchers registered with an appropriate institution within 3 months of publication. However, the Steering Committee will need to be satisfied that any proposed publication is of high quality, honours the commitments made to the study participants in the consent documentation and ethical approvals, and is compliant with relevant legal and regulatory requirements (e.g. relating to data protection and privacy). The Steering Committee will have the right to review and comment on any draft manuscripts prior to publication. Data will be made available in line with the policy and procedures described at: https://www.ndph.ox.ac.uk/data-access. Those wishing to request access should complete the form at https://www.ndph.ox.ac.uk/files/about/data_access_enquiry_form_13_6_2019.docx and e-mailed to

## Acknowledgements

Above all, we would like to thank the thousands of patients who participated in this study. We would also like to thank the many doctors, nurses, pharmacists, other allied health professionals, and research administrators at 176 NHS hospital organisations across the whole of the UK, supported by staff at the National Institute of Health Research (NIHR) Clinical Research Network, NHS DigiTrials, Public Health England, Department of Health & Social Care, the Intensive Care National Audit & Research Centre, Public Health Scotland, National Records Service of Scotland, the Secure Anonymised Information Linkage (SAIL) at University of Swansea, and the NHS in England, Scotland, Wales and Northern Ireland.

The RECOVERY trial is supported by a grant to the University of Oxford from UK Research and Innovation/NIHR (Grant reference: MC_PC_19056) and by core funding provided by NIHR Oxford Biomedical Research Centre, Wellcome,the Bill and Melinda Gates Foundation,the Department for International Development,Health Data Research UK, the Medical Research Council Population Health Research Unit, the NIHR Health Protection Unit in Emerging and Zoonotic Infections, and NIHR Clinical Trials Unit Support Funding. TJ is supported by a grant from UK Medical Research Council (MC_UU_0002/14) and an NIHR Senior Research Fellowship (NIHR-SRF-2015-08-001). WSL is supported by core funding provided by NIHR Nottingham Biomedical Research Centre. Abbvie contributed some supplies of lopinavir-ritonavir for use in this study. Tocilizumab was provided free of charge for this study by Roche Products Limited. REGN-COV2 was provided free of charge for this study by Regeneron.

The views expressed in this publication are those of the authors and not necessarily those of the NHS, the National Institute for Health Research or the Department of Health and Social Care.

## Notes

### Competing Interest Statement

The authors have declared no competing interest.

### Clinical Trial

The trial is registered with ISRCTN (50189673) and clinicaltrials.gov (NCT04381936

### Clinical Protocols

https://www.recoverytrial.net

### Funding Statement

The RECOVERY trial is supported by a grant to the University of Oxford from UK Research and Innovation (Medical Research Council)/National Institute for Health Research(Grant reference: MC_PC_19056)  and by core funding provided by NIHR Oxford Biomedical Research Centre, Wellcome, the Bill and Melinda Gates Foundation, the Department for International Development, Health Data Research UK, the Medical Research Council Population Health Research Unit, the NIHR Health Protection Unit in Emerging and Zoonotic Infections, and NIHR Clinical Trials Unit Support Funding. TJ is supported by a grant from UK Medical Research Council (MC_UU_0002/14) and an NIHR Senior Research Fellowship (NIHR-SRF-2015-08-001). WSL is supported by core funding provided by NIHR Nottingham Biomedical Research Centre. Abbvie contributed some supplies of lopinavir-ritonavir for use in this study. Tocilizumab was provided free of charge for this study by Roche Products Limited. REGN-COV2 was provided free of charge for this study by Regeneron.
The views expressed in this publication are those of the authors and not necessarily those of the NHS, the National Institute for Health Research or the Department of Health and Social Care.

### Author Declarations

The trial is conducted in accordance with the principles of the International Conference on Harmonisation-Good Clinical Practice guidelines and approved by the UK Medicines and Healthcare products Regulatory Agency (MHRA) and the Cambridge East Research Ethics Committee (ref: 20/EE/0101).

