## Supplementary Appendix for "Azithromycin in Hospitalised Patients with COVID-19 (RECOVERY): a randomised, controlled, open-label, platform trial"

#### RECOVERY Collaborative Group

##### Contents

**Details of the RECOVERY Collaborative Group ...... 2**
**Supplementary Methods ...... 23**
Study organization 23
Protocol changes 23
Supplementary statistical methods 26
Webtable 1: Baseline characteristics of patients considered unsuitable for randomisation to azithromycin compared with those randomised to azithromycin versus usual care 38
Webtable 2: Treatments given, by randomised allocation 39
Webtable 3: Effect of allocation to azithromycin on cause-specific 28-day mortality 40
Webtable 4: Effect of allocation to azithromycin on cardiac arrhythmia 41
**Supplementary Figures ...... 42**
Webfigure 1: Effect of allocation to azithromycin on hospital discharge by baseline characteristics 43
Webfigure 2: Effect of allocation to azithromycin on invasive mechanical ventilation or death in those not on invasive mechanical ventilation at randomisation, by baseline characteristics 44

### Details of the RECOVERY Collaborative Group

#### Writing Committee

PW Horby*, A Roddick*, E Spata*, N Staplin, J Emberson, G Pessoa-Amorim, C Brightling, B Prudon, D Chadwick, A Ustianowski, A Ashish, S Todd, B Yates, R Buttery, S Scott, D Maseda, JK Baillie, M Buch, L Chappell, J Day, SN Faust, T Jaki, K Jeffery, E Juszczak, WS Lim, A Montgomery, A Mumford, K Rowan, G Thwaites, M Mafham†, R Haynes†, MJ Landray†

*,† equal contribution

#### Steering Committee

*Co-Chief Investigators* PW Horby, MJ Landray, *Members* JK Baillie, M Buch, L Chappell, J Day, SN Faust, R Haynes, T Jaki, K Jeffery, E Juszczak, WS Lim, M Mafham, A Montgomery, A Mumford, K Rowan, G Thwaites.

#### Data Monitoring Committee

P Sandercock (chair), J Darbyshire, D DeMets, R Fowler, D Lalloo, I Roberts, J Wittes *Non-voting statisticians* J Emberson, N Staplin.

#### RECOVERY Trial Central Coordinating Office

*Co-Chief Investigators* P Horby, MJ Landray; *Clinical Trial Unit Lead* R Haynes; *Trial management* L Fletcher (coordinator), J Barton, A Basoglu, R Brown, W Brudlo, S Howard, G McChlery, K Taylor; *Programming and validation* G Cui, B Goodenough, A King, M Lay, D Murray, W Stevens, K Wallendszus, R Welsh; *Data linkage* C Crichton, J Davies, R Goldacre, C Harper, F Knight, J Latham-Mollart, M Mafham, M Nunn, H Salih, J Welch; *Clinical support* M Campbell, G Pessoa-Amorim, L Peto, A Roddick; *Quality assurance* C Knott, J Wiles; *Statistics* JL Bell, J Emberson, E Juszczak, L Linsell, E Spata, N Staplin; *Communications* G Bagley, S Cameron, S Chamberlain, B Farrell, H Freeman, A Kennedy, A Whitehouse, S Wilkinson, C Wood; *Administrative support* L Howie, M Lunn, P Rodgers

#### National Institute for Health Research Clinical Research Network

*Coordinating Centre* A Barnard, J Beety, C Birch, M Brend, E Chambers, L Chappell, S Crawshaw, C Drake, H Duckles-Leech, J Graham, T Harman, H Harper, S Lock, K Lomme, N McMillan, I Nickson, U Ohia, E OKell, V Poustie**,** S Sam, P Sharratt, J Sheffield, H Slade, W Van't Hoff, S Walker, J Williamson; *Urgent Public Health Clinical Links* A De Soyza, P Dimitri, SN Faust, N Lemoine, J Minton; *East Midlands* K Gilmour, K Pearson *Eastern* C Armah, D Campbell, H Cate, A Priest, E Thomas, R Usher; *North East & North Cumbria* G Johnson, M Logan, S Pratt, A Price, K Shirley, E Walton, P Williams, F Yelnoorkar; *Kent, Surrey & Sussex* J Hanson, H Membrey, L Gill, A Oliver; *North West London* S Das, S Murphy, M Sutu; *Greater Manchester* J Collins, H Monaghan, A Unsworth, S Beddows; *North West Coast* K Barker-Williams, S Dowling, K Gibbons, K Pine; *North Thames* A Asghar, P Aubrey, D Beaumont-Jewell, K Donaldson, T Skinner; *South London* J Luo, N Mguni, N Muzengi, R Pleass, E Wayman; *South West Peninsula* A Coe, J Hicks, M Hough, C Levett, A Potter, J Taylor; *Thames Valley and South Midlands* M Dolman, L Gerdes, C Hall, T Lockett, D Porter *Wessex* J Bartholomew, L Dowden, C Rook, J Walters; *West of England* E Denton, H Tinkler; *Yorkshire & Humber* A Alexander, H Campbell, K Chapman, A Hall, A Rodgers; *West Midlands* P Boyle, M Brookes, C Callens, H Duffy, C Green, K Hampshire, S Harrison, J Kirk, M Naz, L Porter, P Ryan, J Shenton, J Warmington; *Devolved nations* M Amezaga, P Dicks, J Goodwin, H Hodgson, S Jackson, M Odam, D Williamson.

##### Paediatric working group

SN Faust (coordinator), A Bamford, S Bandi, J Bernatoniene, K Cathie, P Dmitri, S Drysdale, M Emonts, J Evans, A Finn, P Fleming, J Furness, C Gale, R Haynes, CE Jones, E Juszczak, D Jyotish, D Kelly, C Murray, N Pathan, L Pollock, A Ramanan, A Riordan, C Roehr, M Wan, E Whittaker.

##### Obstetric working group

L Chappell (coordinator), K Hodson, M Knight, S Pavord, C Williamson.

##### Clinical support

*NHS Lothian Out of Hours support line team* M Odam (coordinator), P Black, B Gallagher, L MacInnes, R O'Brien, K Priestley, A Saunderson; *Clinical Trial Service Unit Out of Hours clinical support* L Bowman, F Chen, R Clarke, M Goonasekara, R Haynes, W Herrington, P Judge, M Mafham, S Ng, D Preiss, C Reith, E Sammons, D Zhu.

##### Health records

*NHS DigiTrials, Southport* H Pinches, P Bowker, V Byrne-Watts, G Chapman, G Coleman, J Gray, A Rees, MJ Landray, M Mafham, N Mather, T Denwood; *Intensive Care National Audit & Research Centre, London* D Harrison; *National Records of Scotland* G Turner; *Public Health Scotland* J Bruce; *SAIL Databank, University of Swansea* C Arkley, S Rees.

##### Drug supply

Public Health England and Department of Health and Social Care (DHSC) Vaccines & Countermeasures teams, DHSC Medicines Supply Contingency Planning Team, NHS England, NHS Improvement, Movianto UK Ltd, Supply Chain Coordination Ltd.

##### Local Clinical Centre RECOVERY trial staff

(listed in descending order of the number of patients randomised per site)

**University Hospitals Of Leicester NHS Trust** C Brightling (PI), N Brunskill (Co-PI), M Wiselka (Co-PI), S Adenwalla, P Andreou, P Bakoulas, S Bandi, S Batham, T Beaver, K Bhandal, M Bourne, L Boyles, M Cannon, A Charalambou, C Kay Cheung, R Cotter, S Diver, A Dunphy, O Elneima, J Fawke, J Finch, C Gardiner-Hill, G Genato, M Graham-Brown, C Haines, B Hargadon, H Holdsworth, W Ibrahim, L Ingram, J Antonio Jesus Silva, K Kaul, M Joshi, S Kapoor, J Whang Kim, A Kuverji, K Kyriaki, A Lea, T Lee, A Lewszuk, L Lock, K Macconaill, R Major, H McAuley, P McCourt, D Mullasseril Kutten, A Palfreeman, E Parker, K Patel, M Patterson, R Phillips, L Plummer, R Russell, S Debbie, H Selvaskandan, S Michaela Southin, K Karali Tsilimpari, C Wiesender, A Yousuf.

**Pennine Acute Hospitals NHS Trust** A Ustianowski (PI), J Raw (Co-PI), R Tully (Co-PI), K Abdusamad, Z Antonina, E Ayaz, B Blackledge, P Bradley, F Bray, M Bruce, C Carty, B Charles, G Connolly, C Corbett, S Dermody, L Durrans, E Falconer, J Flaherty, C Fox, D Hadfield, J Harris, L Hoggett, A Horsley, S Hussain, R Irving, P Jacob, D Johnstone, R Joseph, J Julie Melville, P Kamath, T Khatun, T Lamb, H Law, M Lazo, G Lindergard, S Lokanathan, L Macfarlane, S Mathen, S McCullough, P McMaster, D McSorland, B Mishra, S Munt, A Neal, R Newport, G O'Connor, D O'Riordan, I Page, V Parambil, J Philbin, M Tejha Pinjala, C Rishton, M Riste, J Rothwell, M Sam, Z Sarwar, L Scarratt, A Sengupta, H Sharaf, J Shaw, J Shaw, A Slack, D Symon, H T-Michael, A Uriel, O Walton, S Williams,

**North Tees and Hartlepool NHS Foundation Trust** B Prudon (PI), V Jagannathan (Co-PI), C Adams, N Aung, D Barker, B Campbell, V Collins, J Deane, I Fenner, S Gowans, W Hartrey, M Nafei, L Poole, S Purvis, J Quigley, A Ramshaw, L Shepherd, J Skelton, R Srinivasan, R Taylor, M Walker, M Weetman, B Wetherall, S Wild, D Wilson.

**Nottingham University Hospitals NHS Trust** W S Lim (PI), M Ali, A Andrews, L Anderson, S Ashraf, D Ashton, G Babington, G Bartlett, D Batra, L Bendall, N Benetti, T Brear, A Buck, G Bugg, J Butler, J Butler, R F D Cammack, J Cantliff, L Clark, E Connor, P Davies, M Dent, C Dobson, M Fatemi, A Fatemi, L Fleming, J Grundy, J Hallas, L Hodgen, S Hodgson, S Hodgkinson, L Howard, C Hutchinson, B Jackson, J Kaur, E Keddie-Gray, C Khurana, M Langley, L Looby, M Meredith, L Morris, H Navarra, R Nicol, J Oliver, C Peters, B Petrova, Z Rose, L Ryan, J Sampson, G Squires, J Squires, R Taylor, A Thomas, J Thornton, K Topham, S Warburton, S Wardle, H Waterfall, S Wei, T Wildsmith, L Wilson,

**South Tees Hospitals NHS Foundation Trust** D Chadwick (PI), S Armstrong, D Athorne, M Branch, J Brolly, S Brown, J Cheaveau, H Min Chen, Y Chua, N Cunningham, J Dodds, S Dorgan, D Dunn, P Harper, H Harwood, K Hebbron, F Hunt, A Kala Bhushan, P Lambert, D Leaning, T Manders, B McCarron, N Miller-Biot, C Milne, W Mohammad, M Mollet, A Murad, M Owston, J Potts, C Proctor, S Rao, M A Seelarbokus, P Singh, V Srirathan, L Swithenbank, L Thompson, H Wardy, L Wiblin, J Widdrington, J Williams, P Winder, C Wroe.

**Northampton General Hospital NHS Trust** E Elmahi (PI), M Zaman (Co-I), B Abdul, A Abdulmumeen, N Abdulshukkoor, A Adnan, M H Ahammed Nazeer, A Bazli, N Benesh, W Jie Chin, N Cunningham, H Daggett, E Davies, H Enyi, S Fawohunre, N Geoghegan, J Glover, J W Hague, K Hall, C Hallett, K Haresh, W Ul Hassan, J Hewertson, J Hosea, N Zaya Htoon, M Idrees, C Igwe, H Imtiaz, M Irshad, A Ismail, R Jeffrey, J Jith, P Joshi, R Kaliannan Periyasami, A Khalid, M Usman Khalid, R Kodituwakku, P Lopez, A Mahmood, M Malanca, I Tatenda Mapfunde, V Kumar Maruthamuthu, S Masood, S S M E Masood, M Matharu, A Merchant, F Merchant, S Naqvi, R Natarajan, N Natarajan, K Nawaz, O Ndefo, O Ogunkeye, S Paranamana, N Pugh, A Raj, K Rashid, M Rogers, M Saad, G Selvadurai, A Shah, M Shahzeb, N Shrestha, A Singh, K Smith, B Sohail, M Spinks, L Stockham, A Takyi, Y He Teoh, S Ullah, H Vayalaman, S E I Wafa, T Ward, R Watson, D White, L Ylquimiche Melly, R Zulaikha.

**University Hospitals Birmingham NHS Foundation Trust** C Green (PI), T Whitehouse (Co-PI), I Ahmed, N Anderson, C Armstrong, A Bamford, H Bancroft, M Bates, S Begum, M Bellamy, C Bergin, K Bhandal, E Brandl-Salutz, E Buckingham, E Burke, M Carmody, L Cooper, J Daglish, J Dasgin, A Desai, S Dhani, D Dosanjh, H Ellis, D Gardiner, E Forster, E Grobovaite, B Hopkins, D Hull, Y Hussain, J Jones, L Khan, D Lenton, M Lewis, M Lovell, F Lowe,D Lynch, C McGhee, C McNeill, F Moore, A Nilsson, J Nunnick, C Prest, V Price, J Rhodes, J Sale, M Sangombe, H Smith, I Storey, L Thrasyvoulou, K Tsakiridou, D Walsh, S Welch, H Willis, L Wood, J Woodford, G Wooldridge, C Zullo.

**Calderdale and Huddersfield NHS Foundation Trust** P Desai (PI), D Appleyard, D Bromley, N Chambers, S Dale, L Gledhill, J Goddard, J Greig, A Haigh, K Hallas, K Hanson, K Holroyd, M Home, D Kelly, A Maharajh, L Matapure, S Mellor, E Merwaha, H Riley, M Robinson, K Sandhu, K Schwarz, L Shaw, L Terrett, M Usher, A Wilson, T Wood.

**Manchester University NHS Foundation Trust** T Felton (PI), A Abdul Rasheed, T Abraham, A Ahmed, S Akili, P David Gregory Alexander, C Avram, J Banda, M Baptist, R Bazaz, A Bikov, K Birchall, S Bokhari, J Bradley-Potts, G Calisti, S Carley, S Chilcott, C Chmiel, E Church, R Clark, H Dalgleish, A Desai, G Donohoe, H Durrington, C Eades, G Evans, D Faluyi, S Fowler, T Gorsuch, G Grana, G Gray, J Henry, A Horsley, A Hussain, L James, A John, E Johnstone, Z Kausar, A Khan, E Kolakaluri, C Kosmidis, R William Lord, A Mahaveer, L Manderson, G Margaritopoulos, C Mendonca, H Mistry, C Murray, R Norton, N Odell, R

Osborne, A Palacios, A Panes, L Peacock, S Ratcliffe, C Reynard, E Rice, P Rivera Ortega, T Shanahan, A Simpson, J Soren, M Tin, R Tousis, R Wang, C Whitehead, J R Williams.

**North West Anglia NHS Foundation Trust** K Rege (PI), C Agbo, O Akindolie, A Al-Rabahi, R Ambrogetti, K Thu Aung, A Azman Shah, A Obaloluwa Babs-Osibodu, K Bahadori, J Bhayani, T Bond, H Boughton, S Brooks, N Butterworth-Cowin, R Buttery, P Carter, A Maria Catana, L Cave, S Choi, D Corogeanu, R Croysdill, M Davies, J de Souza, N Duff, L Dufour, O Ebigbola, C Eddings, J Faccenda, A Feroz, L Finch, C Freer, P Goodyear, R Gooentilleke, R Gosling, W Halford, S Havlik, T Hoskins, C Huson, M Ishak, H Javed, T Jones, N Kasianczuk, D Kaur, A Kerr, A Khan, G Koshy, A Kozak Eskenazia, S Lahane, M Kadhim Gulam Lami, J Marshall, K McDevitt, N Muru, A M M Mustafa, S Nazir, I Okpala, T Old, G Oleszkiewicz, H Orme, S O'Sullivan, P Paczko, A Pandey, A Patel, S Pathak, S Poon, M I Rather, R Renu Vattekkat, S H M Rizvi, M Samyraju, J Sanyal, S H Shah, E Smith, S Stacpoole, B Theng Tan Tan, N Temple, K Thazhatheyil, M Saif Uddin, N Veale, D Walter.

**Wrightington, Wigan and Leigh NHS Foundation Trust** A Ashish (PI), J Cooper, D Heaton, S Hough, V Parkinson, E Robinson, T Taylor, C Tierney, A Verma, N Waddington, C Williams, C Zipitis.

**Barts Health NHS Trust** S Tiberi (PI), A Aboaba, E Adeyeye, J Agwada-Akeru, F Runa Ali, R Allen, C Ardley, R Astin-Chamberlain, G Bacon, H Baillie, R Batha, B Bloom, M Bolton, C Borra, G Boyapati, R Buchanan, C Chan, C Chitsenga, B Cipriano, P Foster Cofie, M DeLuna, S Elia, K El-Shakankery, A Fikree, A Ghosh, R Goiriz, P Goldsmith, M Gouldbourne, A Grant, L Greenfield, S Grigoriadou, R Gritton, K Gunganah, J Hand, C Harwood, U Hemmila, J Higgins, D Hobden, L Howaniec, D Hsu, S Iliodromiti, S Issa, P Jones, M Juan, J Kassam, C Keith-Jopp, M Khan, C Ryan Khaw, H Kunst, J Lai, I Lee, D Lieberman, S Liebeschuetz, E Magavern, C Maniero, J Maitland, H Malcolm, H Marshall, N Matin, P May, R McDermott, K Menacho, L Millin, A Mohammed, K Moriarty, S Naeem, T Newman, C Nic Fhogartaigh, M Omar, A Pakozdi, M Parrott, P Pfeffer, J Pott, J Powell, W Ricketts, A Riddell, P Rughani, N Sahdev, V Sarodaya, B Selvarajah, I Skene, K Denise Smallshaw, A So, D Stevenson, C Suarez, T Swaine (Associate PI), S Thomas, A Thomas, J Thomson, N Thorn, C Tierney, S Ullah, R Vathenen, L Velauthar, K Ward, K Wiles, P Woodland, S Youssouf, A Zdanaviciene.

**Cardiff & Vale University LHB** C Fegan (PI), A Balan, B Basker, S Bird, Z Boult, V Britten, L Broad, H Cendl, M Chakraborty, J Cole, M Edgar, T Evans, J Evans, M Evans, J Forton, S Frayling, F Greaves, S Harrhy, M Haynes, H Hill, Z Hilton, L Jones, S Jorgensen, A Kelly, L Knibbs, D Lau, E Maureen, J May, E McGough, A McQueen, J Milner, R Norman, K Nyland, C Oliver, K Paradowski, M Patal, K Rahilly, C Robinson, S Scourfield, M Starr, S Struik, E Thomas, R Thomas-Turner, G Williams, J Williams, M Williams, S Zaher.

**Liverpool University Hospitals NHS Foundation Trust** P Hine (PI), S Todd (Co-PI), I Welters (Co-PI), D Wootton (Co-PI), M Ahmed, R A Ahmed, A Al Balushi, D Barr, A Bennett, H Bond, C Burston, J Byrne, L Chambers, D Coey, T Cross, J Cruise, K Fenlon, S Gould, K Haigh, C Hall, M Harrison, P Hazenberg, S R Hicks, S Hope, K Hunter, A Islim, S Iyer, A Jackson, F Jaime, L Keogan, K Knowles, K Krasauskas, J Lewis, P Lopez, C Lowe, A Mediana, M Middleton, M Nugent, L Pauls, S Pringle, I Quayle, S Raghunath, M Riley, J Sedano, D Shaw, C Smith, S Stevenson, A Stockdale, R Tangney, J Tempany, V Waugh, K Williams.

**Northumbria Healthcare NHS Foundation Trust** B Yates (PI), C Ashbrook-Raby, H Campbell, D Charlton, S Dodds, V Ferguson, T Hall, I Hamoodi, P Heslop, J Luke, N McLarty, L Patterson, S Pick, J Reynolds, S Robinson, C Tanney, L Taylor, L Winder, C Walker.

**Countess Of Chester Hospital NHS Foundation Trust** S Scott (PI), M Abouibrahim, M Ahmad, S Haris Ahmed, A Ajibode, L Alomari, A Asrar, E Austin, P Bamford, A Barclay, L

Barker, K Barker-Williams, W Barnsley, H Batty, I Benton, S Billingham, S Brearey, S Brigham, V Brooker, C Burchett, M Burgess, R Cade, F Cameron, R Cannan, K Cawley, N Chavasse, Z Cheng, R Clarke, E Cole, C Cotton, A Davidson, Y Doi, C Dragos, L Nicola Ellerton, L Gamble, M Grant, J Grounds, H Hodgkins, M Shoaib Irshad, M Iyer, A Johari, C Jones, N Kearsley, B Lim, D Kevin Llanera, S Yin Loh, E London, E Martin, P Maskell, J McBurney, M McCarthy, R McEwen, E Meeks, G Metcalf-Cuenca, S Middleton, L Mihalca-Mason, M Nayyar, F Naz, E Okpo, H Claire Parry, S Anthony Pearson, D Phillips, C Pickering, A Ponnuswamy, V Prescott, J Prince, S Ur Rahman, S Scott, C Steele, C Thorne, S Tomlin, T Trussell, T Webster, L Zammit, E Thant Zin.

**North Cumbria Integrated Care NHS Foundation Trust** C Graham (PI), A Abdelaziz, O Ali, J Atkinson, G Bell, C Brewer, M Clapham, J Gregory, S Hanif,R Harper, M Lane, A McSkeane, U Poultney, K Poulton, S Pritchard, S Shah, C Smit, P Tzavaras, V Vasadi, A Wilson, T Wilson, D Zehnder.

**Luton and Dunstable University Hospital NHS Foundation Trust** D Shaw (PI), S Tariq (Co-PI), N Ahmed, S Ali, S Allen, M Alzetani, C Ambrose, K Aneke, T Angel, Z Myo Aung, R Banerjee, T Baqai, A Batla, M Bergstrom, S Bhakta, N Bibi, T Chapman, A David, L Dirmantaite, M Edmondson, E Elfar, M Magdy Elgamal, H El-Sbahi, D Fishman, C Fornolles, T Forshall, A Francioni, S Gent, N George, A Ibrahim, A Ingram, R James, K Kabiru Dawa, F Khan, M Aamaz Khan, S Lee, C Lingam, C Nisha Luximon, N Marcus, M Masood, R Mejri, A Moharram, C Moss, G Naik, A Yong Kheng Cordero Ng, L Nicholls, M Nisar, V Parmar, F Prasanth Raj, V Puisa, V Quick, B Ramabhadran, A Reddy, N Riaz, B Rudran, S Sabaretnam, H Sagoo, S Sarma, K Savlani, P Shah, D Shaw, S Soo, P Sothirajah, I Southern, M Louise Tate, C Travill, V Singh Uppal, W Wakeford,

**Portsmouth Hospitals NHS Trust** T Brown (PI), J Andrews,M Baker-Moffatt, A Bamgboye, D Barnes, S Baryschpolec, L Bell, J Borbone, M Broadway, F Brogan, R Bungue-Tuble, K Burrows, A Chauhan, M Chauhan, E Cowan, A Darbyshire, M Davey, M David, J Denham, H Downe, C Edwards, L Fox, Z Garner, B Giles, A Gribbin, Y Harrington-Davies, L Hawes, A Hicks, E Hossain, S Howe, B Jones, C Lameirinhas, B Longhurst, M Mamman, S McCready, C Minnis, M Moon, J Mouland, L Murray, S Rose, H Rupani, M Rutgers, T Scorrer, K Scott, R Thornton, A Tiller, C Turner, M Wands, L Watkins, M White, L Wiffen (Associate PI), J Winter.

**Epsom and St Helier University Hospitals NHS Trust** S Winn (PI), R Wake (Co-PI), S Ahamed Sadiq, A Aldana, B Al-Hakim, K Ansu Agyapong, G Azzopardi, R Chicano, I Chukwulobelu, N Colbeck, N Cole, R Dogra, E Doherty, A Elradi, J Emberton, T J Evans, R Ganapathy, M Haque, R Hayre, S Jain, K Jian, L Johnson, A Johnson, J Kotecha, A Kundu, D Langer, Y Mashhoudi, K Mathias, ME Maxan, F Mellor, M Morgan, S Nafees, V Bharadwaj Palagiri Sai, M Phanish, H Rana Rahimi, S Ramanna, J Ratoff, S Rozewicz, TDL Samuel, B Shah, S Shahnazari, R Shail, A Sharif, S Somalanka, R Suckling, P Swift, V Tyagi, N Vilimiene, C Wells.

**Mid Cheshire Hospitals NHS Foundation Trust** D Maseda (PI), C Ball, K Best, G Bridgwood, R Broadhurst, C Brockelsby, T Brockley, J Brown, R Bujazia, S Clarke, J Cremona, C Dixon, S Dowson, H Drogan, F Duncan, C Emmett, M Emms, H Farooq, D Fullerton, C Gabriel, S Hammersley, R Hum, T Jones, S Kay, E Kelly, M Kidd, D Lees, R Lowsby, E Matovu, K McIntyre, H Moulton, K Nourein, M O'BrianK Pagett, A Ritchings,S Smith, J Taylor, K Thomas, K Turbitt, S Yasmin.

**Chesterfield Royal Hospital NHS Foundation Trust** N Spittle (PI), N Weatherly (Co-PI), S Beavis, S Beghini, L Blundell, J Bradder, J Cort, J Cresswell, K Dale, A Foo, J Gardner, R Gascoyne, E Hall, M Kelly-Baxter, E Mackay, K Moxham, A Padmakumar, K Pritchard, J Salmon, A Smith, V Sorice, L Stevenson, A Whileman, E Wolodimeroff.

**Frimley Health NHS Foundation Trust** M Meda (PI), J Democratis (Co-PI), N Barnes, N Brooks, L Chapman, J da Rocha, R Dolman, A Edwards, T Foster, F Fowe, E Gaywood, S Gee, S Jaiswal, M Molloholli, A Raguro, F Regan, L Rowe-Leete, C Smith, M van de Venne, T Weerasinghe.

**University Hospitals Of Derby and Burton NHS Foundation Trust** T Bewick (PI), P Daniel (Co-PI), U Nanda (Co-PI), G Bell, C Downes, K English, A Fletcher, J Hampson, M Hayman, N Jackson, A Jane Matthews, S Ohja, L Prince, J Radford, K Riches, G Robinson, A Sathyanarayanan, F Scothern, L Wilcox, L Wright.

**Oxford University Hospitals NHS Foundation Trust** K Jeffery (PI), M Ainsworth, C Arnison-Newgass, A Bashyal, S Beer, A Bloss, D Buttress, W Byrne, A Capp, P Carter, P Cicconi, R Corrigan, C Coston, L Cowen, N Davidson, L Downs, J Edwards, R Evans, S Gardiner, D Georgiou, A Gillesen, A Harin, M Havinden-Williams, R Haynes, C Hird, A Hudak, P Hutton, R Irons, P Jastrzebska, S Johnston, M Kamfose, K Lewis, T Lockett, F Mendoza Maria del Rocio, J Carlos Martinez Garrido, S Masih, A Mentzer, S Morris, C O'Callaghan, Z Oliver, S Paulus, E Perez, L Periyasamy, L Peto, D Porter, S Prasath, C Purdue, M Ramasamy, CC Roehr, A Rudenko, V Sanchez, A Sarfatti, M Segovia, T Sewdin, J Seymour, V Skinner, L Smith, A Sobrino Diaz, M Taylor-Siddons, H Thraves, C Tsang, M Vatish, Y Warren, E Wilcock.

**Dartford and Gravesham NHS Trust** B Khan (PI), D Ail,R Aldouri, G Awadzi, B Bassoy, R Bhalla, J Billings, S Bokhari,G Boniface, J Cernova, T Chen, P Chimbo, N Chitalia, S Danso-Bamfo, D Depala, A Dhanoa, T Edmunds, E Fernandez, T Ferrari, B Fuller, A Gherman, R Heire, L Ilves, L Lacey, E Lawrence, M Lewis, A Maric, W Martin, Z Min, C Newman, R Nicholas, O Olufuwa, N Pieniazek, T Qadeer, S Rathore, S Sathianandan, C Scott, A Shonubi (Associate PI), S Siddique, G Sisson, M Soan, D Streit, C Stuart, M Szekely, W Umeojiako, S Urruela, B Warner, M Waterstone, S White, K Yip, A-M Zafar, S Zaman.

**Buckinghamshire Healthcare NHS Trust** R West (PI), J Abrams, A Baldwin, O Bannister, J Barker, H Beddall, H Blamey, E Chan, J Chaplin, B Chisnall, C Cleaver, S Crotty, P Dey, L Downs, M Kononen, S Kudsk-Iversen, A Kudzinskas, M Laurenson, J Mandeville, S McLure, E Morgan-Smith, A Ngumo, R Oxlade, A Parekh (Associate PI), M Rahman, C Robertson, S Shah, J Tebbutt, N Vella, M Veres, A J R Watson, N Wong, M Zammit-Mangion, M Zia.

**NHS Lothian: Royal Infirmary of Edinburgh** A Gray (PI), K Baillie (Co-I), M Adam, A Anand, R Anderson, D Baird, T Balaskas, J Balfour, P Black, C Blackstock, S Brady, R Campbell, J Carter, P Chapman, C Cheyne, A Christides, D Christmas, L Crisp, D Cryans, J Dear, K Dhaliwal, M Docherty, R Dodds, L Donald, S Dummer, M Eddleston, N Fethers, E Foster, R Frake, E Gaughan, D Gilliland, E Godson, J Grahamslaw, A Grant, N Grubb, S Hainey, Z Harding, M Harris, M Harvey, D Henshall, S Hobson, N Hunter, Y Jaly, J Jameson, D Japp, H Htet Ei Khin, L Kitto, S Krupej, C Langoya, R Lawrie, A Lloyd, B Lyell, D Lynch, J Macfarlane, L MacInnes, A MacRaild, M Marecka, A Marshall, M Martin, C McCann, F McCurrach, E Moatt, W Morley, M Morrissey, D Newby, K Nizam Ud Din, R O'Brien, E O'Sullivan, M Odam, A Peterson, P Phelan, G Pickering, T Quinn, N Robertson, L Rooney, N Rowan, R Salman, E Small, P Stefanowska, A Stevenson, S Stock, A Summers, J Teasdale, I Walker, K Walker, A Williams.

**Wirral University Teaching Hospital NHS Foundation Trust** A Wight (PI), L Bailey, S Owais Bokhari, S Brownlee, A Bull, J Corless, C Denmade, N Ellard, A Farrell, A Hufton, R Jacob, K Elizabeth Jones, H Kerss, J McEntee, N Morris, R Myagerimath, T Newcombe, M Parsonage, H Peake, D Pearson, R Penfold, S Rath, R Saunders, A Sharp, B Spencer, A Suliman, S Sutton, H Tan, D Tarpey, L Thompson, T Thornton, E Twohey, D Wagstaff, Z Wahbi, S Williams.

**Barking, Havering and Redbridge University Hospitals NHS Trust** M Phull (PI), A Umaipalan (Co-PI), J Ah-Chuen, A Ainsley, G Baijiu, A Basumatary, C Calderwood, P Dugh, K Dunne, P Greaves, K Hunt, M Islam, V Katsande, MR Khan, S King, A Loverdou, A McGregor, A Misbahuddin, A Mohamed, N Nagesh, D Nicholls, N O'Brien, L Parker, T Pogreban, L Rosaro, E Salciute, M Sharma, H Smith, E Visentin, L Walshaw.

**Bradford Teaching Hospitals NHS Foundation Trust** D Saralaya (PI), N Akhtar, V Beckett, L Brear, V Drew, J Eedle, N Hawes, S Kmachia, S Moss, S Oddie, J Paget, K Regan, D Ryan-Wakeling, A Shenoy, K Storton, R Swingler, J Syson, J Todd, R Wane, A Wilson.

**South Tyneside and Sunderland NHS Foundation Trust** E Fuller (PI), A MacNair (Co-PI), C Brown, A Burns, C Caroline, R Davidson, M Dickson, B Duncan, N Elkaram, I Emmerson, L Fairlie, M Hashimm, J Henderson, K Hinshaw, J Holden, R Hovvels, S Laybourne, P Madgwick, K Martin, M McKee, J Moore, N Mullen, P Murphy, L Palmer, G Parish, M Rangar, R Shahrul, A Smith, L Smith, M Smith, B Stidolph, L Terry, A Trotter, F Wakinshaw, E Walton, M Walton.

**Royal Berkshire NHS Foundation Trust** M Frise (PI), R Arimoto, S Black, C Camm, H Coles, H De Berker, A Fries, E Gabbitas, N Hasan, Z Htet, N Jacques, S Kashif, L Keating, S Leafe, Z Milne, B Mitchell, T Okeke, W Orchard, A Parmar, M Raffles, N Shields, M Thakker, M Thomas, H Wakefield, A Walden.

**North Middlesex University Hospital NHS Trust** J Moreno-Cuesta (PI), S Rokadiya (Co-PI), A Govind, A Haldeos, K Leigh-Ellis, C van Someren, R Vincent, L Walker.

**Basildon and Thurrock University Hospitals NHS Foundation Trust** K Thomas (PI), T George (Co-PI), E Cannon, A Ikomi, L Jayasekera, L Kittridge, G Maloney, B Manoharan, M Mushabe, A Nicholson, A Pai, J Riches, J Samuel, N Setty, A Solesbury, D Southam, S Tisi, M Vertue, K Wadsworth, B Yung.

**County Durham and Darlington NHS Foundation Trust** J Limb (PI), V Atkinson, M Birt, E Brown, A Cowton, V Craig, D Egginton, D Fernandes, A Ivy, D Jayachandran, J Jennings, A Kay, M Kent, S McAuliffe, S Naylor, G Nyamugunduru, J O'Brien, K Postlethwaite, K Potts, P Ranka, G Rogers, S Sen, J Temple, S Wadd, H Walters, J Yorke.

**London North West University Healthcare NHS Trust** A Whittington (PI), M Adamus, J Barrett, F Cawa, S Chhabra, S Chita, E Dhillon, S Ann Filson, J Goodall, R Gravell, A Gupta-Wright, S Gurram, H Houston, G Hulston (Associate PI), S Isralls, C Kukadiya, M Lang, J Mae Low, C Macleod, J Milburn, K Muralidhara, B Nayar, O Ojo, P Papineni, V Parris, M Patel, S Quaid, J Sethi, A Sturdy (Associate PI), B Tyagi, L Cheyenne Vaccari, N van der Stelt, G Wallis, E Watson.

**Medway NHS Foundation Trust** R Sarkar (PI), I Ahmed, S Ahmed, S Ambler, F Babatunde, S Banerjee, N Bhatia, L Brassington, F Brokke, D Bruce, B Cassimon, R Singh Chauhan, A Chengappa, N Divikar, C Donnelly, C Froneman, C Gnanalingam, T Gower, H Harizaj, G Hettiarachchi, M Hollands, S Jansz, B Josiah, M Kamara, S Kidney, T Kyere-Diabour, K Lewiston, N Miah, S Millington, L Mires, A Mitchell, C Mizzi, K Naicker, I Petrou, M Mubeen Phulpoto, A Ross-Parker, I Ramadan, A Roy, A Ryan, E Samuels, T Sanctuary, A Sharma, S Singham, J Sporrer, W Ul Hassan, P Vankayalapati, B Velan, L Vincent Smith, E Vyras, J Wood, N Zuhra (Associate PI).

**Sandwell and West Birmingham Hospitals NHS Trust** S Clare (PI), M Cecilia Ahmed, Y Beuvink, K Blachford, S Clamp, J Colley, P De, M Fenton, B Gammon, A Hayes, L Henry, S Hussain, S Joseph, F Kinney, T Knight, R Kumar, W Leong, T Lim, B Mahay, Y Nupa, A Orme, W Osborne, Z Pilsworth, S Potter, S Prew, N Rajaiah, A Rajasekaran, H Senya, N Shah, N

Shamim, S Sivakumar, L Smith, P Thozthumparambil, N Trudgill, A Turner, L Wagstaff, S Willetts, H Willis, M Yan.

**East Kent Hospitals University NHS Foundation Trust** N Richardson (PI), A Alegria (Co-PI), R Kapoor (Co-PI), K Adegoke, L Allen, E Beranova, G Boehmer, N Crisp, J Deery, J Hansen, A Elgohary, T Elsefi, C Hargreaves, T Hazelton, R Hulbert, A Ionita, A Knight, C Linares, D Loader, E Matisa, J McAndrew, M Montasser, A Moon, C Oboh, P Offord, C Price, A Rajasri, J Rand, N Schumacher, D Stephensen, S Stirrup, L Tague, S Tilbey, S Turney, V Vasu, M Venditti, H Weston, Z Woodward.

**University Hospitals Of North Midlands NHS Trust** T Kemp (PI), J Alexander, W Al-Shamkhani, N Bandla, A Bland, N Bodasing, A Cadwgan, M Yafaa Naveed Chaudhary, L Yin Cheng, S Rebecca Church, F Clark, M Davies, L Diwakar, E Eaton, M Evans, A Farmer, F Farook, M Gellamucho, K Glover, M Haris, J Humphries, I Hussain, S Khan, L Korcierz, J Lee, J Machin, J Marshall, H McCreedy, G Muddegowda, I Mustapha, K Nettleton, Z Noori, W Osman, H Parker, N Patel, A Quinn, M Ram, A Remegoso, T Scott, N Sheikh, R Swift, C Thompson, J Tomlinson, H Turner, L Walker, J Weeratunga, C White, J White, P Wu.

**Aneurin Bevan University LHB** T Szakmany (PI), S Champanerkar (Co-I), A Griffiths (Co-I), A Alina Ionescu (Co-I), C Somashekar (Co-I), K Zalewska (Co-I), L Aitken, E Baker, D Barnett, M Brouns, J Cann, S Cherian, R Codd, S Cutler, W Davies, A Dell, M Edwards, S Edwards, S Fairbairn, N Hawkins, S Hodge, G Hodkinson, R Hughes, C Ivenso, T James, M Jones, S Jones, A Lucey, J MacCormac, G Marshall, S McKain, H Nassa, J Northfield, S Palmer, L Peter, C Price, M Pynn, A Roynon-Reed, L Shipp, J Singh, M Singh, K Swarnkar, P Torabi, R Venkataramakrishnan, E Wall, A Waters, K Wild, M Winstanley, K Wyness.

**East Suffolk and North Essex NHS Foundation Trust** V Kushakovsky (PI), M Ramali (Co-PI), S Alam, S Bartholomew, A Bataineh, D Beeby, S Bell, N Broughton, C Buckman, C Calver, J Campbell, C Chabo, M Chowdhury, K Cooke, N Deole, C Driscoll, A Elden, H Eldew, N Entwistle, F Farnworth, R Francis, E Galloway, M Garfield, A Ghosh, G Gray, P Greenfield, M Hadjiandreou, H Hewer, M S Hossain, R Howard-Griffin, A Islam, E Jamieson, Z Jiao, K Johannessen, S Han Lee, R Lewis, R Lloyd, L Andrew Mabelin, D Morris, S Nallapareddy, H C Ooi (Associate PI), R Osagie, C Parkinson, H Patel, H Prowse, B Purewal, P Ridley, V Rivers, J Rosier, S Sharma, A Sheik, R Skelly, R Smith, R Sreenivasan, A Taylor, P Tovey, A Turner, K Turner, K Vithian, J Zhixin.

**Surrey and Sussex Healthcare NHS Trust** E Potton (PI), N Jain (Sub-I), A Khadar (Sub-I), P Morgan (Sub-I), J Penny (Sub-I), E Tatam (Sub-I), S Abbasi, D Acharya, A Acosta, L Ahmed, S Ali, M Alkhusheh, V Amosun, A Arter, M Babi, J Bacon, K Bailey, N Balachandran, S Bandyopadhyam, L Banks, J Barla, T Batty, S Bax, A Belgaumkar, G Benison-Horner, A Boles, N Broomhead, E Cetti, C Chan, I Chaudhry, D Chudgar, J Clark, S Clueit, S Collins, E Combes, G Conway, O Curtis, M Das, M Daschel, S Davies, E Potton, S Abbasi, D Acharya, A Acosta, L Ahmed, S Ali, M Alkhusheh, V Amosun, A Arter, M Babi, J Bacon, K Bailey, N Balachandran, S Bandyopadhyam, L Banks, J Barla, T Batty, S Bax, A Belgaumkar, G Benison-Horner, A Boles, N Broomhead, E Cetti, C Chan, I Chaudhry, D Chudgar, J Clark, S Clueit, S Collins, E Combes, G Conway, O Curtis, M Das, M Daschel, S Davies, A Day, M Dhar, K Diaz-Pratt, C Dragan, H Dube, V Duraiswamy, J Elias, A Ellis, T-Y Ellis, J Emmanuel, A Engden, Y Fahmay, B Field, K Fishwick, U Ganesh, C Gilbert, T Giokanini-Royal, E Goudie, S Griffith, S Gurung, R Habibi, C Halevy, A Haqiqi, R Hartley, A Hayman, J Hives, M Horsford, S Hughes, C Hui, R Hussain, C Iles, L Jackson, A James, D Jayaram, E Jessup-Dunton, T Joefield, N Khan, W Kieffer, E Knox, V Kumar, R Kumar, V Kurmars, H Lafferty, F Lamb, R Layug, N Leitch, W Lim, U Limbu, R Loveless, M Mackenzie, N Maghsoodi, S Maher, M Maljk, I Man, N McCarthy, B Mearns, C Mearns, K Morgan-Jones, G Mortem, G Morton, B Moya, G Murphy, S Mutton, A Myers, T Nasser, J Navaratnam, S Nazir, S Nepal, K Nimako, L Nimako, C O'Connor, A Patel, K Patel, V Phongsathorn, PA Pillai, M Poole, N Qureshi, S Ranjan, A

Rehman, T Samuels, E Scott, G Sekadde, A Sharma, G Sharp, S Shotton, O Simmons, P Singh, S Smith, K Sri Paranthamen, S Suresh, K Thevarajah, L Thomas, H Timms, N Tomasova, S Tucker, S Vara, C Vaz, S Weller, J White, M Wilde, I Wilkinson, C Williams, M Win, D Woosey, D Wright.

**Sherwood Forest Hospitals NHS Foundation Trust** M Roberts (PI), K Amsha(Co-I), G Cox (Co-I), N Downer (Co-I), D Hodgson (Co-I), J Hutchinson (Co-I), S Kalsoom (Co-I), A Molyneux (Co-I), Z Noor (Co-I), L Allsop, T Brear, P Buckley, L Dunn, M Gill, C Goodwin, C Heeley, M Holmes, R Holmes, E Langthorne, C Moulds, D Nash, J Rajeswary, S Shelton, K Slack, S Smith, B Valeria, I Wynter, M Yanney, I Ynter,

**York Teaching Hospital NHS Foundation Trust** J Azam (PI), KJ Chandler (PI), M Abdelfattah, A Abung, J Anderson, P Antill, S Appleby, P Armtrong, T Berriman, D Bull, O Clayton, A Corlett, D Crocombe, S Davies, K Elliott, L Fahel, J Ghosh, N Gott, D Greenwood, T Holder, P Inns, W Lea, E Lindsay, K Mack, N Marshall, S McMeekin, T Momoniat, P Nikolaos, M O'Kane, G Patrick, H Pearson, P Ponnusamy, A Poole, N Price, R Proudfoot, G Purssord, S Roche, C Sefton, S Shahi, D Smith, B Sohail, R Thomas, A Turnbull, L Turner, H J Watchorn, E Wiafe, J Wilson, D Yates.

**The Royal Bournemouth and Christchurch Hospitals NHS Foundation Trust** M Schuster Bruce (PI), D Baldwin, Z Clark, M Dale, D Griffiths, E Gunter, S Horler, T Joyce, M Keltos, S Kennard, N Lakeman, L Mallon, R Miln, S Nix, S Orr, S Pitts, L Purandare, L Rogers, J Samways, E Stride, L Vamplew, L Wallis.

**University Hospitals Bristol and Weston NHS Foundation Trust** J Willis (PI), N Blencowe (Co-PI), P Singhal (Co-PI), M Abraham, B Al-Ramadhani, A Archer, G Aziz, A Balcombe, K Bateman, M Baxter, L Beacham, K Belfield (Associate PI), N Bell, M Beresford, J Bernatoniene, D Bhojwani, S Biggs, J Blazeby, K Bobruk, N Brown, L Buckley, P Butler, C Caws, E Chakkarapani, K Chatar, B Chivima, C Clemente de la Torre, K Cobain, D Cotterill, E Courtney, S Cowman, K Coy, H Crosby, K Curtis, P Davis, O Drewett, H Dymond, K Edgerley, M Ekoi, M Elokl, B Evans, T Farmery, N Fineman, A Finn, L Gamble, F Garty, B Gibbison, L Gourbault, D Grant, K Gregory, M Griffin, R Groome, M Hamdollah-Zadeh, A Hannington, J Heywood, A Hindmarsh, N Holling, R Houlihan, J Hrycaiczuk, H Hudson, K Hurley, R Jarvis, B Jeffs, A Jones, R E Jones, E King-Oakley, E Kirkham, R Kumar, M Kurdy, L Kyle (Associate PI), S Lang, L Leandro, H Legge, F Loro, A Low, H Martin, L McCullagh, G McMahon, L Millett, K Millington, J Mok, J-H Moon, L Morgan, S Mulligan, C O'Donovan, E Payne, C Penman, J Pickard, C Plumptre, A Ramanan, J Ramirez, S Ratcliffe, J Robinson, M Roderick, S Scattergood, A Schadenberg, R Sheppeard, C Shioi, D Simpson, A Skorko, R Squires, M Stuttard, P Sugden, S Sundar, T Swart, E Swift, K Thompson, K Turner, S Turner, A Tyer, S Vergnano, R Vincent, R Ward, S Wilkinson, J Williams, S Williams, J Willis, H Winter, L Woollen, R Wright, A Younes Ibrahim.

**Sheffield Teaching Hospitals NHS Foundation Trust** P Collini (PI), A Ang, J Belcher, L Chapman, K Chin, D Cohen, J Cole, HE Colton, R Condliffe, M Cribb, S Curran, T Darton, D de Fonseka, T de Silva, A Dunn, E Ferriman, R Anne Foster, J Greig, J Hall, M Ul Haq, S Hardman, E Headon, C Holden, L Hunt, E Hurditch, F Kibutu, T Kitching, L Lewis, T Locke, L Mair, P May, J Meiring, J Middle, JT Middleton, P Morris, T Newman, L Passby, R Payne, G Farid Rana, S Renshaw, A Rothman, D Sammut, S Sherwin, P Simpson, M Sterrenburg, B Stone, M Surtees, A Telfer, R Thompson, N Vethanayagam, R West, T Williams.

**University Hospitals Coventry and Warwickshire NHS Trust** K Patel (PI), C Imray (Co-PI), N Aldridge, A Campbell, G Evans, E French, R Grenfell, S Hewins, D Hewitt, J Jones, R Kumar, E Mshengu, S Quenby, K Read, P Satodia, E Sear, M Truslove,

**Great Western Hospitals NHS Foundation Trust** A Kerry (PI), A Beale, A Brooks, C Browne, J Butler, J Callaghan, B Chandrasekaran, C Coombs, R Davies, L Davies, T Elias, E Fowler, G Gowda, J Gregory, A Ipe, A Jaffery, Q Jones, L Kyeremeh, I Laing-Faiers, H Langton, C Lewis-Clarke, C Mackinlay, P Mappa, A Maxwell, W Mears, E Mousley, T Onyirioha, L Pannell, S Peglar, A Pereira, J Pointon, I Ponte Bettencourt dos Reis, E Price, A Quayle, Q Qurratulain, S Small, H Smith, E Stratton, M Tinkler, A van der Meer, E Wakefield, R Waller, M Walton, M Watters, L Whittam, T Wiliams, Z Xia, K Yein, V Zinyemba.

**Blackpool Teaching Hospitals NHS Foundation Trust** J Cupitt (PI), N Ahmed, O Assaf, A Barnett, L Benham, P Bradley, Z Bradshaw, M Brunton, T Capstick, M Caswell, V Cunliffe, R Downes, N Latt, R McDonald, A Mulla, E Mutema, J Navin, A Parker, S Preston, N Slawson, E Stoddard, S Traynor, V Vasudevan, E Ward, S Andrew Warden, J Wilson, A Zmierczak,

**University Hospitals Of Morecambe Bay NHS Foundation Trust** S Bari (PI), A Higham (Co-PI), M Al-Jibury, K Allison, V Anu, C Bartlett, S Bhuiyan, L Bishop, K Burns, A Davies, A Fielding, M Gorst, C Hay, J Keating, T Khan, F Mahmood, P Mallinder, S Peters, D Power, J Ritchie, K Simpson, H Spickett, C Stokes, H Thatcher, A Varghese, T Wan, F Wood.

**Southern HSC Trust** R Convery (PI), J Acheson, J Brannigan, D Cosgrove, C McCullough, D McFarland, R McNulty, S Sands, O Thompson.

**Bolton NHS Foundation Trust** M Balasubramaniam (PI), C Subudhi (Co-PI), C Acton, A Ajmi, R Ahmed, A Ajmi, A Al-Asadi, A Amin, A Bajandouh, M Bhalme, Z Carrington, J Chadwick, S Cocks, C Dawe, A Eusuf, S Farzana, P Hill, R Holmes, G Hughes, R Hull, K Ibrahim, M Ijaz, R J Kalayi, S Khurana, S Latham, K Lipscomb, JP Lomas, N Natarajan, D Nethercott, P Nicholas, D Obeng, V Priyash, K Rhead, M Saleh, O Sharma, Z Shehata, J Shurmer (Associate PI), R Sime, S Singh, R Smith, E Tanton, D Tewkesbury, P Thet, S Thornton, N Wang, M Watts, I Webster.

**Hull University Teaching Hospitals NHS Trust** N Easom (PI), K Adams, L Baldwin, G Barlow, R Barton, H Bexhell, D Clark, P Gunasekera, A Harvey, M Hayes, M Ivan, A James, X Kassianides, S Khan, M Kolodziej, P Lillie, V Mathew, S Mongolu, I A Muazzam, P O'Reilly, T Perinpanathan, C Philbey, B Pickwell-Smith, L Rollins, A Samson, T Sathyapalan, L Sherris, K Sivakumar, T Taynton, H Yates.

**United Lincolnshire Hospitals NHS Trust** M Chablani (PI), R Barber (Co-PI), S Archer, S Beck, S Butler, A Chingale, C Flood, O Francis, C Hewitt, A Hilldrith, A Kirkby, R Mishra, K Netherton, M Okubanjo, L Osborne, A Reddy, A Sloan.

**King's College Hospital NHS Foundation Trust** M McPhail (PI), D Rao (Co-PI), J Adeyemi, J Aeron-Thomas, M Aissa, F Batalla, S Mae Candido, K Clark, K El Bouzidi, J Galloway, N Griffiths, A Gupta, Y Hu, E Jerome, N Kametas, N Long, H Martin, V Patel, T Pirani, S Ratcliff, S Rodrigues, B Sari, N Sikondari, J Smith, C Soto, A Te, R Uddin, J Vidler (Associate PI), M Waller, M West, C Williamson, M Yates, A Zamalloa.

**Kettering General Hospital NHS Foundation Trust** N Siddique (PI), M Abedalla, A Ul Amna Abeer Ul Amna, K Adcock, S Adenwalla, OA Adesemoye, RA Ali, A Ali, MZ Ashraf, MB Ashraq, S Ashton, H Asogan, ACT Aung, H Aung, AM Baral, FS Bawani, S Beyatli, S Coburn, J Dales, R Deylami, A Elliott, S Gali, A Gkioni, L Hollos, M Hussam El-Din, A Ibrahim, Y Jameel, G Keyte, J Khatri, RS Kusangaya, S Little, A Madu, G Margabanthu, M Mustafa, H Naeem, D Ncomanzi, G Nikonovich, A Nisha James, Y Owoseni, N Pandian, D Patel, D Patel, M Raceala, D Ramdin, S Saunders, AY Shaikh, S Stapley, S Sudershan, N Veli, A Mubashar Virk, A Wazir, S White, J Wood, N Zakir.

**NHS Greater Glasgow and Clyde: Glasgow Royal Infirmary** K Puxty (PI), H Bayes (Co-I), J Alexander, L Bailey, A Begg, S Carmichael, S Cathcart, R Colbert, I Crawford, A Dougherty, K Gardiner, T Grandison, N Hickey, J Ireland, A Jamison, D Jenkins, J Johnstone, L Martin, M McIntyre, A Munro, S Nelson, H Peddie, G Piper, L Pollock, D Rimmer, G Semple, S Thornton.

**Royal Free London NHS Foundation Trust** B Caplin (PI), H Tahir (Co-PI), R Abdul-Kadir, I Alshaer, M Anderson, G R Badhan, S Bhagani, E Cheung, V Conteh, E Damian, R Davies, H Hughes, V Jennings, V Krishnamurthy, A Kurani (Associate PI), H Mahdi, S O'Farrell, P Patel, T Sobande.

**Gateshead Health NHS Foundation Trust** R Allcock (PI), M Armstrong, J Barbour, M Bokhar, J Curtis, A Dale, V Deshpande, I Hashmi, E Johns, L M Jones, R King, E Ladlow (Associate PI), R Mackie, D Mansour, B McClelland, W McCormick, C McDonald, C Moller-Christensen, R Petch, S Razvi, R Sharma, L Southern, G Stiller, E Watkins, H Wilkins.

**The Newcastle Upon Tyne Hospitals NHS Foundation Trust** A De Soyza (PI), R Agbeko, K Baker, A Barr, E Cameron, Q Campbell Hewson, C Duncan, M Emonts, A Fenn, S Francis, J Glover Bengtsson, A Greenhalgh, A Hanrath, K Houghton, D Jerry, G Jones, S Kelly, N Lane, J Gray MacFarlane, P McAlinden, I J McCullagh, S McDonald, O Mohammed, R Obukofe, J Parker, A Patience, B Payne, D Price, Z Razvi, S Robson, A Sanchez, E Stephenson, R Welton, S West, E Wong, F Yelnoorkar.

**Milton Keynes University Hospital NHS Foundation Trust** R Stewart (PI), S Bowman (Co-PI), A Chakraborty (Co-PI), L How (Co-PI), D Mital (Co-PI), L Anguvaa, J Bae, G Bega, S Bosompem, E Clare, A Dooley, S Fox, J Mead, S Mehdi, L Mew, L Moran, E Mwaura, M Nathvani, A Oakley, A Rose, A Sanaullah, D Scaletta, S Shah, L Siamia, J Smith, O Spring, S Velankar, F Williams, L Wren, F Wright.

**Lancashire Teaching Hospitals NHS Foundation Trust** S Laha (PI), A Ashfaq, J Beishon (Associate PI), A Bellis, D R Cameron, M Chiu, W Choon Kon Yune, M Deeley, W Flesher, K Gandhi, S Gudur, R Gupta, A Huckle, G Long, A McCarrick, J Mills, P Mulgrew, S Nawaz, J Nixon, O Parikh, S Punnilath Abdulsamad, D Rengan, E Seager, S Sherridan, B So, R Sonia, T Southworth, K Spinks, A Timoroksa, B Vernon, A Williams, K Williams, H Wu.

**Swansea Bay University Local Health Board** B Healy (PI), S Bareford, I Blyth, A Bone, E Brinkworth, R Chudleigh, Y Ellis, D Evans, S Georges, S Green, R Harford, J Harris, A Holborow, C Johnston, P Jones, M Krishnan, N Leopold, F Morris, A Mughal, C Murphy, L O'Connnell, E Pratt, T Rees, S Richards, M Ryan, G Saleeb, J Watts, M Williams.

**Imperial College Healthcare NHS Trust** G Cooke (PI), E O Adewunmi, Z Al-Saadi, R Ashworth, J Barnacle, A Daunt, L Evison, S Fernandez Lopez, C Gale, M Gibani, A Jimenez Gil, B Jones, J Labao, N Madeja, S Mashate, H McLachlan, A Perry, R Thomas, E Whittaker, C Wignall, P Wilding, L Young, C Yu.

**James Paget University Hospitals NHS Foundation Trust** J Patrick (PI), B Burton (PI), A Ayers, R Brooks, J Chapman, V Choudhary, S Cotgrove, G Darylile, L Felton, D Griffiths, C Hacon, H Hall, W Harrison, P Hassell, A Hearn, F I Sait (Associate PI), K Mackintosh, J North, S Parslow-Williams, H Sutherland, M Whelband (Associate PI), CB Whitehouse, E Wilhelmsen, J S Y Wong, J Woods.

**Southend University Hospital NHS Foundation Trust** G Koduri (PI), S Gokaraju (Co-PI), F Hayes (Co-PI), V Warrier Vijayaraghavan Nalini (Co-PI), S Badhrinarayanan, V Gupta, P Harman, D Qureshi, M S Rabbani (Associate PI).

**North Bristol NHS Trust** N Maskell (PI), S Barratt (Co-PI), J Dodd (Co-PI), H Adamali, M Alvarez, D Arnold, S Bevins, R Bhatnagar, A Bibby, L Bradshaw, C Burden, A Cashell, H Cheshire, S Clarke, A Clive, P Creber, A Dipper, K Farmer, G Hamilton, D Higbee, A Jenkins, L Jennings, A Jeyabalan, C Kilby, H Lee, N Maskell, H McNally, A Milne, E Moran, A Morley, E Perry, N Rippon, V Sandrey, K Smith, L Solomon, L Stadon, J Townley, R Wach, D Warbrick, C Watkins, H Welch.

**Hampshire Hospitals NHS Foundation Trust** R Partridge (PI), A Goldsmith (Co-PI), Y Abed El Khaleq, AM Arias, E Bevan, J Conyngham, E Cox, E Defever, D Griffin, A Heath, B King, E Levell, X Liu, J Martin, A Nejad, F Stourton, R Thomas, D Trodd, B White, G Whitlingum, L Winckworth , C Wrey Brown, S Zagalo.

**Doncaster and Bassetlaw Teaching Hospitals NHS Foundation Trust** C-H Wong (PI), A Adeni, J Allen, S Allen, A Bassaly, M Beaumont, P Cawley, R Chadwick, R Codling, F Dunning, A Ermenyi, D Grabowska, D Graham, N Hammoud, G Herdman, M Highcock, S Hussain, N Khota, G Kirkman, C Knapp, M Kyi, A Mandal, J Maskill, V Maxwell, S McGonagle, S Mukhtar, A Nasimudeen, A Natarajan, D Pryor, D Sagar, N Saqib, P Shannon, Y Syed, D Trushell-Pottinger, L Warren, N Wilkinson, T Wilson.

**Northern Lincolnshire and Goole NHS Foundation Trust** A Mitra (PI), M N Akhtar, Hassan Al-Moasseb, S Amamou, T Behan, S Biuk, M Brazil, M Brocken, C Burnett, C Chatha, M Cheeseman, L-J Cottam, T Cruz Cervera, O Davies, K Dent, C Dyball, K Edwards, R Elmahdi, Q Farah, S Farooq, S Gooseman, J Hargreaves, MA Haroon, J Hatton, E Heeney, J Hill, E Horsley, R Hossain, D Hutchinson, J Hyde-Wyatt, M Iqbal, N James, S Khalil, M Madhusudhana, A Marriott, M T Masood, G McTaggart, K Mellows, R Miller, U Nasir, M Newton, GCE Ngui, S Pearson, C Pendlebury, R Pollard, N Pothina, D Potoczna, SD Raha, A Rehan, SAS Rizvi, A Saffy, K Shams, C Shaw, A Shirgaonkar, S Spencer, R Stead, R Sundhar, D Taylor, E Thein, L Warnock, KY Wong.

**NHS Greater Glasgow and Clyde: Royal Alexandra Hospital** A Corfield (PI), C Clark, P Clark, S Drysdale, M Hair, M Heydtmann, E Hughes, L Imam-Gutierrez, I Keith, D McGlynn, G Ray, N Rodden, K Rooney, N Thomson.

**Mid Yorkshire Hospitals NHS Trust** A Rose (PI), J Ashcroft (Co-I), P Blaxill (Co-I), S Bond (Co-I), A Dwarakanath (Co-I), C Hettiarachchi (Co-I), B Sloan (Co-I), S Taylor (Co-I), M Thirumaran (Co-I), R Beckitt, S Buckley, G Castle, E Clayton, N De Vere, J Ellam, D Gomersall, S Gordon, C Hutsby, R Kousar, K Lindley, S Oddy, A Poole (Associate PI), L Slater, B Taylor.

**Bedford Hospital NHS Trust** E Thomas (PI), D Bagmane (Co-PI), B Jallow (Co-PI), I Nadeem (Co-PI), M Negmeldin (Co-PI), A Vaidya (Co-PI), A Amjad, A Anthony-Pillai, I Armata, R Arora, R Bhanot, P Chrysostomou, F De Santana Miranda, C Donaldson, S Farnworth, N Fatimah, L Grosu, A Haddad, M Hannun, M Hikmat, B Jallow, U F Khatana, I Koopmans, E Lister, R Lorusso, N Nathaniel, C S Ong, K Pandya, M Penacerrada, Q Quratulain, S Rahama, L Salih, W T Tan, S Trussell, A Vaidya, J Valentine, F Wang, R Wulandari.

**The Dudley Group NHS Foundation Trust** H Ashby (PI), P Amy, S Ashman-Flavell, S de Silva, J Dean, N Fisher, E Forsey, J Frost, S Jenkins, A John, D Kaur, A Lubina Solomon, S Mahadevan-Bava, T Mahendiran, M O'Toole, S Pinches, D Rattehalli, U Sinha, M Subramanian, J Vamvakopoulos, S Waidyanatha.

**Ashford and St Peter's Hospitals NHS Foundation Trust** C Russo (PI), M Croft, V Frost, M Gavrila, K Gibson, A Glennon, C Gray, N Holland, J Law, R Pereira, P Reynolds, H Tarft, J Thomas, L Walding, A Williams.

**St George's University Hospitals NHS Foundation Trust** T Bicanic (PI), T Harrison (Co-PI), Y Aceampong, A Adebiyi, M Ali, D Baramova, S Drysdale, J Hayat, A Janmohamed, A Khalil, A Lisboa, A Rana, N Said, T Samakovna, A Seward, O Skelton, K Spears, A Sturdy, V Tavoukjian, S Tinashe.

**Shrewsbury and Telford Hospital NHS Trust** J Moon (PI), R Baldwin-Jones, N Biswas, A Bowes, H Button, M Carnhan, E Crawford, S Deshpande, D Donaldson, C Fenton, S Hester, Y Hussain, M Ibrahim, J Jones, S Jose, H Millward, N Motherwell, M Rees, N Schunke, A Stephens, J Stickley, M Tadros, H Tivenan.

**University Hospitals Plymouth NHS Trust** D Lewis (PI), D Affleck, O Anichtchik, K Bennett, J Corcoran, M Cramp, H Davies, J Day, M Dobranszky Oroian, E Freeman, L Madziva, P Moodley (Associate PI), C Morton, M Mwadeyi, H Notman, C Orr, A Patrick, N T Phoo (Associate PI), L Pritchard, G Selby, J Shawe, H Tan.

**The Rotherham NHS Foundation Trust** A Hormis (PI), C Brown, D Collier, C Dixon, J Field, J Ingham, S Poku, S Sampath, R Walker, L Zeidan.

**NHS Lothian: Western General Hospital** O Koch (PI), A Abu-Arafeh, E Allen, C Balmforth, A Barnett-Vanes, R Baruah, S Blackley, G Clark, S Clifford, A Clarke, M Curtin, M Evans, C Ferguson, S Ferguson, N Fethers, V Francois, N Freeman, E Gaughan, E Godden, R Harrison, B Hastings, S Htwe, A J W Kwek, O Lloyd, O Lloyd, C Mackintosh, A MacRaild, W Mahmood, E Mahony, J McCrae, E Moatt, S Morris, C Mutch, K Nunn, D O'Shea, I Page, M Perry, J Rhodes, N Rodgers, A Shepherd, R Sutherland, A Tasiou, A Tufail, D Waters, T Wilkinson, R Woodfield (Associate PI), J Wubetu.

**Chelsea and Westminster Hospital NHS Foundation Trust** P Shah (PI), B Mann (Co-PI), K Alatzoglou, K Alizaedeh, M Al-Obaidi, A Barker, C Bautista, M Boffito, M Bourke, R Bull, C Caneja, J Carungcong, P Costa, E Dwyer, C Fernandez, J Girling, E Hamlyn, A Holyome, L Horsford, N Hynes, M Johnson, U Kirwan, C Lloyd, S Maheswaran, M Martineau, M Nelson, T Ngan, K Nundlall, T Peters, A Sayan, A Schoolmeesters, M Svensson, A Tana, M Thankachen, A Thayanandan, E Vainieri,C Winpenny, O Zibdeh.

**St Helens and Knowsley Teaching Hospitals NHS Trust** G Barton (PI), N Collins, S Dealing, R Garr, S Greer, N Hornby, J Keating, S Mayor, A McCairn, S Rao, K Shuker, A Tridente.

**Tameside and Glossop Integrated Care NHS Foundation Trust** B Ryan (PI), A Abraheem, C Afnan, B Ahmed, O Ahmed, M Anim-Somuah, A Armitage, P Arora, M Beecroft, T Bull, Al-Tahoor Butt, J Fallon, J Foster, I Foulds, N Garlick, H Ghanayem, S Gulati, R Hafiz-Ur-Rehman, M Hamie, A Hewetson, B Ho, V Horsham, W Hughes, W Hulse, A Humphries, M Hussain, N Johal, E Jude, M Kelly, A Kendall-Smith, M Khan, R Law, J Majumdar, J McCormick, O Mercer, T Mirza, B Obale, P Potla, S Pudi, K Qureshi, M Rafique, R Rana, R Roberts, J Roddy, C Rolls, M Sammut, H Savill, M Saxton, V Turner, A Tyzack.

**Royal Devon and Exeter NHS Foundation Trust** M Masoli (PI), H Bakere, A Bowring, T G Burden, P Czylok, L Dobson, A Forrest, E Goodwin, H Gower, A Hall, L Knowles, H Mabb, A Mackey, V Mariano, E McEvoy, L Mckie, P Mitchelmore, L Morgan, R Oram, N Osborne, S Patten, I Seaton, R Sheridan, D Sykes, J Tipper, S Whiteley, S Wilkins, N Withers, K Zaki.

**Whittington Health NHS Trust** C Parmar (PI), W W Ang, H J Bateman, D Burrage, M Christy, P Dlouhy, J Flor, R Fromson, K Gilbert, F Green, L Howard, I Jenkin, M Kousteni, N Kulkarni, A Kyei-Mensah, I Lim, S Lock, L Ma, J Merritt, S Mindel, S Myers, L Parker, B Pattenden, A Pender, A Pratley, N Prevatt, A Reid, S Rudrakumar, J Sabale, S Sharma, P Sharratt, K Simpson, S Taylor, L Veys, N Wolff, A Zuriaga-Alvaro.

**NHS Greater Glasgow and Clyde: Queen Elizabeth University Hospital** M Sim (Co-PI), K G Blyth (Co-PI), L Jawaheer (Co-PI), L Pollock (Co-PI), S Wishart (Co-PI), M McGettrick (Sub-I), J Rollo (Sub-I), N Baxter, J Ferguson, K Ferguson, Rosemary Hague, S Henderson, L Kelly, S Kennedy-Hay, A Kidd, M A Ledingham, F Lowe, M Lowe, G McCreath, R McDougall, M McFadden, N McGlinchey, L McKay, B McLaren, C McParland, J McTaggart, J Millar, A Phillips, L Rooney, H Stubbs, M Wilson.

**Leeds Teaching Hospitals NHS Trust** J Minton (PI), S Ahmed, A Ashworth, J Bailey, M Baum, L Bonney, J Calderwood, C Coupland, M Crow, J Furness, S Hemphill, K Johnson, A Jones, M Kacar, K Khokhar, P Lewthwaite, G Lockley-Ault, F McGill, D Mistry, J Murira, Z Mustufvi, S O'Riordan, K Robinson, R Saman,J Stahle, S Straw, A Westwood, N Window.

**Warrington and Halton Teaching Hospitals NHS Foundation Trust** M Moonan (PI), M Murthy (PI), R Arya, R (P-C) Chan, L Connell, L Ditchfield, N Marriott, H Prady, L Roughley, H Whittle.

**Norfolk and Norwich University Hospitals NHS Foundation Trust** E Mishra (PI) C Atkins (Co-PI), KS Myint (Co-PI), J Nortje (Co-PI), D Archer, M Cambell-Kelly, E Chiang, P Clarke, L Coke, M Cornwell, M Del Forno, H Gorrick, A Haestier, M Harmer, L Harris, L Hudig, K Jethwa, L Jones, A Kamath, J Kennedy, J Keshet-Price, E Kolokouri, V Licence, E Lowe, E Malone, G Marayan, P Moondi, M-A Morris, K S Myint, K Shonit Nagumantry, W J Ng, J Nortje, G Randell, A F Sanz-Cepero, D Watts.

**Western Sussex Hospitals NHS Foundation Trust** L Hodgson (PI), M Margarson (Co-PI), L Albon, K Amin, M Bailey, Y Baird, I Balagosa, A Brereton, S Bullard, A Butler, V Cannons, P Carr, C Chandler, G Chow, V Dandavate, R Duckitt, A Elkhawad, S Floyd, L Folkes, K Forcer, H Fox, S Funnell, N Gent, A Ghazanfar, J Gilbert, R Gomez-Marcos, N Hedger, K Hedges, D Helm, A Hetreed, G Hobden, H Htet, D Hunt, P Jane, D Jennings, A Kanish, R Khan, S Kimber, K King, Z Krejcarova, T Leckie, M Linney, L Lipskis, C Long, J Margalef, T Martindale, A Matthew, M McCarthy, P McGlone, E Meadows, S Moore, T Moore, S Murphy, M Nelmes, L Nguyen, R Njafuh, L Norman, N Numbere, M Parson, E Pineles, M Purcell, L Ramsawak, C Ranns, D Raynard, D Reynish, L Riddles, C Ridley, E Robinson, J Russell, T Shafi, Tomos G Shaw, S Sinha, Jessica Smith, S Stone, P Tate, B Thillainathan, Y Thirlwall, E Vamvakiti, R Venn, J Villiers, N White, J Wileman, E Yates.

**West Suffolk NHS Foundation Trust** M Moody (PI), S Barkha (Sub-PI), S Bhagat, H Cockerill, J Godden, J Kellett, T Murray, P Oats, A Saraswatula, A Williams, L Wood.

**Cwm Taf Morgannwg University LHB** C Lynch (PI), B Deacon, S Eccles, A Gaurav, B Gibson, M Hibbert, C Lai, L Margarit, D S Nair, S Owen, J Pugh, L Roche, S Sathe, J Singh, D Tetla, C Woodford.

**South Eastern HSC Trust** D Alderdice (PI), J Courtney (Co-I), J Elder (Co-I), D Hart (Co-I), K Henry (Co-I), R Hewitt (Co-I), A Kerr (Co-I), J McKeever (Co-I), C O'Gorman (Co-I), S Rowan (Co-I), T Trinick (Co-I), B Valecka (Co-I), P Yew (Co-I), V Adell, J Baker, A Campbell, J Foreman, P Gillen, S Graham, S Hagan, L Hammond, J MacIntyre, L Moore, S Regan, A Smith, G Young.

**Kingston Hospital NHS Foundation Trust** S Mahendran (PI), A Joseph (Co-PI), A Baggaley, G Bambridge, F Bazari, P Beak, L Boustred, T Carlin, A Conroy, J Crooks, B Crowe, E Donnelly, A Edwards, S El-Sayeh, A Feben, J Fox, R Gisby, J Haugh, R Heath, R Herdman-Grant, E Jackson, D Jajbhay, O James, S Jones, A Joseph, T Leahy, S Luck, M Madden, J May, G McKnight, L Mumelj, G Natarajan, D Newman, A Nicholson, T O'Brien, J Odone, R Patterson, J Poxon, A Ratnakumar, R Rodriguez-Belmonte, T Sanderson, S Sharma, R

Simms, D Sivakumaran, A Sothinathan, A Swain, S Swinhoe, M Taylor, M Trowsdale Stannard, J Vance-Daniel, H Warren-Miell, T Woodhead, D Zheng.

**Worcestershire Acute Hospitals NHS Trust** C Hooper (PI), K Austin, T Dawson, A Durie, C Hillman-Cooper, M Ling, S Stringer, H Tranter, J Tyler, P Watson, H Wood.

**Poole Hospital NHS Foundation Trust** H Reschreiter (PI), S August, C Barclay, S Blunden, S Bokhandi, J Camsooksai, S Chessell, C Colvin, J Dube, S Grigsby, C Humphrey, S Jenkins, S Patch, A Shah, M Tighe, L Vinayakarao, B Wadams, E Woodward, M Woolcock.

**Walsall Healthcare NHS Trust** A Garg (PI), K Shalan (PI), S Al-Hity, A Alina, B Allman, J Bearpark, S Bibi, L Botfield, A Bowes, L Boyd, W Campbell, Z Chandler, J Collins, G K Dhaliwal, L Dwarakanath, A Farg, A Foot, A Gondal, T Gupta, T Jemima, P A Joseph, O Khan, J Khatri, R Krishnamurthy, H Mahmoud, R Marsh, R Mason, M Matonhodze, S Misra, J Muhammad, A Naqvi, C Newson, E Nicholls, S Ann Nortcliffe, S W Odelberg, M Phipps, A J Plant, K Punia, H Qureshi, G Rajmohan, P Ranga, A Raymond-White, N Richardson, L Rogers, A Sheikh, A Srirajamadhuveeti, N Sunni, R Turel, E Virgilio, F Wyn-Griffiths.

**Southport and Ormskirk Hospital NHS Trust** S Pintus (PI), A Ahmed (Co-I), A Nune (Co-I), S Abdelbadiee, L Afari, L Aitchson, A Ali, S Asam, N Babajan, B Bainton, L Bishop, K Choudhary, A Christie, R Cox, M Diwan, W Gaba, H Gibson, Z Haslam, A Hassan, C Hutchcroft, M Jackson, A Liaretidou, M Mahmood, E McDonald, A Morris, M Morrison, N Ndoumbe, S O'Brien, Niranjani Ramachandran, S Rehman, N Shami, L Smith, Vaidyanathan Subramanian, L Undrell, K Wahdati, M Wood.

**The Royal Wolverhampton NHS Trust** S Gopal (PI), R Barlow, C H Cheong, D Churchill, K Davies, M Green, N Harris, A Kumar, S Metherell, S Milgate, L Radford, J Rogers, A Smallwood, L Wild.

**Western HSC Trust** M Kelly (PI), D Concannon, K Ferguson, D McClintock, V Mortland, N Smyth, J Wieboldt.

**Mid Essex Hospital Services NHS Trust** A Hughes (PI), J Radhakrishnan (Co-PI), R Arnold, T Camburn, C Catley, E Dawson, C Fox, N Fox, H Gerrish, S Gibson, H Guth, F McNeela, A Rao, S Reid, B Singizi, S Smolen, S Williams, L Willsher, J Wootton.

**The Queen Elizabeth Hospital, King's Lynn, NHS Foundation Trust** M Blunt (PI), S M Abubacker, J Ali, K Beaumont, K Bishop, H Bloxham, P Chan, Z Coton, H Curgenven, M Elsaadany, T Fuller, M Iqbal, M Israa, S Jeddi, S A Kamerkar, E T Lim, E Nadar, K Naguleswaran, O Poluyi, H Rangarajan, G Rewitzky, S Ruff, S Shedwell, A Velusamy, H Webb.

**Brighton and Sussex University Hospitals NHS Trust** M Llewelyn (PI), H Brown, E Barbon, G Bassett, L Bennett, A Bexley, P Bhat, Z Cipinova, A V Elkins, Lynn Evans, J Gaylard, Z He, S Jujjavarapu, C Laycock, D Mullan, J Newman, C Richardson, V Sellick, D Skinner, M Smith, D Yusef.

**Royal Papworth Hospital NHS Foundation Trust** R C Rintoul (PI), F Bottrill, A Butchart, K Dorey, R Druyeh, M Earwaker, C East, S Fielding, A Fofana, C Freeman, C Galloway, L Garner, A Gladwell, V Hughes, R Hussey, N Jones, S Mepham, A Michael, H Munday, K Paques, H Parfrey, C Pathvardhan, G J Polwarth, A Rubino, S Webb, S Woods, K Woodall, A Vuylsteke, J Zamikula.

**NHS Grampian: Aberdeen Royal Infirmary** J Cooper (PI), V Bateman, M Black, R Brittain-Long, K Colville, D Counter, S Devkota, P Dospinescu, J Irvine, C Kaye, L Kane (Associate

PI),A Khan, R Laing, A Mackenzie, M J MacLeod, G Malik, J McLay, D Miller, K Norris, Y W Pang, R Soiza, V Taylor, I Tonna.

**Gloucestershire Hospitals NHS Foundation Trust** C Sharp (PI), F Ahmed, O Barker, O Bintcliffe, P Brown, R Bulbulia, J Collinson, T Cope, A Creamer, C Davies, W Doherty, M Fredlund, J Glass, S Harrington, A Hill, H Iftikhar, M James, S Jones, C Lim, S Message, H Munby, J Ord, J Patel, R Peek, T Pickett, A Simpson, M Slade, C Thompson, H Uru, D Ward, R Woolf.

**University Hospital Southampton NHS Foundation Trust** S Fletcher (PI), J Bigg, K Cathie, S Chabane, M Coleman, S N Faust, M Felongco, J Forbes, T Francis-Bacon, E Holliday, M Johnson, CE Jones, T Jones, S Michael, M Nelson, M Petrova, L Presland, A Procter, N Rayner, R Samuel, T Sass, M Shaji, C S Moniz, T Thomas, S Triggs, C Watkins, S Wellstead, H Wheeler.

**Dorset County Hospital NHS Foundation Trust** J Chambers (PI), J Birch, L Bough, J Colton, J Graves, S Horton, J Rees, R Thomas, W Verling, S Williams, P Williams, B Winter-Goodwin, S Wiseman, D Wixted.

**NHS Lothian: St John's Hospital** S Lynch (PI), R Anderson, S Begg, M Colmar, C Cheyne, R Frake, A Gatenby, C Geddie, F Guarino, C Hurley, C Kuronen-Stewart, A MacRaild, M Mancuso-Marcello, G McAlpine, A Megan, T North, M Odam, OK Otite, L Primrose, L Rooney, Anne Saunderson, A Stevenson, S Stock, A Wakefield, E Walsh, J Wraight.

**George Eliot Hospital NHS Trust** S George (PI), S Bukhari, K Ellis, V Gulia, J Gunn, J Heys, E Hoverd, T Kannan, R Musanhu, N Navaneetham, D Suter.

**NHS Fife** D Dhasmana (PI), F Adam, K Aniruddhan, J Boyd, N Bulteel, P Cochrane, K Gray, L Hogg, S Iwanikiw, M Macmahon, A Morrow, J Penman, J Pickles, A Scullion, H Sheridan, D Sloan, C Stewart, M Topping.

**Croydon Health Services NHS Trust** T Castiello (PI), J Adabie-Ankrah, G Adkins, B Ajay, S Ashok, A Dean, S Dillane, F Fedel, V Florence, D Griffiths, I Griffiths, J Hajnik, J Hetherington, C Jones, A Latheef, S Lee, J McCammon, S Patel, A Raghunathan, P Shah, J Talbot-Ponsonby, G Tsinaslanidis, G Upson.

**NHS Ayrshire and Arran: University Hospital Crosshouse** A Clark (PI), T Adams, S Allen, K Bain, A Bal, C Burns, D Callaghan, N Connell, V Dey, F Elliott, K Gibson, D Gilmour, H Hartung, M Henry, G Houston, L McNeil, A Murphy, S Smith, S Walton, D Wilkin, M Wilson, S Wood.

**NHS Lanarkshire: University Hospital Monklands** M Patel (PI), C McGoldrick (Co-PI), C Beith, D Cairney, L Ferguson, L Glass, P Grant, S MacFadyen, A McAlpine, M McLaughlin, S Rundell, C Sykes, M Taylor, B Welsh.

**University College London Hospitals NHS Foundation Trust** H Esmail (PI), R S Heyderman (Co-PI), D Moore (Co-PI), F Beynon, P N Bodalia, X H Chan, Z Chaudhry, CY Chung, D Crilly, J Cohen, S Eisen, N Fard, J Gahir, L Germain, J Glanville, V Johnston, E Kilich, N Lack, M M Morrillas, J Millard, N Platt, S Roy, I Skorupinska, M Skorupinska, J Spillane.

**Maidstone and Tunbridge Wells NHS Trust** M Szeto (PI), K Cox (Co-PI), A Abbott, S Anandappa, B Babiker, C Bailey, M Barbosa, G Chamberlain, D Datta, M Davey, R Gowda, A Gupta, R Hammond-Hall, E Harlock, C Hart, A Henderson, S Y Husaini, E Hutchinson, A

Keough, S Kumar, T-K Loke, S Matthew, R Nemane, I Pamphlett, C Pegg, P Rajaopalan, A Richards, S Siddavaram, H Slater, G Sluga, O Solademi, P Tsang, AWaller.

**Northern HSC Trust** P Minnis (PI), J Burns, L Davidson, A Fryatt, J Gallagher, M A Kawser, L Kingsmore, C McGoldrick, M McMaster, M Nugdallah.

**Royal United Hospitals Bath NHS Foundation Trust** J Suntharalingam (PI), J Avis, C Broughton, S Burnard, C Demetriou, J Fiquet, J Ford, O Griffiths, R Hamlin, T Hartley, S Jones, J Macaro, R M Ross, C Marchand, V Masani, S Mitchard, A Palmer, L Ramos, M Rich, J Rossdale, S Sturney, G Towersey, J Tyler, K White.

**NHS Tayside: Ninewells Hospital** J Chalmers (PI), H Abo-Leyah, C Almadenboyle, C Deas, H Loftus, A Nicoll, L Smith, A Strachan, J Taylor, C Tee.

**Stockport NHS Foundation Trust** R Stanciu (PI), M Afridi, M A Dakhola, S Bennett, L Brown, C Cooper, A Davison, D Eleanor, J Farthing, A Ferrera, S Ghandi, L Gomez, P Haywood, C Heal, H Jackson, J Johnston, A Lloyd, S McCaughey, R Owen, A Pemberton, F Rahim, H Robinson, N Sadiq, R Samlal, V Subramanian, D Suresh, H Wieringa, I Wright.

**Wye Valley NHS Trust** I DuRand (PI), P Ryan (Deputy PI), J Al-Fori, J Birch, N Bray, A Carrasco, M Cohn, E Collins, S Cooper, A Davies, M Evans, H Gashau, K Hammerton, A Hassan, S Maryosh, S Meyrick, B Mwale, L Myslivecek, A Salam, C Seagrave, F Suliman, S Turner, J Woolley.

**NHS Lanarkshire: University Hospital Wishaw** M Patel (PI), K Black, R Boyle, S Clements, J Fleming, L Glass, L Hamilton, E Jarvie, C MacDonald, N Moody (Associate PI), D Vigni, B Welsh, P Wu.

**Betsi Cadwaladr LHB: Ysbyty Gwynedd** C Subbe (PI), N Boyle, C Butterworth, M Joishy, E Knights, G Rieck, W Scrase, A Thomas, C Thorpe.

**The Princess Alexandra Hospital NHS Trust** U Ekeowa (PI), Q Shah (Co-PI), S Sakthi (Co-Pi), M Anwar, G Arunachalam, B Badal, K Bamunuarachchi, G Cook, A Daniel, J Finn, C Freer, A Gani, E Haworth, E Holmes, L Hughes, K Ixer, G Lucas, C Muir, S Naik, R Ragatha, P Russell, R Saha, L Sandhu, E Shpuza, N Staines, S Waring, L Wee, F Weidi, T White.

**Cambridge University Hospitals NHS Foundation Trust** M Knolle (PI), E Gkrania-Klotsas (Co-PI), P Bailey, K Beardsal, R Bousefield, K Bunclark, S Burge, J Chung, T Dymond, A Edwards, M Fisk, K Gajewska-Knapik, J Galloway, C Harris, A Jha, R Kumar, K Leonard, C Ma, A Martinelli, Z McIntyre, N Pathan, S Rossi, J Sahota, G Stewart, A Sutton-Cole, M E Torok, M Toshner, C Yong.

**East Lancashire Hospitals NHS Trust** S Chukkambotla (PI), Ana Batista, H Collier, S Duberley, W Goddard, B Hammond**,** A Konstantinidis**,** K Marsden, Mulla, A Newby, J Nugent, D Rusk, C Spalding, A Sur, D Sutton, J Umeadi.

**NHS Forth Valley: Forth Valley Royal Hospital** M Spears (PI), A Baggott, G Clark, J Donnachie, S Huda, G Jayasekera, I Macpherson, M Maycock, S McKenna, J McMinn, D Morrison, A Pearson, L Prentice, C Rafique, D Salutous, L Symon, A Todd, P Turner.

**Harrogate and District NHS Foundation Trust** A Kant (PI), C Taylor (Co-PI), A Amin, C Bennett, O Cohen, A Daly, S-J Foxton, E Lau, C Morgan, N Singh, A Williamson, M Tripouki, L Wills.

**East Sussex Healthcare NHS Trust** A Marshall (PI), S Ahmed (Associate PI), S Blankley, H Brooke-Ball, T Christopherson, M Clark, T De Freitas, E De Sausmarez, A Ekunola, D Hemsley, J Highgate, O Kankam, A Lowe, S Merritt, Y N S Mohammed, T Morley, A Newby, S Panthakalam, S Qutab, R Reddy, N Roberts, M J Sinclair, K Subba, S Tieger, A Trimmings, R Venn, F Willson, T T Win, M Yakubi, A Zubir.

**NHS Dumfries and Galloway: Dumfries & Galloway Royal Infirmary** D Williams (PI), M McMahon (Co-PI), P Cannon (Associate PI), J Duignan, C Jardine, A Mitra, P Neill, S Svirpliene, S Wisdom.

**Homerton University Hospital NHS Foundation Trust** K Woods (PI), A Claxton (Co-PI), Y Akinfenwa, N Aladangady, Ayesha Begum, H Bouattia, R Brady, A Chiapparino, R Corser, R Frowd, H Furreed, C Holbrook, S Jain, M Jagpal, S Jain, J Kaur, C Mitchell-Inwang, R Mullett, T Tanqueray, L Terry, E Timlick,

**Belfast HSC Trust** D Downey (PI), A Blythe, S Carr, D Comer, D Dawson, R Ingham, J Kidney, J Leggett, A Redfern-Walsh.

**Betsi Cadwaladr LHB: Glan Clwyd Hospital** D Menzies (PI), A Abou-Haggar, S Ambalavanan, K Darlington, F Davies, G Davis, I Davis, J Easton, T Grenier, S Horrocks, M Joishy, R Lean, J Lewis, C Mackay, R Poyner, R Pugh, X Qui, S Rees, N Sengupta, H Williams.

**Lewisham and Greenwich NHS Trust** S Kegg (PI), A Aghababaie, H Azzoug, E Bates, M Chakravorty, K Chan, P G L Coakley, F Chukwunonyerem, E Gardiner, A Hastings, D Jegede, J Juhl, S Khatun, M Magriplis, C Milliken, J Muglu, D Mukimbiri, M Nadheem, T Nair, M Nyirenda, T Oconnor, T Ogbara, R Olaiya, C Onyeagor, V Palaniappan, A Pieris, S Pilgrim, C Saad, N Sengreen, T Simpson, A Taylor, K Wesseldine, M Woodman, E Woolley.

**The Hillingdon Hospitals NHS Foundation Trust** S Kon (PI), T Bate, L Camrasa, A Danga, S Dubrey, J Ganapathi, B Haselden, M Holden, E Kam, J Korolewicz, S-J Lam, G Landers, P Law, N Mahabir, M K Majumder, N Malhan, M Nasseri, T Nishiyama, P Palanivelu, J Potter, S Ramraj, A Seckington, T Sugai, A Trivedi, S I Vandeyoon, W Varney, D Wahab.

**Royal Cornwall Hospitals NHS Trust** D Browne (PI), Z Berry, H Chenoweth, A Collinson, F Hammonds, L Jones, E Laity, T Nisbett, R Sargent, K Watkins, L Welch.

**Taunton and Somerset NHS Foundation Trust** J Pepperell (PI), J Ashcroft, C Branfield, S Crouch, I Cruickshank, J Foot, C Lanaghan, D Lewis, C Lorimer, H Mills, G Modgrill, A Moss, M Nixon, S Northover, K O'Brien, K Roberts, J Rogers, C Thompson, N Thorne, R Wallbutton, E Zebracki.

**Guy's and St Thomas' NHS Foundation Trust** H Winslow (PI), L Brace, K Brooks, L Chappell, J Cordle, M Flanagan, B Hamilton, D Hydes, J Kenny, A Lewin, L Martinez, G Nishku, C Singh, E Wayman, C Williamson, H Winslow, C Yearwood Martin.

**Yeovil District Hospital NHS Foundation Trust** A Broadley (PI), S Board, A Daxter, I Doig, A Getachew, L Howard, R Jonnalagadda, A Kubisz-Pudelko, A Lewis, K Mansi, R Mason, A S Melinte, B Mulhearn, J Reid, A Shah, R Smith, D Wood.

**Airedale NHS Foundation Trust** T Gregory (PI), M Babirecki, H Bates, E Docks, E Dooks, F Farquhar, B Hairsine, S Nallapeta, S Packham.

**Salford Royal NHS Foundation Trust** P Dark (PI), C Bethan, B Blackledge, N D Bakerly, L Catlow, B Charles, J Harris, A Harvey, K Knowles, S Lee, T Marsden, L McMorrow, J Perez, M Poulaka, R Sukla, M Taylor, V Thomas, S Warran.

**Torbay and South Devon NHS Foundation Trust** T Clarke (PI), I Akinpelu, S Atkins, J Blackler, J Clouston, G Curnow, Foulds, J Graham, C Grondin, S Howlett, C Huggins, L Kyle (Associate PI), S Martin, P Mercer, W O'Rourke, A Redome, J Redome, R Tozer, J Turvey.

**Royal Surrey County Hospital NHS Foundation Trust** K McCullough (PI), H-J Abu, C Beazley, H Blackman, P Carvelli, P Chaturvedi, B Creagh-Brown, J De Vos, S Donlon, C Everden, J Fisher, H Gale, E Gallagher, D Greene, O Hanci, E Harrod, N Jeffreys, J Jones, R Jordache, R Mehra, N Michalak, O Mohamed, S Mtuwa, K Penhaligon, V Pristopan, M Sanju, E Smith, S Stone, S Tluk.

**Barnsley Hospital NHS Foundation Trust** K Inweregbu (PI), M Cunningham, A Daniels, C Green, R Gupta, L Harrison, A Hassan, S Hope, M Hussain, A Khalil, S Meghjee, A Nicholson, A Sanderson.

**Isle Of Wight NHS Trust** A R Naqvi (PI), M Pugh (PI), A Brown, R Fasina, A D Gardener (Associate PI), S Grevatt, E Jenkins, S Knight, X Liu, Magier, S A A Mukhtar, E Nicol, Emma O'Bryan, Joseph Selley, J Wilkins.

**Betsi Cadwaladr LHB: Wrexham Maelor Hospital** D Southern (PI), M Garton (Co-I), S Ahmer, G Bennett, S David, S Davies, E Heselden, M Howells, R Hughes, S Kelly, A Lloyd, Maraj, H Reddy, S Robertson, S Smuts, J Smyth, G Spencer, G Szabo, S Tomlins.

**NHS Borders: Borders General Hospital** A Scott (PI), S Alcorn, J Aldridge, J Bain, A Campbell, J Dawson, E Dearden, T Downes, A Duncan, C Evans, C Flanders, N Hafiz, L Jansen, L Knox, J Lonnen, C Murton, B Muthukrishnan, F Rodger, B Soleimani, M Tolson.

**East and North Hertfordshire NHS Trust** M Chaudhury (PI), C Cruz (Co-I), M Ebon (Co-I), N Pattison (Co-I), J Asplin, P Baker, D Banner, H Beadle, C Cruz, S Dabbagh, M Ebon, V Elliott, A Fajardo, P Ferranti, J Gilmore, S Gohil, A Hood, T Ingle, E Jenner, Z Kantor, J Kefas, C Matei, J Mathers, K Mccord, K Narula, J Newman, Y Odedina, D Palit, L Peacock, M Raithatha, S Sarai, L Ventilacion, E Vilar, R Yellon, R Zill-E-Huma.

**Northern Devon Healthcare NHS Trust** R Manhas (PI), U Akudo, K Allen, A Attiq, V Ayra, C Baldwick, F Bellis, H Black, L Brunton, M Bryce, K Causer, S Cockburn, R Crowder, Dermot Dalton, D Davies, C Ferreira-De Almeida, M Freeborn, H Goss, E Gray, I Gurung, G Hands, R Hartley, B Holbrook, N Hollister, R Horn, J Hunt, M S Jeelani, S Kyle, M Lamparski, Eleni Lekoudis, M Lewis, S Ley, L Lindenbaum, S Mole, A Moody, J Morrison, J Raza, T Reynolds, G Rousseau, B Rowlands, M Ruiz, G Sacher, C Smith, D Tharmaratnam, B Theron, A Umeh, L van Koutrik, N Vernon, C White, E Willis.

**Hywel Dda LHB: Prince Philip Hospital** S Ghosh (PI), S Coetzee, K Davies, K Lewis, L O'Brien, Z Omar, C V Williams, C Wollard.

**West Hertfordshire Hospitals NHS Trust** V Page (PI), R Vancheeswaran (Co-PI), L Norris, T Varghese, X Zhao.

**NHS Lanarkshire: University Hospital Hairmyres** M Patel (PI), F Burton (Co-PI), D Bell, R Boyle, D Cairney, K Douglas, L Glass, E Jarvie, E Lee, L Lennon, S Naidoo, B Welsh.

**NHS Highland** B Sage (PI), F Barrett, W Beadles, C J Bradley, A Cochrane, R Cooper, A Goh, S Makin, J Matheson, D McDonald, C Millar, K Monaghan, L Murray, D Patience, G Simpson.

**NHS Ayrshire and Arran: University Hospital Ayr** K Walker (PI), C Burns, D Callaghan, R Cuthbertson, K Gibson, D Gilmour, M Henry, J Locke, L McNeil, S Meehan, A Murphy, K Naismith, K Prasad, M Rodger, C Turley, S Walton, M Wilson.

**NHS Greater Glasgow and Clyde: Inverclyde Royal Hospital** M Azharuddin (PI), H Papaconstantinou (Co-PI), D Cartwright, W Gallagher, T McClay, E Murray, O Olukoya.

**Salisbury NHS Foundation Trust** M Sinha (PI), A Anthony, L Bell, S Diment, S Gray, A Hawkins, M Johns, V King, I Leadbitter, W Matimba-Mupaya, A Rand, S Salisbury, S Strong-Sheldrake, F Trim.

**South Warwickshire NHS Foundation Trust** S Tso (PI), P Parsons (Co-PI), S Bird, C Bannon, R Browne, B Campbell, S Dhariwal, G Kakoullis, F Mackie, C O'Brien, K Webb.

**Royal Brompton & Harefield NHS Foundation Trust** A Shah (PI), A Reed (Co-PI), A Aramburo, B Araba, L Banton, R Mordi, T Poonian, C Prendergast, P Rogers, N Soussi, V Teli, J Wallen.

**The Royal Marsden NHS Foundation Trust** K Tatham (PI), S Jhanji (Co-I), P Angelini, E Bancroft, E Black, A Dela Rosa, E Durie, M Hogben, I Leslie, A Okines, I Sana, S Shepherd, N Taylor, S Wong.

**East Cheshire NHS Trust** T Nagarajan (PI), M Holland, L Huhn, M A Husain, N Keenan, X Lee, L Wilkinson, K Wolffsohn.

**Birmingham Women's and Children's NHS Foundation Trust** K Morris (PI), J Groves, K Hong, D Jyothish, S Sultan.

**Great Ormond Street Hospital For Children NHS Foundation Trust** M Peters (PI), A Bamford, L Grandjean (Co-PI), E Abaleke, O Akinkugbe, H Belfield, G Jones, T McHugh, L O'Neill, S Ray, A LuisaTomas.

**Alder Hey Children's NHS Foundation Trust** D Hawcutt (PI), D Afolabi, K Allison, S McWilliam, L O'Malley, L Rad, N Rogers, P Sanderson, G Seddon, J Whitbread.

**Hywel Dda LHB: Bronglais General Hospital** M Hobrok (PI), D Asandei, S Jenkins, K Khan, R Loosley, D McKeogh, L Raisova, A Snell, H Tench, T Wareham, R Wolf-Roberts.

**Hywel Dda LHB: Withybush Hospital** J Green (PI), R Hughes, C Macphee, H Thomas.

**The Christie NHS Foundation Trust** V Kasipandian (PI), A Binns, J King, P Mahjoob-Afag, R Mary-Genetu, P Nicola, A Patel, R Shotton, D Sutinyte.

**NHS Western Isles** G Stanczuk (PI), I Garcia Deniz, A Apostolopoulos (Co-PI) S Klaczek, M Murdoch.

**The Walton Centre NHS Foundation Trust** R Davies (PI), H Arndt, A Clyne, E Hetherington.

**Sheffield Children's NHS Foundation Trust** P Avram (PI), C Kerrison (sub PI), A Bellini, F Blakemore, S Borg, K Bourne, J Bryant, C Chambers, H Chisem, J Clemens, H Cook, P Dimitri, M Dockery, M Elfadil, S Gormley, D Hawley, A Howlett, G Margabanthu, A-M McMahon, J Nolan, B O'Shea, N Roe, J Sowter, T Williams.

**Velindre NHS Trust** J Powell (PI), R Adams (Co-PI), A Jackson.

**Liverpool Women's NHS Foundation Trust** R McFarland (PI), P Corlett, C Cunningham, M Dower, S Holt, K Knowles, J McKenzie, C Morgan, E Neary, A Smith, M Turner.

**NHS Golden Jubilee National Hospital** B Shelley (PI), V Irvine, F Thompson.

**Dragon's Heart Hospital** J Coulson (PI), B Moore.

### Supplementary Methods

#### Study organization

The RECOVERY trial is an investigator-initiated, individually randomised, open-label, controlled trial to evaluate the efficacy and safety of a range of putative treatments in patients hospitalized with COVID-19. The protocol is available at www.recoverytrial.net. The trial was conducted at 176 National Health Service (NHS) hospital organizations in the United Kingdom. The trial was coordinated by a team drawn from the Clinical Trial Service Unit and the National Perinatal Epidemiology Clinical Trials Unit within the Nuffield Department of Population Health at University of Oxford, the trial sponsor. Support for local site activities was provided by the National Institute for Health Research Clinical Research Network.

Treatment supply to local sites was supported by National Health Service (NHS) England and Public Health England. Access to relevant routine health care and registry data was supported by NHS DigiTrials, the Intensive Care National Audit and Research Centre, Public Health Scotland, National Records Service of Scotland, and the Secure Anonymised Information Linkage (SAIL) at University of Swansea.

#### Protocol changes

RECOVERY is a randomised trial among patients hospitalized for COVID-19. All eligible patients receive usual standard of care in the participating hospital and are randomly allocated between no additional treatment and one of several active treatment arms. Over time, additional treatment arms have been added (see Table). As outlined in the protocol, if one or more of the active treatments was not available at the hospital or is believed, by the attending clinician, to be contraindicated (or definitely indicated) for the specific patient, then random allocation was between the remaining treatment arms.

The original and final protocol relevant to azithromycin are included in the supplementary material to this publication, together with summaries of the changes made.

**Table. Protocol changes to treatment comparisons**

| Protocol version | Date | Randomisation | Treatment arms |
|---|---|---|---|
| 1.0 | 13-Mar-2020 | Main (part A) | No additional treatment<br>Lopinavir-ritonavir[a]<br>Low-dose corticosteroid[b]<br>Nebulised Interferon-ß-1a (never activated) |
| 2.0 | 23-Mar-2020 | Main (part A) | No additional treatment<br>Lopinavir-ritonavir[a]<br>Low-dose corticosteroid[b]<br>Hydroxychloroquine |
| 3.0 | 07-Apr-2020 | Main (part A) | No additional treatment<br>Lopinavir-ritonavir[a]<br>Low-dose corticosteroid[b]<br>Hydroxychloroquine[c]<br>Azithromycin |

| Protocol version | Date | Randomisation | Treatment arms |
|---|---|---|---|
| 4.0 | 14-Apr-2020 | Main (part A)<br><br><br><br><br><br>Second[d] | No additional treatment<br>Lopinavir-ritonavir[a]<br>Low-dose corticosteroid[b]<br>Hydroxychloroquine[c]<br>Azithromycin<br><br>No additional treatment<br>Tocilizumab |
| 5.0 | 24-Apr-2020 | - | (no change – extension to children <18 years old) |
| 6.0 | 14-May-2020 | Main (part A)<br><br><br><br><br><br>Main (part B factorial)<br><br><br>Second[d] | No additional treatment<br>Lopinavir-ritonavir[a]<br>Low-dose corticosteroid[b]<br>Hydroxychloroquine[c]<br>Azithromycin<br><br>No additional treatment<br>Convalescent plasma<br><br>No additional treatment<br>Tocilizumab |
| 7.0 | 18-Jun-2020 | Main (part A)<br><br><br><br><br>Main (part B factorial)<br><br><br>Second[d] | No additional treatment<br>Lopinavir-ritonavir[a]<br>Low-dose corticosteroid[b]<br>Azithromycin<br><br>No additional treatment<br>Convalescent plasma<br><br>No additional treatment<br>Tocilizumab |
| 8.0 | 03-Jul-2020 | Main (part A)<br><br><br><br><br><br>Main (part B factorial)<br><br><br>Second[d] | No additional treatment<br>Low-dose corticosteroid[b]<br>Intravenous immunoglobulin[e]<br>High-dose corticosteroid[e]<br>Azithromycin<br><br>No additional treatment<br>Convalescent plasma<br><br>No additional treatment<br>Tocilizumab |

| **Protocol version** | **Date** | **Randomisation** | **Treatment arms** |
|---|---|---|---|
| 9.1 | 18-Sep-2020 | Main (part A) | No additional treatment<br>Low-dose corticosteroid[b]<br>Intravenous immunoglobulin[e]<br>High-dose corticosteroid[e]<br>Azithromycin |
| | | Main (part B factorial) | No additional treatment<br>Convalescent plasma<br>REGEN-COV2 |
| | | Second[d] | No additional treatment<br>Tocilizumab |
| 10.1 | 01-Nov-2020 | Main (part A) | No additional treatment<br>Low-dose corticosteroid[b]<br>Intravenous immunoglobulin[e]<br>High-dose corticosteroid[e]<br>Azithromycin[f] |
| | | Main (part B factorial) | No additional treatment<br>Convalescent plasma<br>REGEN-COV2 |
| | | Main (part C factorial) | No additional treatment<br>Aspirin |
| | | Second[d] | No additional treatment<br>Tocilizumab |
| 11.1 | 27-Nov-2020 | Main (part A) | No additional treatment<br>Low-dose corticosteroid[b]<br>Intravenous immunoglobulin[e]<br>High-dose corticosteroid[e]<br>Colchicine |
| | | Main (part B factorial) | No additional treatment<br>Convalescent plasma<br>REGEN-COV2 |
| | | Main (part C factorial) | No additional treatment<br>Aspirin |
| | | Second[d] | No additional treatment<br>Tocilizumab |

[a] enrolment ceased 29 June 2020 when the Data Monitoring Committee advised that the Chief Investigators review the unblinded data.

[b] enrolment of adults ceased 8 June 2020 as more than 2,000 patients had been recruited to the active arm

[c] enrolment ceased 5 June 2020 when the Data Monitoring Committee advised that the Chief Investigators review the unblinded data.

[d] for patients with (a) oxygen saturation <92% on air or requiring oxygen or children with significant systemic disease with persistent pyrexia; and (b) C-reactive protein ≥75 md/dL)

[e] for children only

[f] enrolment of adults ceased 27 November 2020 as more than 2,500 patients had been recruited to the active arm

#### Supplementary statistical methods

*Sample size*

As stated in the protocol, appropriate sample sizes could not be estimated when the trial was being planned at the start of the COVID-19 pandemic. As the trial progressed, the Trial Steering Committee, blinded to the results of the study treatment comparisons, formed the view that sufficient patients should be enrolled to each comparison to provide at least 90% power at two-sided P=0.01 to detect a proportional reduction in 28-day mortality of one-fifth. Thus, if 28-day mortality was 20% then a comparison of at least 2000 participants allocated to active drug and 4000 to usual care alone would suffice.

However, if the 28-day mortality was lower, then more participants would be required for equivalent statistical power. Based on review of the blinded data, the Trial Steering Committee determined that recruitment to the comparison of azithromycin should cease once at least 2500 patients had been allocated to the active drug and could be compared with at least 5000 patients allocated to the usual care alone arm. If 28-day mortality was 18% in the usual care group, a study of this size would have 90% power at two-sided P=0.01 to detect a proportional reduction of one-fifth.

*Baseline-predicted risk*

Baseline−predicted risk of 28-day mortality was estimated through the formula 100 x exp(a)/(1 + exp(a)), where a = −1·23 − 2·85 (if age <50) − 2·03 (if age 50−59) − 1·21 (if age 60−69) − 0·51 (if age 70−79) + 0·42 (if male) − 0·34 (if >7 days since symptom onset) + 0·86 (if on oxygen only at randomisation) + 2·18 (if on invasive mechanical ventilation at randomisation) − 0·01 (if history of diabetes) + 0·22 (if history of heart disease) + 0·21 (if history of chronic lung disease) + 0·50 (if history of kidney disease). These regression coefficients were derived from a multivariable logistic regression model using data from 10,702 trial participants who had complete 28-day mortality follow-up data by 22 June 2020. The regression model additionally adjusted for treatment allocation (with usual care designated the reference category) and for all possible two-way interactions between the above baseline characteristics and treatment allocation. These additional terms were ignored when calculating baseline-predicted risk, however, in order to ensure that the estimates corresponded to risk *if assigned usual care*. Patients were then subdivided into three approximately equally-sized groups (across all RECOVERY participants) on the basis of their predicted risk: <30%, ≥30% to <45%, and ≥45%.

#### Ascertainment and classification of study outcomes

Information on baseline characteristics and study outcomes was collected through a combination of electronic case report forms (see below) completed by members of the local research team at each participating hospital and linkage to National Health Service, clinical audit, and other relevant health records. Full details are provided in the RECOVERY Definition and Derivation of Baseline Characteristics and Outcomes Document which was first published online (www.recoverytrial.net) on 9 June 2020.

*Randomisation form*

The Randomisation form (shown below) was completed by trained study staff. It collected baseline information about the participant (including demographics, COVID-19 history, comorbidities and suitability for the study treatments) and availability of the study treatments. Once completed and electronically signed, the treatment allocation was displayed.

The following modifications were made to the Randomisation form during the trial:

| **Randomisation form version** | **Date of release** | **Major modifications from previous version** |
|---|---|---|
| 1.0 | 19-Mar-20 | Initial version (protocol V1.0) |
| 2.0 | 25-Mar-20 | For protocol V2.0<br>• Hydroxycholoroquine added as treatment<br>• Known long QT syndrome added to comorbidities<br>• Severe depression removed from comorbidities |
| 3.0 | 09-Apr-20 | For protocol V3.0<br>• Azithromycin added as treatment<br>• Suspected SARS-CoV-2 infection included in eligibility criteria |
| [Second randomisation form introduced] | 23-Apr-20 | For protocol 4.0<br>• Eligibility criteria for second randomisation<br>• Tocilizumab vs control as treatment allocations |
| 4.0 | 09-May-20 | For protocol V5.0<br>• Age ≥18 years removed from eligibility criteria<br>• Additional questions on child's age and weight added |
| 5.0 | 21-May-20 | For protocol V6.0<br>• Convalescent plasma added as treatment |
| 6.0 | 28-May-20 | Baseline use of remdesivir |
| 7.0 | 01-Jul-20 | For protocol V7.0<br>• Participants eligible if convalescent plasma is only available and suitable treatment<br>• Hydroxychloroquine, dexamethasone (adults) and lopinavir-ritonavir removed |
| 8.0 | 13-Aug-20 | For protocol V8.0<br>• Addition of low-dose and high-dose corticosteroids and intravenous immunoglobulin for children (and removal of dexamethasone for children) |
| 9.0 | 24-Sep-20 | For protocol V9.0<br>• REGEN-COV2 added as treatment<br>• Additional baseline information |
| 10.0 | 06-Nov-20 | For protocol V10.1<br>• Aspirin added as treatment |
| 11.0 | 27-Nov-20 | For protocol V11.1<br>• Colchicine added as treatment<br>• Azithromycin removed |

RECOVERY
Randomised Evaluation of COVID-19 Therapy

### Sample Form (v10.00 - 05/11/20)

#### Randomisation Program

Call Freefone **0800 138 5451** to contact the RECOVERY team for **URGENT** problems using the Randomisation Program or for medical advice. All **NON-URGENT queries** should be emailed to ****

Logged in as: **RECOVERY Site**

##### Section A: Baseline and Eligibility

Date and time of randomisation: 5 Nov 2020 14:00

**Treating clinician**

**A1.** Name of treating clinician

**Patient details**

**A2.** Patient surname

Patient forename

**A3.** NHS number ☐ Tick if not available

**A4.** What is the patient's date of birth? 01 / January / 2000

**A5.** What is the patient's sex?

**Inclusion criteria**

**A6.** Has consent been taken in line with the protocol?
If answer is No patient cannot be enrolled in the study

**A7.** Does the patient have proven or suspected SARS-CoV-2 infection?
If answer is No patient cannot be enrolled in the study

**A8.** Does the patient have any medical history that might, in the opinion of the attending clinician, put the patient at significant risk if they were to participate in the trial?

**A8B.** Is the patient willing to receive convalescent plasma?

**A9.** COVID-19 symptom onset date: / /

**A10.** Date of hospitalisation: / /

**A11.** Does the patient require oxygen?

**A12.** Please select one of the following to describe the current level of ventilation support

**A12.1** Enter latest oxygen saturation measurement (%)

**A12.2** Enter latest CRP measurement since admission to hospital (mg/L) ☐ Tick if not measured ☐ Tick if greater than limit of measurement
Enter 0 if below the limit of measurement

**A12.3** Enter latest creatinine measurement since admission to hospital (μmol/L) ☐ Tick if not measured

**A12.4** Enter latest D-dimer measurement since admission to hospital (ng/mL) ☐ Tick if not measured ☐ Tick if greater than limit of measurement
Enter 0 if below the limit of measurement

**Does the patient have any CURRENT comorbidities or other medical problems or treatments?**

**A13.1** Diabetes

**A13.2** Heart disease

**A13.3** Chronic lung disease

**A13.4** Tuberculosis

**A13.5** HIV

**A13.6** Severe liver disease

**A13.7** Severe kidney impairment (eGFR<30 or on dialysis)

**A13.8** Known long QT syndrome

**A13.9** Current treatment with macrolide antibiotics which are to continue
Macrolide antibiotics include clarithromycin, azithromycin and erythromycin

**A13.10** Antiplatelet therapy
Includes aspirin, clopidogrel, ticagrelor, prasugrel, dipyridamole

**A13.11** Previous adverse reaction to blood or blood product transfusion

**Are the following treatments UNSUITABLE for the patient?**
**If you answer Yes it means you think this patient should NOT receive this drug.**

**A14.1** Azithromycin

**A14B.1** Convalescent plasma

**A14B.2** Synthetic monoclonal antibodies (REGN10933+REGN10987)

**A14C.1** Aspirin

**Are the following treatments available?**

**A15.1** Azithromycin

**A15B.1** Convalescent plasma

**A15B.2** Synthetic monoclonal antibodies (REGN10933+REGN10987)

**A15C.1** Aspirin

**Current medication**

**A16.1** Is the patient currently prescribed remdesivir?

**A16.2** Is the participant currently prescribed systemic corticosteroids (dexamethasone, prednisolone, hydrocortisone, methylprednisolone)?
Please do not include topical or inhaled treatments

**A16.4** Is the patient currently on warfarin or a direct oral anticoagulant?
Includes apixaban, rivaroxaban

**A16.5** What venous thromboembolism prophylaxis is the patient receiving?
Standard = usual for hospitalised patients (not increased due to COVID-19); Higher dose = treatment dose or increased prophylaxis due to COVID-19

**Please sign off this form once complete**

Surname:

Forename:

Professional email:

Continue

Cancel

Home

*Follow-up form*

The Follow-up form (shown on the next page) collected information on study treatment adherence (including both the randomised allocation and use of other study treatments), vital status (including date and provisional cause of death if available), hospitalisation status (including date of discharge), respiratory support received during the hospitalisation, occurrence of any major cardiac arrhythmias and renal replacement therapy received. Questions on thrombotic and bleeding events were added with V10.1 of the protocol so these data were collected for only a small proportion of people in the azithromycin comparison.

The following modifications were made to the Follow-up form during the trial:

| **Follow-up form version** | **Date of release** | **Modifications from previous version** |
|---|---|---|
| 1.0 | 30-Mar-20 | Initial version |
| 2.0 | 09-Apr-20 | Information on other treatments used during admission:<br>• Azithromycin, IL-6 receptor antagonist<br>Fact and result of SARS-CoV-2 PCR test |
| 3.0 | 09-Apr-20 | Update to functionality; no changes to questions |
| 4.0 | 23-Apr-20 | Duration of treatments added |
| 5.0 | 12-May-20 | Capture of major cardiac arrhythmias added |
| 6.0 | 28-May-20 | Updates to wording of questions.<br>Information on other treatments used during admission:<br>• Remdesivir, convalescent plasma |
| 7.0 | 18-Jun-20 | Clarification of question wording |
| 8.0 | 10-Jul-20 | Information on new treatments for children adherence |
| 9.0 | 24-Sep-20 | Information on REGEN-COV2 adherence |
| 10.0 | 06-Nov-20 | Information on aspirin adherence<br>Capture of thrombotic and bleeding events added<br>Information of enrolment into other studies added |
| 11.0 | 16-Nov-20 | Minor changes to in-form validation |
| 12.0 | 27-Nov-20 | Information on colchicine adherence |

### Follow-up

#### Date of randomisation

**Please only report events that occurred from first randomisation until 28 days later on this form (except for Q2).**

**Patient's date of birth** *

yyyy-mm-dd

**1. Which of following treatment(s) did the patient definitely receive as part of their hospital admission after randomisation?** *

*(NB Include RECOVERY study-allocated drug, only if given, PLUS any of the other treatments if given as standard hospital care)*

- [ ] No additional treatment
- [ ] Lopinavir-ritonavir
- [ ] Corticosteroid (dexamethasone, prednisolone, hydrocortisone or methylprednisolone)
- [ ] Hydroxychloroquine
- [ ] Azithromycin or other macrolide (eg, clarithromycin, erythromycin)
- [ ] Tocilizumab or sarilumab
- [ ] Remdesivir
- [ ] Intravenous immunoglobulin
- [ ] Synthetic monoclonal antibodies (REGN10933+REGN10987)
- [ ] Aspirin

**Please select number of days the patient received lopinavir-ritonavir**

○ 1 ○ 2 ○ 3 ○ 4 ○ 5 ○ 6 ○ 7 ○ 8 ○ 9 ○ 10

**Please select number of days the patient received corticosteroid (dexamethasone, prednisolone, hydrocortisone or methylprednisolone)**

○ 1 ○ 2 ○ 3 ○ 4 ○ 5 ○ 6 ○ 7 ○ 8 ○ 9 ○ 10

**Please select number of days the patient received hydroxychloroquine**

○ 1 ○ 2 ○ 3 ○ 4 ○ 5 ○ 6 ○ 7 ○ 8 ○ 9 ○ 10

**Please select number of days the patient received azithromycin**

○ 0 ○ 1 ○ 2 ○ 3 ○ 4 ○ 5 ○ 6 ○ 7 ○ 8 ○ 9 ○ 10

**Please select number of days the patient received other macrolides (eg, clarithromycin, erythromycin)**

○ 0 ○ 1 ○ 2 ○ 3 ○ 4 ○ 5 ○ 6 ○ 7 ○ 8 ○ 9 ○ 10

**Please select number of doses of tocilizumab or sarilumab the patient received**

○ 1 ○ >1

**Please select number of days the patient received remdesivir**

○ 1 ○ 2 ○ 3 ○ 4 ○ 5 ○ 6 ○ 7 ○ 8 ○ 9 ○ 10

**Please select the proportion of days the patient received aspirin or other antiplatelet (eg, clopidogrel, prasugrel, ticagrelor, dipyridamole)**

○ Most days (≥90%) ○ Some days (≥50% <90%) ○ Few days (<50% of days, but not zero) ○ None

#### » Convalescent Plasma

**How many convalescent plasma infusions did the patient receive?**

*This is plasma given as part of a trial, not any standard fresh frozen plasma or other blood products that the patient may have been given*

○ 0 ○ 1 ○ 2

**Were any infusions stopped early for any reason ie, the patient did not receive the full amount?**

○ Yes ○ No

**How many were stopped early?**

○ 1 ○ 2

#### » Health Status

**2. Was a COVID-19 test done for this patient at any point during the admission?** *

*(If multiple tests were done, and the results were positive and negative, please tick Yes – positive result and Yes – negative result)*

☐ Yes – positive result

☐ Yes – negative result

☐ Not done

**3. What is the patient's vital status?** *

○ Alive

○ Dead

**3.1 What is the patient's current hospitalisation status?** *

○ Inpatient

○ Discharged

The patient has been enrolled in the trial for **NaN** days

**3.1.1 Date follow-up form completed**

yyyy-mm-dd

**3.1.1 What was the date of discharge?** *

yyyy-mm-dd

**3.1 What was the date of death?** *

yyyy-mm-dd

**3.2 What was the underlying cause of death?** *

*This can be obtained from the last entry in part 1 of the death certificate*

○ COVID-19

○ Other infection

○ Cardiovascular

○ Other

**Please give details**

**4. Did the patient require any form of assisted ventilation (ie, more than just supplementary oxygen) from day of randomisation until 28 days later?** *

○ Yes

○ No

**Please answer the following questions:**

**4.1 For how many days did the patient require assisted ventilation?** *

**4.2 What type of ventilation did the patient receive?**

| | Yes | No | Unknown |
|---|---|---|---|

| | | | |
|---|---|---|---|
| CPAP alone | ○ | ○ | ○ |
| Non-invasive ventilation (eg, BiPAP) | ○ | ○ | ○ |
| High-flow nasal oxygen (eg, AIRVO) | ○ | ○ | ○ |
| Mechanical ventilation (intubation/tracheostomy) | ○ | ○ | ○ |
| ECMO | ○ | ○ | ○ |

**Total number of days the patient received invasive mechanical ventilation (intubation/tracheostomy) from randomisation until discharge/death/28 days after randomisation**

**5. Has the patient been documented to have a NEW cardiac arrhythmia at any point since the main randomisation until 28 days later?** *

- ○ Yes
- ○ No
- ○ Unknown

**5.1 Please select all of the following which apply**

- ☐ Atrial flutter or atrial fibrillation
- ☐ Supraventricular tachycardia
- ☐ Ventricular tachycardia (including torsades de pointes)
- ☐ Ventricular fibrillation
- ☐ Atrioventricular block requiring intervention (eg, cardiac pacing)

**6. Did the patient require use of renal dialysis or haemofiltration from main randomisation until 28 days later?** *

- ○ Yes
- ○ No

**7. During the first 28 days after randomisation, did the participant have a thrombotic event?** *

- ○ Yes
- ○ No
- ○ Unknown

**7.1 Please indicate the type of thrombotic event**

##### 7.1 Please indicate the type of thrombotic e[illegible]

*Select all that apply*

- [ ] Pulmonary embolism
- [ ] Deep-vein thrombosis
- [ ] Ischaemic stroke
- [ ] Myocardial infarction
- [ ] Systemic arterial embolism
- [ ] Other

##### 8. During the first 28 days after randomisation, did the participant experience clinically-significant bleeding ie, intra-cranial bleeding or bleeding that required intervention (eg, surgery, endoscopy or vasoactive drugs) or a blood transfusion? *

- ○ Yes
- ○ No
- ○ Unknown

##### 8.1 Please indicate the site(s) of bleeding *

*Select all that apply*

- [ ] Intra-cranial
- [ ] Gastrointestinal
- [ ] Other

##### 8.2 Please indicate which interventions were required to manage the bleed *

*Select all that apply*

- [ ] Blood transfusion
- [ ] Surgery
- [ ] Endoscopy
- [ ] Vasoactive drugs (e.g. inotropes on ICU)
- [ ] None of the the above

##### 9. Please enter UKOSS case ID if known *

*Enter the full UKOSS case ID ie, COR_123*

..............................

*(select if you do not know the UKOSS case ID)*

- [ ] Not known

##### 10. Please indicate if the participant participated in any other COVID-19 trials

*Select all that apply*

- [ ] PRINCIPLE
- [ ] REMAP-CAP
- [ ] Other treatment trial(s)
- [ ] COVID-19 vaccine trial(s)

**Please give name of other treatment trial(s)**

..........

**Please give name of COVID-19 vaccine trial(s)**

..........

**Interim analyses: role of the Data Monitoring Committee**

The independent Data Monitoring Committee reviews unblinded analyses of the study data and any other information considered relevant at intervals of around 2 weeks. The committee is charged with determining if, in their view, the randomised comparisons in the study provide evidence on mortality that is strong enough (with a range of uncertainty around the results that was narrow enough) to affect national and global treatment strategies. In such a circumstance, the Committee would inform the Steering Committee who would make the results available to the public and amend the trial arms accordingly. Unless that happened, the Steering Committee, investigators, and all others involved in the trial would remain blind to the interim results until 28 days after the last patient had been randomised to a particular intervention arm. Further details about the role and membership of the independent Data Monitoring Committee are provided in the protocol.

The Data Monitoring Committee determined that to consider recommending stopping a treatment early for benefit would require at least a 3 to 3·5 standard error reduction in mortality. The Committee concluded that examinations of the data at every 10% (or even 5%) of the total data would lead to only a marginal increase in the overall type I error rate.

### Supplementary Tables

#### Webtable 1: Baseline characteristics of patients considered unsuitable for randomisation to azithromycin compared with those randomised to azithromycin versus usual care

| | **Randomised (n=7764)** | **Unsuitable (n=4570)** |
|---|---|---|
| Age, years | 65.3 (15.7) | 66.0 (15.7) |
| <70 | 4523 (58%) | 2618 (57%) |
| ≥70 to <80 | 1782 (23%) | 998 (22%) |
| ≥80 | 1459 (19%) | 954 (21%) |
| Sex | | |
| Male | 4819 (62%) | 2963 (65%) |
| Female | 2945 (38%) | 1607 (35%) |
| Ethnicity | | |
| White | 5640 (73%) | 3256 (71%) |
| Black, Asian, and minority ethnic | 1071 (14%) | 731 (16%) |
| Unknown | 1053 (14%) | 583 (13%) |
| Number of days since symptom onset | 8 (5-11) | 8 (5-12) |
| Number of days since admission to hospital | 2 (1-4) | 2 (1-4) |
| Respiratory support received | | |
| No oxygen received | 1408 (18%) | 818 (18%) |
| Oxygen only * | 5904 (76%) | 3351 (73%) |
| Invasive mechanical ventilation | 452 (6%) | 401 (9%) |
| Previous diseases | | |
| Diabetes | 2133 (27%) | 1319 (29%) |
| Heart disease | 2043 (26%) | 1301 (28%) |
| Chronic lung disease | 1935 (25%) | 1138 (25%) |
| Tuberculosis | 19 (<1%) | 24 (<1%) |
| HIV | 29 (<1%) | 18 (<1%) |
| Severe liver disease † | 110 (1%) | 100 (2%) |
| Severe kidney impairment ‡ | 489 (6%) | 340 (7%) |
| Any of the above | 4521 (58%) | 2796 (61%) |
| Use of corticosteroids | | |
| Yes | 4739 (61%) | 1727 (38%) |
| No | 582 (7%) | 160 (4%) |
| Unknown^ | 2443 (31%) | 2683 (59%) |
| Severe acute respiratory syndrome coronavirus 2 test result | | |
| Positive | 6916 (89%) | 3963 (87%) |
| Negative | 566 (7%) | 474 (10%) |
| Unknown | 282 (4%) | 133 (3%) |

Data are mean (SD), n (%), or median (IQR). * Includes non-invasive ventilation. † Defined as requiring ongoing specialist care. ‡ Defined as estimated glomerular filtration rate <30 mL/min per 1.73 $m^2$. ^ Information on use of corticosteroids was collected from 18 June 2020 onwards following announcement of the results of the dexamethasone comparison from the RECOVERY trial.

#### Webtable 2: Treatments given, by randomised allocation

| | Treatment allocation | |
|---|---|---|
| | **Azithromycin (n=2582)** | **Usual care (n=5182)** |
| Compliance data available | 1986 | 3924 |
| Received azithromycin | 1760 (89%) | 55 (1%) |
| Received other macrolide | 317 (16%) | 558 (14%) |
| Subtotal: Received azithromycin and/or other macrolide | 1836 (92%) | 606 (15%) |
| Other treatments received | | |
| Lopinavir-ritonavir | 3 (<1%) | 6 (<1%) |
| Corticosteroid | 918 (46%) | 1925 (49%) |
| Hydroxychloroquine | 2 (<1%) | 8 (<1%) |
| Tocilizumab or sarilumab | 126 (6%) | 305 (8%) |
| Remdesivir | 407 (20%) | 862 (22%) |
| Convalescent plasma | 336 (17%) | 689 (18%) |
| REGEN-COV2 | 55 (3%) | 103 (3%) |

Percentages are of those with a completed follow-up form. Of those allocated azithromycin who received at least one dose, 75% received it every day they were in hospital (or every day except one) and 90% received it on at least half the days they were in hospital. The median number of days it was taken was 6 days (IQR 3-9 days).

#### Webtable 3: Effect of allocation to azithromycin on cause-specific 28-day mortality

| | Treatment allocation | | |
|---|---|---|---|
| Cause of death | Azithromycin (n=2582) | Usual care (n=5182) | Absolute difference, % (95% CI) |
| COVID | 353 (13.7%) | 726 (14.0%) | -0.34 (-1.97,1.29) |
| Other infection | 10 (0.4%) | 14 (0.3%) | 0.12 (-0.16,0.40) |
| Cardiac | 9 (0.3%) | 6 (0.1%) | 0.23 (-0.01,0.48) |
| Stroke | 3 (0.1%) | 6 (0.1%) | 0.00 (-0.16,0.16) |
| Other vascular | 6 (0.2%) | 4 (0.1%) | 0.16 (-0.05,0.36) |
| Cancer | 9 (0.3%) | 14 (0.3%) | 0.08 (-0.19,0.35) |
| Other medical | 24 (0.9%) | 52 (1.0%) | -0.07 (-0.53,0.39) |
| External | 2 (0.1%) | 0 (0.0%) | 0.08 (-0.03,0.18) |
| Unknown cause | 80 (3.1%) | 175 (3.4%) | -0.28 (-1.11,0.55) |
| Total: 28-day mortality | 496 (19.2%) | 997 (19.2%) | -0.03 (-1.89,1.83) |

#### Webtable 4: Effect of allocation to azithromycin on cardiac arrhythmia

| | Treatment allocation | |
|---|---|---|
| | Azithromycin (n=2582) | Usual care (n=5182) |
| Number with follow-up form* | 1794 | 3540 |
| Atrial flutter or atrial fibrillation | 55 (3.1%) | 121 (3.4%) |
| Other supraventricular tachycardia | 13 (0.7%) | 19 (0.5%) |
| Subtotal: Supraventricular tachycardia | 65 (3.6%) | 136 (3.8%) |
| Ventricular tachycardia | 11 (0.6%) | 15 (0.4%) |
| Ventricular fibrillation | 1 (0.1%) | 2 (0.1%) |
| Subtotal: Ventricular tachycardia or fibrillation | 11 (0.6%) | 17 (0.5%) |
| Atrioventricular block requiring intervention | 2 (0.1%) | 7 (0.2%) |
| Total: Any major cardiac arrhythmia | 78 (4.3%) | 157 (4.4%) |

* Information on new cardiac arrhythmias was only collected on follow-up forms from 12 May 2020 onwards; percentages are of those with such a form completed.

### Supplementary Figures

### Webfigure 1: Effect of allocation to azithromycin on hospital discharge by baseline characteristics

| | Azithromycin | Usual care | RR (95% CI) |
|---|---|---|---|
| **Age, years ($\chi^2_1$= 2.6; p=0.11)** | | | |
| <70 | 1015/1508 (67%) | 2047/3015 (68%) | 1.00 (0.93−1.08) |
| ≥ 70 <80 | 311/615 (51%) | 573/1167 (49%) | 1.03 (0.90−1.19) |
| ≥ 80 | 228/459 (50%) | 446/1000 (45%) | 1.17 (0.99−1.39) |
| **Sex ($\chi^2_1$= 10.7; p=0.001)** | | | |
| Men | 902/1603 (56%) | 1855/3216 (58%) | 0.96 (0.88−1.04) |
| Women | 652/979 (67%) | 1211/1966 (62%) | 1.19 (1.07−1.31) |
| **Ethnicity ($\chi^2_1$=0.0; p=0.84)** | | | |
| White | 1150/1867 (62%) | 2287/3773 (61%) | 1.03 (0.96−1.11) |
| Black, Asian and Minority Ethnic | 229/363 (63%) | 447/708 (63%) | 1.05 (0.89−1.24) |
| **Days since symptom onset ($\chi^2_1$= 0.0; p=0.94)** | | | |
| ≤ 7 | 694/1163 (60%) | 1383/2355 (59%) | 1.03 (0.94−1.14) |
| >7 | 859/1418 (61%) | 1676/2812 (60%) | 1.03 (0.95−1.12) |
| **Respiratory support at randomisation ($\chi^2_1$= 0.6; p=0.44)** | | | |
| No oxygen received | 370/490 (76%) | 701/918 (76%) | 0.99 (0.87−1.13) |
| Oxygen only* | 1156/1940 (60%) | 2317/3964 (58%) | 1.04 (0.97−1.12) |
| Invasive mechanical ventilation | 28/152 (18%) | 48/300 (16%) | 1.17 (0.73−1.89) |
| **Use of corticosteroids ($\chi^2_1$= 0.4; p=0.53)** | | | |
| Yes | 884/1567 (56%) | 1781/3172 (56%) | 1.03 (0.95−1.12) |
| No | 116/183 (63%) | 267/399 (67%) | 0.95 (0.76−1.19) |
| Unknown | 554/832 (67%) | 1018/1611 (63%) | 1.07 (0.96−1.19) |
| **Baseline risk ($\chi^2_1$= 0.5; p=0.48)** | | | |
| <30% | 1194/1718 (69%) | 2350/3389 (69%) | 1.02 (0.95−1.09) |
| ≥ 30% <45% | 246/523 (47%) | 497/1058 (47%) | 0.99 (0.84−1.15) |
| ≥ 45% | 114/341 (33%) | 219/735 (30%) | 1.17 (0.93−1.48) |
| **All participants** | **1554/2582 (60%)** | **3066/5182 (59%)** | **1.03 (0.97−1.10) p=0.29** |

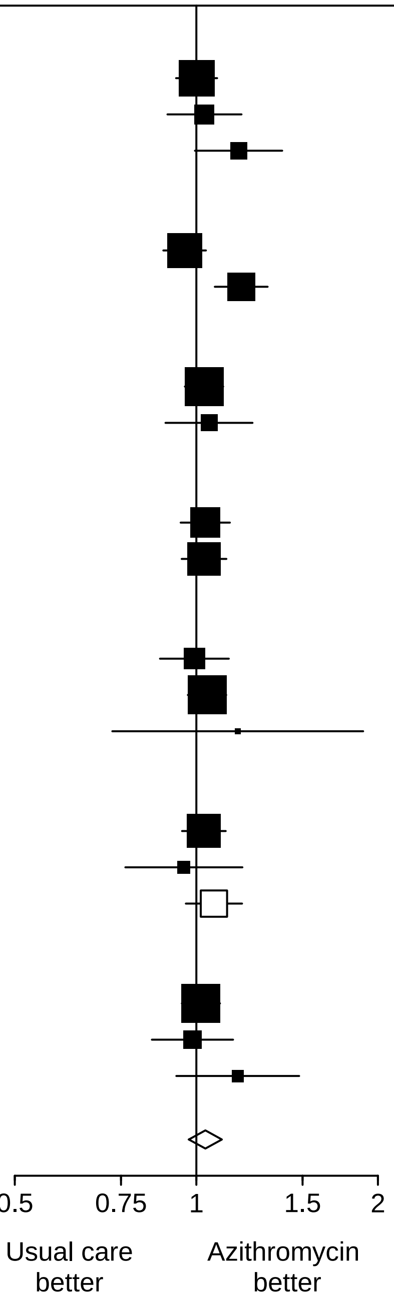

### Webfigure 2: Effect of allocation to azithromycin on invasive mechanical ventilation or death in those not on invasive mechanical ventilation at randomisation, by baseline characteristics

| | Azithromycin | Usual care | RR (95% CI) |
|---|---|---|---|
| **Age, years ($\chi^2_1$= 1.1; p=0.29)** | | | |
| <70 | 180/1392 (13%) | 350/2786 (13%) | 1.03 (0.87−1.22) |
| ≥ 70 <80 | 173/583 (30%) | 328/1103 (30%) | 1.00 (0.86−1.16) |
| ≥ 80 | 164/455 (36%) | 391/993 (39%) | 0.92 (0.79−1.06) |
| **Sex ($\chi^2_1$= 1.7; p=0.20)** | | | |
| Men | 352/1497 (24%) | 695/3000 (23%) | 1.01 (0.91−1.14) |
| Women | 165/933 (18%) | 374/1882 (20%) | 0.89 (0.75−1.05) |
| **Ethnicity ($\chi^2_1$=4.1; p=0.04)** | | | |
| White | 422/1788 (24%) | 834/3628 (23%) | 1.03 (0.93−1.14) |
| Black, Asian and Minority Ethnic | 57/319 (18%) | 146/620 (24%) | 0.76 (0.58−1.00) |
| **Days since symptom onset ($\chi^2_1$= 0.2; p=0.69)** | | | |
| ≤ 7 | 277/1123 (25%) | 586/2277 (26%) | 0.96 (0.85−1.08) |
| >7 | 240/1306 (18%) | 478/2590 (18%) | 1.00 (0.87−1.15) |
| **Respiratory support at randomisation ($\chi^2_1$= 0.5; p=0.49)** | | | |
| No oxygen received | 68/490 (14%) | 119/918 (13%) | 1.07 (0.81−1.41) |
| Oxygen only* | 449/1940 (23%) | 950/3964 (24%) | 0.97 (0.88−1.07) |
| **Use of corticosteroids ($\chi^2_1$= 0.0; p=0.83)** | | | |
| Yes | 287/1495 (19%) | 592/3024 (20%) | 0.98 (0.86−1.11) |
| No | 24/177 (14%) | 55/378 (15%) | 0.93 (0.60−1.45) |
| Unknown | 206/758 (27%) | 422/1480 (29%) | 0.95 (0.83−1.10) |
| **Baseline risk ($\chi^2_1$= 1.9; p=0.16)** | | | |
| <30% | 235/1669 (14%) | 437/3286 (13%) | 1.06 (0.91−1.23) |
| ≥ 30% <45% | 163/499 (33%) | 345/1021 (34%) | 0.97 (0.83−1.13) |
| ≥ 45% | 119/262 (45%) | 287/575 (50%) | 0.91 (0.78−1.06) |
| **All participants** | **517/2430 (21%)** | **1069/4882 (22%)** | **0.97 (0.89−1.07) p=0.54** |

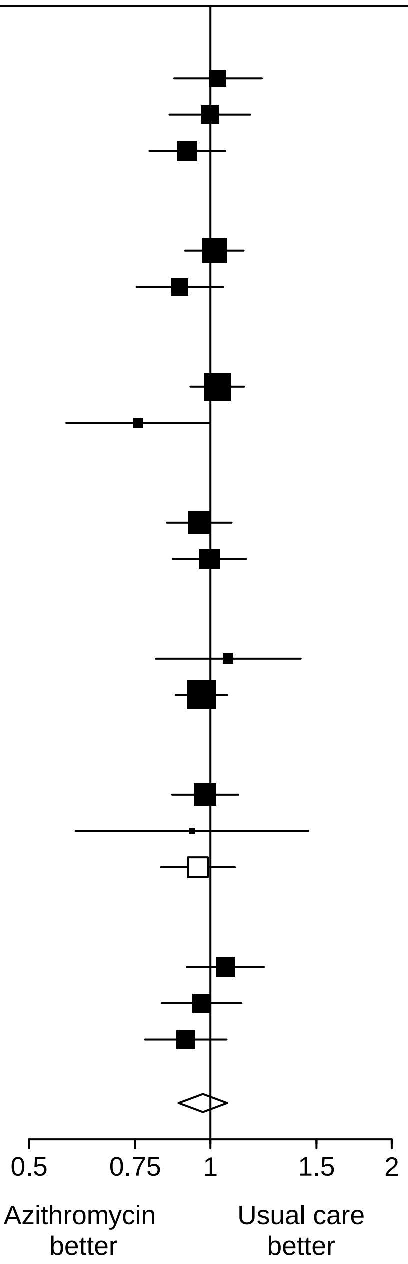
