## Supplementary material for "Azithromycin in Hospitalised Patients with COVID-19 (RECOVERY): a randomised, controlled, open-label, platform trial": Protocol and Statistical Analysis Plan

#### SUPPLEMENTARY APPENDIX
#### PROTOCOL AND STATISTICAL ANALYSIS PLAN

**RECOVERY Collaborative Group**

**Contents**

**RECOVERY Trial Protocols** ........ **2**
RECOVERY Trial Protocol V3.0 3
RECOVERY Trial Protocol V10.1 27
Substantial amendments to RECOVERY trial protocol 67
**Statistical Analysis Plans** ........ **69**
Statistical Analysis Plan V1.0 70
Statistical Analysis Plan V2.1 (including summary of changes) 91

### RECOVERY Trial Protocols

### RECOVERY Trial Protocol V3.0

RECOVERY
Randomised Evaluation of COVID-19 Therapy

### RANDOMISED EVALUATION OF COVID-19 THERAPY (RECOVERY)

**Background:** In early 2020, as this protocol was being developed, there were no approved treatments for COVID-19, a disease induced by the novel coronavirus SARS-CoV-2 that emerged in China in late 2019. The UK New and Emerging Respiratory Virus Threats Advisory Group (NERVTAG) advised that several possible treatments should be evaluated, including Lopinavir-Ritonavir, low-dose corticosteroids, and hydroxychloroquine. These groups also advised that other treatments will soon emerge that require evaluation. A World Health Organization (WHO) expert group issued broadly similar advice.

**Eligibility and randomisation:** This protocol describes a randomised trial among adults hospitalised for COVID-19. All eligible patients are randomly allocated between several treatment arms, each to be given in addition to the usual standard of care in the participating hospital: No additional treatment *vs* Lopinavir-Ritonavir *vs* Low-dose Corticosteroids *vs* Hydroxychloroquine *vs* Azithromycin. For patients for whom not all the trial arms are appropriate or at locations where not all are available, randomisation will be between fewer arms.

**Adaptive design:** The interim trial results will be monitored by an independent Data Monitoring Committee (DMC). The most important task for the DMC will be to assess whether the randomised comparisons in the study have provided evidence on mortality that is strong enough (with a range of uncertainty around the results that is narrow enough) to affect national and global treatment strategies. In such a circumstance, the DMC will inform the Trial Steering Committee who will make the results available to the public and amend the trial arms accordingly. New trial arms can be added as evidence emerges that other candidate therapeutics should be evaluated.

**Outcomes:** The main outcomes will be death, discharge, need for ventilation and need for renal replacement therapy. For the main analyses, follow-up will be censored at 28 days after randomisation. Additional information on longer term outcomes may be collected through review of medical records or linkage to medical databases such as those managed by NHS Digital and equivalent organisations in the devolved nations.

**Simplicity of procedures:** To facilitate collaboration, even in hospitals that suddenly become overloaded, patient enrolment (via the internet) and all other trial procedures are greatly streamlined. Informed consent is simple and data entry is minimal. Randomisation via the internet is simple and quick, at the end of which the allocated treatment is displayed on the screen and can be printed or downloaded. Follow-up information is recorded at a single timepoint and may be ascertained by contacting participants in person, by phone or electronically, or by review of medical records and databases.

**Data to be recorded:** At randomisation, information will be collected on the identity of the randomising clinician and of the patient, age, sex, major co-morbidity, pregnancy, COVID-19 onset date and severity, and any contraindications to the study treatments. The main outcomes will be death (with date and probable cause), discharge (with date), need for ventilation (with number of days recorded) and need for renal replacement therapy. Reminders will be sent if outcome data have not been recorded by 28 days after randomisation. Suspected Unexpected Serious Adverse Reactions (SUSARs) to one of the study medication (eg, Stevens-Johnson syndrome, anaphylaxis, aplastic anaemia) will be collected and reported in an expedited fashion. Other adverse events will not be recorded but may be available through linkage to medical databases.

**Numbers to be randomised:** The larger the number randomised the more accurate the results will be, but the numbers that can be randomised will depend critically on how large the epidemic becomes. If substantial numbers are hospitalised in the participating centres then it may be possible to randomise several thousand with mild disease and a few thousand with severe disease, but realistic, appropriate sample sizes could not be estimated at the start of the trial.

**Heterogeneity between populations:** If sufficient numbers are studied, it may be possible to generate reliable evidence in certain patient groups (e.g. those with major co-morbidity or who are older). To this end, data from this study may be combined with data from other trials of treatments for COVID-19, such as those being planned by the WHO.

**Add-on studies:** Particular countries or groups of hospitals, may well want to collaborate in adding further measurements or observations, such as serial virology, serial blood gases or chemistry, serial lung imaging, or serial documentation of other aspects of disease status. While well-organised additional research studies of the natural history of the disease or of the effects of the trial treatments could well be valuable (although the lack of placebo control may bias the assessment of subjective side-effects, such as gastro-intestinal problems), they are not core requirements.

**To enquire about the trial, contact the RECOVERY Central Coordinating Office**

---

RECOVERY Central Coordinating Office:
Nuffield Department of Population Health
Richard Doll Building
Old Road Campus
Roosevelt Drive
Oxford OX3 7LF
United Kingdom

Website: www.recoverytrial.net

---

**To RANDOMISE a patient, visit:**

---

Website: www.recoverytrial.net

---

RECOVERY
Randomised Evaluation of COVID-19 Therapy

RECOVERY
Randomised Evaluation of COVID-19 Therapy

### 1 BACKGROUND AND RATIONALE

##### 1.1.1 Setting

In 2019 a novel coronavirus-induced disease (COVID-19) emerged in Wuhan, China. A month later the Chinese Center for Disease Control and Prevention identified a new beta-coronavirus (SARS coronavirus 2, or SARS-CoV-2) as the aetiological agent.[1] The clinical manifestations of COVID-19 range from asymptomatic infection or mild, transient symptoms to severe viral pneumonia with respiratory failure. As many patients do not progress to severe disease the overall case fatality rate per infected individual is low, but hospitals in areas with significant community transmission have experienced a major increase in the number of hospitalised pneumonia patients, and the frequency of severe disease in hospitalised patients can be as high as 30%.[2-4] The progression from prodrome (usually fever, fatigue and cough) to severe pneumonia requiring oxygen support or mechanical ventilation often takes one to two weeks after the onset of symptoms.[2] The kinetics of viral replication in the respiratory tract are not well characterized, but this relatively slow progression provides a potential time window in which antiviral therapies could influence the course of disease.

##### 1.1.2 Treatment Options

There are currently no approved anti-viral or host-directed treatments for COVID-19. This protocol allows reliable assessment of the effects of multiple different treatments (including re-purposed and novel drugs) on major outcomes in COVID-19. All patients will receive usual care for the participating hospital.

Randomisation may be between the following treatment arms (although not all arms may be available at any one time):

**No additional treatment:** There are currently no approved anti-viral or host-directed treatments for COVID-19.

**Lopinavir-Ritonavir:** Lopinavir is a human immunodeficiency virus 1 (HIV-1) protease inhibitor, which is combined with ritonavir to increase lopinavir's plasma half-life. Lopinavir-Ritonavir has shown activity against SARS and MERS CoVs.

**Low dose corticosteroids:** Favourable immune response modulation by low-dose corticosteroids might help treat severe acute respiratory coronavirus infections, including COVID-19, SARS and MERS.

**Hydroxychloroquine:** Hydroxychloroquine, a derivative of chloroquine, has been used for many decades to treat malaria and rheumatological diseases. It has antiviral activity against SARS-CoV-2 in cell culture.

**Azithromycin:** Azithromycin is a macrolide antibiotic with immunomodulatory properties that has shown benefit in inflammatory lung disease.

Further details on each of these treatment options is provided in Appendix 1 (see section 9.1.1).

**Modifications to the number of treatment arms:** Other arms can be added if evidence emerges that there are suitable candidate therapeutics. Conversely, in some patient populations, not all trial arms are appropriate (e.g. due to contraindications based on co-morbid conditions or concomitant medication); in some hospitals, not all treatment arms will be available (e.g. due to manufacturing and supply shortages); and at some times, not all treatment arms will be active (e.g. due to lack of relevant approvals and contractual agreements). In any of these situations, randomisation will be between fewer arms.

##### 1.1.3 Design Considerations

The RECOVERY Protocol describes an overarching trial design to provide reliable evidence on the efficacy of candidate therapies for suspected or confirmed COVID-19 infection in hospitalised adult patients receiving usual standard of care.

There are no known treatments for COVID-19. The anticipated scale of the epidemic is such that hospitals, and particularly intensive care facilities, may be massively overstretched. Under some models of pandemic spread, up to 50% of the adult population may fall sick over a period of 8-12 weeks, of whom around 10% may require hospitalisation. This would involve about 2 million hospital admissions. In this situation, even treatments with only a moderate impact on survival or on hospital resources could be worthwhile. Therefore, the focus of RECOVERY is the impact of candidate treatments on mortality and on the need for hospitalisation or ventilation.

Critically, the trial is designed to minimise the burden on front-line hospital staff working within an overstretched care system during a major epidemic. Eligibility criteria are therefore simple and trial processes (including paperwork) are minimised.

The protocol is deliberately flexible so that it is suitable for a wide range of settings, allowing:

- a broad range of patients to be enrolled in large numbers;
- randomisation between only those treatment arms that are *both* available at the hospital *and* not believed by the enrolling doctor to be contraindicated (e.g. by particular co-morbid conditions or concomitant medications);
- treatment arms to be added or removed according to the emerging evidence; and
- additional sub-studies may be added to provide more detailed information on side effects or sub-categorisation of patient types but these are not the primary objective and are not required for participation.

In a cohort of 191 hospitalised COVID-19 patients with a completed outcome, the median time from illness onset to discharge was 22·0 days (IQR 18·0–25·0) and the median time to death was 18·5 days (15·0–22·0). Thirty-two patients (17%) required invasive mechanical ventilation and the median time from onset to mechanical ventilation was 14.5 days. Therefore, early endpoint assessment, such as 28 days after randomisation, is likely to provide largely complete outcome data and will permit early assessment of treatment efficacy and safety.[5]

### 2 DESIGN AND PROCEDURES

##### 2.1.1 Eligibility

Patients are eligible for the study if all of the following are true:

(i) Aged at least 18 years
(ii) Hospitalised
(iii) SARS-CoV-2 infection (clinically suspected or laboratory confirmed)
(iv) No medical history that might, in the opinion of the attending clinician, put the patient at significant risk if he/she were to participate in the trial

In addition, if the attending clinician believes that there is a specific contra-indication to one of the active drug treatment arms (see Appendix 2; section 9.1.2) or that the patient should definitely be receiving one of the active drug treatment arms then that arm will not be available for randomisation for that patient.

##### 2.1.2 Consent

Informed consent should be obtained from each patient before enrolment into the study. However, if the patient lacks capacity to give consent due to the severity of their medical condition (e.g. acute respiratory failure or need for immediate ventilation) or prior disease, then consent may be obtained from a relative acting as the patient's legally designated representative or independent doctor. Further consent will then be sought with the patient if they recover sufficiently.

Due to the poor outcomes in COVID-19 patients who require ventilation (>90% mortality in one cohort[5]), patients who lack capacity to consent due to severe disease (e.g. needs ventilation), and for whom a relative to act as the legally designated representative is not immediately available, randomisation and consequent treatment will proceed with consent provided by a treating clinician (independent of the clinician seeking to enrol the patient) who will act as the legally designated representative. Consent will then be obtained from the patient's personal legally designated representative (or directly from the patient if they recover promptly) at the earliest opportunity.

##### 2.1.3 Baseline information

The following information will be recorded on the web-based form by the attending clinician or delegate:

- Patient details (e.g. name, NHS number, date of birth, sex)
- Clinician details (e.g. name)
- COVID-19 symptom onset date
- COVID-19 severity as assessed by need for supplemental oxygen or ventilation/extracorporeal membrane oxygenation
- Major comorbidity (e.g. heart disease, diabetes, chronic lung disease) and pregnancy
- Date of hospitalisation
- Contraindication to the study drug regimens (in the opinion of the attending clinician)

- Name of person completing the form

The person completing the form will then be asked to confirm that they wish to randomise the patient and will then be required to enter their name and e-mail address.

##### 2.1.4 Randomisation

Eligible patients will be allocated using a central web-based randomisation service (without stratification or minimisation) in to one of the following treatment arms (in addition to usual care):

- **No additional treatment**

- **Lopinavir 400mg-Ritonavir 100mg** by mouth (or nasogastric tube) every 12 hours for 10 days.

- **Corticosteroid** in the form of dexamethasone administered as an oral (liquid or tablets) or intravenous preparation 6 mg once daily for 10 days. In pregnancy, prednisolone 40 mg administered by mouth (or intravenous hydrocortisone 80 mg twice daily) should be used instead of dexamethasone.
(Note: It is permitted to switch between the two routes of administration according to clinical circumstances.)

- **Hydroxychloroquine** by mouth for a total of 10 days as follows:

| Timing | Dose |
|---|---|
| Initial | 800 mg |
| 6 hours after initial dose | 800 mg |
| 12 hours after initial dose | 400 mg |
| 24 hours after initial dose | 400 mg |
| Every 12 hours thereafter for 9 days | 400 mg |

- **Azithromycin 500mg** by mouth (or nasogastric tube) or intravenously once daily for 10 days.

Study treatments do not need to be continued after discharge from hospital.

The randomisation program will allocate patients in a ratio of 2:1 between the no additional care arm and each of the other arms available. Hence if 5 arms are available, then the randomisation will be in the ratio 2:1:1:1:1. If one or more of the active drug treatments is not available at the hospital or is believed, by the attending clinician, to be contraindicated (or definitely indicated) for the specific patient, then this fact will be recorded via the web-based form prior to randomisation; random allocation will then be between the remaining arms (i.e. in a 2:1:1:1, 2:1:1 or 2:1 ratio).

##### 2.1.5 Administration of allocated treatment

The details of the allocated study treatment will be displayed on the screen and can be printed or downloaded. The hospital clinicians are responsible for administration of the allocated treatment. The patient's own doctors are free to modify or stop study treatment if they feel it is in the best interests of the patient without the need for the patient to withdraw from the study (see section 2.1.8). This study is being conducted within hospitals. Therefore use of medication will be subject to standard pharmacy reviews (typically within 48 hours of enrolment) which will guide modifications to both the study treatment and use of concomitant medication (e.g. in the case of potential drug interactions).

##### 2.1.6 Collecting follow-up information

The following information will be ascertained at the time of death or discharge or at 28 days after randomisation (whichever is sooner):

- Vital status (alive / dead, with date and presumed cause of death, if appropriate)
- Hospitalisation status (inpatient / discharged, with date of discharge, if appropriate)
- Use of ventilation (with days of use and type, if appropriate)
- Use of renal dialysis or haemofiltration

This information will be obtained and entered into the web-based IT system by a member of the hospital clinical or research staff.

Follow-up information is to be collected on all study participants, irrespective of whether or not they complete the scheduled course of allocated study treatment. Study staff will seek follow-up information through various means including medical staff, reviewing information from medical notes, routine healthcare systems, and registries.

##### 2.1.7 Duration of follow-up

All randomised participants are to be followed up until death, discharge from hospital or 28 days after randomisation (whichever is sooner). It is recognised that in the setting of this trial, there may be some variability in exactly how many days after randomisation, information on disease status is collected. This is acceptable and will be taken account of in the analyses and interpretation of results, the principle being that some information about post-randomisation disease status is better than none.

Longer term (up to 10 years) follow-up will be sought through linkage to electronic healthcare records and medical databases including those held by NHS Digital, Public Health England and equivalent bodies, and to relevant research databases (e.g. UK Biobank, Genomics England).

##### 2.1.8 Withdrawal of consent

A decision by a participant that they no longer wish to continue receiving study treatment should **not** be considered to be a withdrawal of consent for follow-up. However, participants are free to withdraw consent for some or all aspects of the study at any time if they wish to do so. In accordance with regulatory guidance, de-identified data that have already been collected and incorporated in the study database will continue to be used (and any identifiable data will be destroyed).

For participants who lack capacity, if their legal representative withdraws consent for treatment or methods of follow-up then these activities would cease.

### 3 STATISTICAL ANALYSIS

All analyses for reports, presentations and publications will be prepared by the coordinating centre at the Nuffield Department of Population Health, University of Oxford. A more detailed statistical analysis plan will be developed by the investigators and published on the study website prior to any analyses of aggregated unblinded data being conducted.

##### 3.1.1 Outcomes

For each pairwise comparison with the 'no additional treatment' arm, the **primary objective** is to provide reliable estimates of the effect of study treatments on all-cause mortality at 28 days after randomisation (with subsidiary analyses of cause of death and of death at various timepoints following discharge).

The **secondary objectives** are to assess the effects of study treatments on duration of hospital stay; the need for (and duration of) ventilation; and the need for renal replacement therapy.

Data from routine healthcare records (including linkage to medical databases held by organisations such as NHS Digital) and from relevant research studies (such as UK Biobank and Genomics England) will allow subsidiary analyses of the effect of the study treatments on particular non-fatal events (e.g. ascertained through linkage to Hospital Episode Statistics), the influence of pre-existing major co-morbidity (e.g. diabetes, heart disease, lung disease, hepatic insufficiency, severe depression, severe kidney impairment, immunosuppression), and longer-term outcomes (e.g. 6 month survival) as well as in particular sub-categories of patient (e.g. by genotype, pregnancy).

##### 3.1.2 Methods of analysis

Comparisons will be made between all participants randomised to the different treatment arms, irrespective of whether they received their allocated treatment ("intention-to-treat" analyses).

For time-to-event analyses, each treatment group will be compared with the no additional treatment group using the log-rank test. Kaplan-Meier estimates for the time to event will also be plotted (with associated log-rank p-values). The log-rank 'observed minus expected' statistic (and its variance) will also be used to estimate the average event rate ratio (and its confidence interval) for those allocated to each treatment group versus the no additional treatment group. For binary outcomes where the timing is unknown, the odds ratio and absolute risk difference will be calculated with confidence intervals and p-value reported. For the primary outcome, death within 28 days of randomisation, discharge alive before 28 days will assume safety from the event (in the absence of additional data confirming otherwise).

Pairwise comparisons will be made between each treatment arm and the no additional treatment arm (reference group). However, since not all treatments may be available or suitable for all patients, those in the no additional treatment arm will only be included in a given comparison if, at the point of their randomisation, they *could* alternatively have been randomised to the active treatment of interest. Adjustment for multiple treatment comparisions due to the multi-arm design will be made using the Dunnett test. All p-values will be 2-sided.

Pre-specified subgroup analysis will be conducted for the primary outcome using the statistical test for interaction (or test for trend where appropriate) for the following: disease severity; time since onset of symptoms; sex; age group).

Further details will be fully described in the Statistical Analysis Plan.

### 4 DATA AND SAFETY MONITORING

##### 4.1.1 Recording Suspected Serious Adverse Reactions

The focus is on those events that, based on a single case, are highly likely to be related to the study medication. Examples include anaphylaxis, Stevens Johnson Syndrome, or bone marrow failure, where there is no other plausible explanation.

Any Serious Adverse Event[a] that is believed with a reasonable probability to be due to one of the study treatments will be considered a Suspected Serious Adverse Reaction. In making this assessment, there should be consideration of the probability of an alternative cause (for example, COVID-19 itself or some other condition preceding randomisation), the timing of the event with respect to study treatment, the response to withdrawal of the study treatment, and (where appropriate) the response to subsequent re-challenge.

All Suspected Serious Adverse Reactions should be reported by telephone to the Central Coordinating Office and recorded on the study IT system immediately.

##### 4.1.2 Central assessment and onward reporting of SUSARs

Clinicians at the Central Coordinating Office are responsible for expedited review of reports of SSARs received. Additional information (including the reason for considering it both serious and related, and relevant medical and medication history) will be sought.

The focus of SUSAR reporting will be on those events that, based on a single case, are highly likely to be related to the study medication. To this end, anticipated events that are either efficacy endpoints, consequences of the underlying disease, or common in the study

[a] Serious Adverse Events are defined as those adverse events that result in death; are life-threatening; require in-patient hospitalisation or prolongation of existing hospitalisation; result in persistent or significant disability or incapacity; result in congenital anomaly or birth defect; or are important medical events in the opinion of the responsible investigator (that is, not life-threatening or resulting in hospitalisation, but may jeopardise the participant or require intervention to prevent one or other of the outcomes listed above).

population will be exempted from expedited reporting. Thus the following events will be exempted from expedited reporting:

(i) Events which are the consequence of COVID-19; and
(ii) Common events which are the consequence of conditions preceding randomisation.

Any SSARs that are not exempt will be reviewed by a Central Coordinating Office clinician and an assessment made of whether the event is "expected" or not (assessed against the relevant Summary of Product Characteristics or Investigator Brochure). Any SSARs that are not expected would be considered a Suspected Unexpected Serious Adverse Reaction (SUSAR).

All confirmed SUSARs will be reported to the Chair of the DMC and to relevant regulatory authorities, ethics committees, and investigators in an expedited manner in accordance with regulatory requirements.

##### 4.1.3 Recording other Adverse Events

In addition to recording Suspected Serious Adverse Reactions (see section 4.1.1), information will be collected on all deaths and efforts will be made to ascertain the underlying cause. Other serious or non-serious adverse events will not be recorded. It is anticipated that for some sub-studies, more detailed information on adverse events (e.g. through linkage to medical databases) or on other effects of the treatment (e.g. laboratory or radiological features) will be recorded and analysed but this is not a requirement of the core protocol.

##### 4.1.4 Role of the Data Monitoring Committee (DMC)

During the study, interim analyses of all study data will be supplied in strict confidence to the independent DMC. The DMC will request such analyses at a frequency relevant to the emerging data from this and other studies.

The DMC will independently evaluate these analyses and any other information considered relevant. The DMC will determine if, in their view, the randomised comparisons in the study have provided evidence on mortality that is strong enough (with a range of uncertainty around the results that is narrow enough) to affect national and global treatment strategies. In such a circumstance, the DMC will inform the Trial Steering Committee who will make the results available to the public and amend the trial arms accordingly. Unless this happens, the Steering Committee, Chief Investigator, study staff, investigators, study participants, funders and other partners will remain blind to the interim results until 28 days after the last patient has been randomised for a particular intervention arm (at which point analyses may be conducted comparing that arm with the no additional treatment arm).

##### 4.1.5 Blinding

This is an open-label study. However, while the study is in progress, access to tabular results by allocated treatment allocation will not be available to the research team, patients, or members of the Steering Committee (unless the DMC advises otherwise).

### 5 QUALITY MANAGEMENT

##### 5.1.1 Quality By Design Principles

In accordance with the principles of Good Clinical Practice and the recommendations and guidelines issued by regulatory agencies, the design, conduct and analysis of this trial is focussed on issues that might have a material impact on the wellbeing and safety of study participants (hospitalised patients with suspected or confirmed SARS-CoV-2 infection) and the reliability of the results that would inform the care for future patients.

The critical factors that influence the ability to deliver these quality objectives are:

- to minimise the burden on busy clinicians working in an overstretched hospital during a major epidemic
- to ensure that suitable patients have access to the trial medication without impacting or delaying other aspects of their emergency care
- to provide information on the study to patients and clinicians in a timely and readily digestible fashion but without impacting adversely on other aspects of the trial or the patient's care
- to allow individual clinicians to use their judgement about whether any of the treatment arms are not suitable for the patient
- to collect comprehensive information on the mortality and disease status

In assessing any risks to patient safety and well-being, a key principle is that of proportionality. Risks associated with participation in the trial must be considered in the context of usual care. At present, there are no proven treatments for COVID-19, basic hospital care (staffing, beds, ventilatory support) may well be overstretched, and mortality for hospitalised patients may be around 10% (or more in those who are older or have significant co-morbidity).

##### 5.1.2 Training and monitoring

The focus will be on those factors that are critical to quality (i.e. the safety of the participants and the reliability of the trial results). Remedial actions would focus on issues with the potential to have a substantial impact on the safety of the study participants or the reliability of the results.

The study will be conducted in accordance with the principles of International Conference on Harmonisation Guidelines for Good Clinical Research Practice (ICH-GCP) and relevant local, national and international regulations. Any serious breach of GCP in the conduct of the clinical trial will be handled in accordance with regulatory requirements. Prior to initiation of the study at each Local Clinical Centre (LCC), the Central Coordinating Office (CCO) will confirm that the LCC has adequate facilities and resources to carry out the study. LCC lead investigators and study staff will be provided with training materials.

In the context of this epidemic, visits to hospital sites is generally not appropriate as they could increase the risks of spreading infection, and in the context of this trial they generally would not influence the reliability of the trial results or the well-being of the participants. In exceptional circumstances, the CCO may arrange monitoring visits to LCCs as considered appropriate based on perceived training needs and the results of central statistical monitoring of study data.[6,7] The purpose of such visits will be to ensure that the study is being conducted in accordance with the protocol, to help LCC staff to resolve any local problems, and to provide extra training focussed on specific needs. No routine source data verification will take place.

##### 5.1.3 Data management

LCC clinic staff will use the bespoke study web-based applications for study management and to record participant data (including case report forms) in accordance with the protocol. Data will be held in central databases located at the CCO or on secure cloud servers. In some circumstances (e.g. where there is difficulty accessing the internet or necessary IT equipment), paper case report forms may be required with subsequent data entry by either LCC or CCO staff. Although data entry should be mindful of the desire to maintain integrity and audit trails, in the circumstances of this epidemic, the priority is on the timely entry of data that is sufficient to support reliable analysis and interpretation about treatment effects. CCO staff will use be responsible for provision of the relevant web-based applications and for generation of data extracts for analyses.

All data access will be controlled by unique usernames and passwords, and any changes to data will require the user to enter their username and password as an electronic signature in accordance with regulatory requirements.[8] Staff will have access restricted to the functionality and data that are appropriate for their role in the study.

##### 5.1.4 Source documents and archiving

Source documents for the study constitute the records held in the study main database. These will be retained for at least 25 years from the completion of the study. Identifiable data will be retained only for so long as it is required to maintain linkage with routine data sources (see section 2.1.7). The sponsor and regulatory agencies will have the right to conduct confidential audits of such records in the CCO and LCCs (but should mindful of the workload facing participating hospitals and the infection control requirements during this epidemic).

#### 6 OPERATIONAL AND ADMINISTRATIVE DETAILS

##### 6.1.1 Sponsor and coordination

The University of Oxford will act as the trial Sponsor. The trial will be coordinated by a Central Coordinating Office within the Nuffield Department of Population Health staffed by members of the two registered clinical trials units – the Clinical Trial Service Unit and the National Perinatal Epidemiology Unit Clinical Trials Unit. The data will be collected, analysed and published independently of the source of funding.

##### 6.1.2 Funding

This study is supported by a grant to the University of Oxford from UK Research and Innovation/National Institute for Health Research (NIHR) and by core funding provided by NIHR Oxford Biomedical Research Centre, the Wellcome Trust, the Bill and Melinda Gates Foundation, Health Data Research UK, and the Medical Research Council Population Health Research Unit, and NIHR Clinical Trials Unit Support Funding.

##### 6.1.3 Indemnity

The University has a specialist insurance policy in place which would operate in the event of any participant suffering harm as a result of their involvement in the research (Newline Underwriting Management Ltd, at Lloyd's of London). NHS indemnity operates in respect of the clinical treatment that is provided.

##### 6.1.4 Local Clinical Centres

The study will be conducted at multiple hospitals (Local Clinical Centres) within the UK. At each LCC, a lead investigator will be responsible for trial activities but much of the work will be carried out by medical staff attending patients with COVID-19 within the hospital and by hospital research nurses, medical students and other staff with appropriate education, training, and experience.

##### 6.1.5 Supply of study treatments

For licensed treatments (e.g. Lopinavir-Ritonavir, corticosteroids) all aspects of treatment supply, storage, and management will be in accordance with standard local policy and practice for prescription medications. Treatment issue to randomised participants will be by prescription.

For unlicensed treatments, manufacture, packaging and delivery will be the responsibility of the pharmaceutical company and Department of Health and Social Care. Treatment issue to randomised participants will be in accordance with local practice (and may be in line with the processes required for routine prescriptions or compassionate use).

Study treatments will not be labelled beyond other than as required for routine clinical use. They will be stored alongside other routine medications with no additional monitoring. No accountability records will be kept beyond those used for routine prescriptions.

##### 6.1.6 End of trial

The end of the scheduled treatment phase is defined as the date of the last follow-up visit of the last participant. In the UK, it is intended to extend follow-up for a year or more beyond the final study visit through linkage to routine medical records and central medical databases. The end of the study is the date of the final data extraction from NHS Digital (anticipated to be 10 years after the last patient is enrolled).

##### 6.1.7 Publications and reports

The Steering Committee will be responsible for drafting the main reports from the study and for review of any other reports. In general, papers initiated by the Steering Committee (including the primary manuscript) will be written in the name of the RECOVERY Collaborative Group, with individual investigators named personally at the end of the report (or, to comply with journal requirements, in web-based material posted with the report).

The Steering Committee will also establish a process by which proposals for additional publications (including from independent external researchers) are considered by the Steering Committee. The Steering Committee will facilitate the use of the study data and approval will not be unreasonably withheld. However, the Steering Committee will need to be satisfied that any proposed publication is of high quality, honours the commitments made to the study participants in the consent documentation and ethical approvals, and is compliant with relevant legal and regulatory requirements (e.g. relating to data protection and privacy). The Steering Committee will have the right to review and comment on any draft manuscripts prior to publication.

##### 6.1.8 Substudies

Proposals for substudies must be approved by the Steering Committee and by the relevant ethics committee and competent authorities (where required) as a substantial amendment or separate study before they begin. In considering such proposals, the Steering Committee will need to be satisfied that the proposed substudy is worthwhile and will not compromise the main study in any way (e.g. by impairing recruitment or the ability of the participating hospitals to provide care to all patients under their care).

### 8 VERSION HISTORY

| Version number | Date | Brief Description of Changes |
|---|---|---|
| 1.0 | 13-Mar-2020 | Initial version |
| 2.0 | 21-Mar-2020 | Addition of hydroxychloroquine. Administrative changes and other clarifications. |
| 3.0 | 07-Apr-2020 | Extension of eligibility to those with suspected COVID-19<br>Addition of azithromycin arm..<br>Addition of inclusion of adults who lack permanently lack capacity.<br>Change to primary outcome from in-hospital death to death within 28 days of randomization. |

### 9 APPENDICES

##### 9.1.1 Appendix 1: Information about the treatment arms

All patients will receive usual care in the participating hospital.

**No additional treatment:** There are no proven therapies for COVID-19.

**Lopinavir-Ritonavir:** Lopinavir is a human immunodeficiency virus 1 (HIV-1) protease inhibitor, which is combined with ritonavir to increase lopinavir's plasma half-life. It is licensed in adults and children from the age of 14 days (2 years in Scotland). It has been widely used in pregnant women.[1] Lopinavir has in vitro inhibitory activity against SARS coronavirus (SARS-CoV) and MERS-CoV.[2-4] [5] In common marmosets infected with MERS-CoV, animals treated with lopinavir/ritonavir had improved clinical, radiological, and pathological outcomes and reduced viral loads compared with untreated animals.[6] In one single-center, open-label study of the addition of lopinavir 400mg/ritonavir 100mg to ribavirin and corticosteroids in SARS patients the risk of adverse clinical outcomes (acute respiratory distress syndrome [ARDS] or death) was significantly lower (2.4% v 28.8%, $p<0.001$) compared to a historical control group.[2]

The most common short-term side effects in adults are diarrhoea, nausea, and vomiting. It must not be used by patients with severe liver disease. It should not be co-administered with medicinal products that are highly dependent on CYP3A for clearance and for which elevated plasma concentrations are associated with serious and/or life-threatening events (see Summary of Product Characteristics). Storage should be as per conditions in the Summary of Product Characteristics.

**Dexamethasone:** Favourable modulation of the immune response is considered one of the possible mechanisms by which corticosteroids might be beneficial in the treatment of severe acute respiratory coronavirus infections, including COVID-19, SARS and MERS. Common to severe cases of these infections is the presence of hypercytokinemia (a cytokine 'storm') and development of acute lung injury or adult respiratory distress syndrome (ARDS).[7-10] Pathologically, diffuse alveolar damage is found in patients who die from these infections.[11] A growing volume of clinical trial data from patients with severe community acquired pneumonia, ARDS and septic shock suggest benefit from low-to-moderate dose corticosteroids in relation to mortality and length of stay.[12-14]

In trials of low-to-moderate doses of corticosteroids, the main adverse effect has been hyperglycaemia.[13,15] A systematic review of (mainly low-dose) corticosteroid trials in severe sepsis and septic shock did not identify any increased risk of gastroduodenal bleeding, superinfection or neuromuscular weakness; an association with an increased risk of hyperglycaemia (RR 1.16, 95% CI 1.07 to 1.25) and hypernatraemia (RR 1.61, 95% CI 1.26 to 2.06) was noted.[16] Dexamethasone has a) minimal mineralocorticoid activity and does not affect sodium and water balance, thus avoiding potential problems with fluid retention which are not uncommon in severe viral pneumonitis/ARDS, and b) a comparatively long biological half-life of 36 to 54 hours enabling once a day dosing. In pregnancy, prednisolone 40 mg administered by mouth (or intravenous hydrocortisone 80 mg twice daily) should be used instead of dexamethasone. Storage should be as per conditions in the Summary of Product Characteristics.

**Hydroxycholoroquine:** Chloroquine (CQ), an antimalarial drug discovered in 1934 and introduced generally in 1947, is the drug to which humans have been most exposed, with an annual global consumption of hundreds of metric tonnes for over 50 years. It is inexpensive, simple to administer, and, at the appropriate doses, has an excellent safety profile in all age groups and has been the prophylactic drug of choice in pregnancy [17]. In addition to its antimalarial use both chloroquine and the closely related hydroxychloroquine (HCQ) are used in continuous daily dosing for rheumatoid arthritis, systemic and discoid lupus erythematosus and psoriatic arthritis. HCQ is reported to have better safety profile than CQ, better gastrointestinal tolerability, and less retinal toxicity [18].

CQ has significant antiviral activity against SARS-CoV-2 in cell culture ($EC_{50}$ = 1.13 μM; $CC_{50}$ > 100 μM, SI > 88.50), as it does for the related SARS-CoV-1 [19-22]. CQ blocks virus infection by increasing endosomal pH required for virus/ cell fusion, as well as interfering with the glycosylation of cellular receptors of SARS-CoV.[21] In SARS-CoV-2 infected Vero cells, HCQ ($EC_{50}$=0.72 μM) has been reported to be more potent than CQ ($EC_{50}$=5.47 μM) [23], although Liu et al reported that CQ was more potent than HCQ.[24] These are relatively high levels by comparison with therapeutic exposures in the treatment of malaria but could be achieved with daily oral dosing. Chloroquine has complex pharmacokinetic properties and although the relationship between plasma concentrations and concentrations in respiratory epithelium is not known precisely, in rats the concentration in lung is between 124 and 748-fold that in plasma [25]. If active, HCQ concentrations in the human lung would be expected to exceed those required for the $EC_{90}$ after an initial dose. There are preliminary reports emerging from China and France of clinical benefit in the treatment of COVID-19 infections [26,27].

The recommended adult dosing of chloroquine for treatment of non-falciparum malaria (BNF) is: Initially 620 mg, then 310 mg after 6-8 hours, then 310 mg daily for 2 days. This is equivalent to 930mg base in first 24 hours. This is a loading dose to ensure the necessary blood concentrations are achieved rapidly.

Hydroxychloroquine is very similar to chloroquine. It is used mainly to treat rheumatoid arthritis and other related conditions. The adult dose is usually 400-600mg per day (equivalent to 310 to 465 mg base). Sometimes 800mg per day is given.

The dose in RECOVERY is Hydroyxchloroquine (155mg base per 200 mg tablet):

| | |
|---|---|
| Initial dose: | 4 tablets |
| 6 hours after initial dose: | 4 tablets |
| 12 hours after initial dose: | 2 tablets |
| 24 hours after initial dose: | 2 tablets |
| Thereafter: | 2 tablets every 12 hours for a total of 10 days |

12x155mg = 1860mg base = in first 24 hours

So the loading dose in RECOVERY is twice the normal dose for treating malaria. However, this dose has been selected based on the available data of the $IC_{50}$ for SARS-

CoV-2. The objective is to reach plasma concentrations that are inhibitory to the virus as soon as safely possible. The plasma concentrations that will result are at the higher end of those encountered during steady state treatment of rheumatoid arthritis. Given the significant mortality in patients hospitalised with COVID-19, this dose is felt to be justified. This is the schedule that has been adopted by the World Health Organisation. No dose adjustment is required for weight based on the doses defined in this protocol.

**Azithromycin:** Azithromycin is a macrolide antibiotic. In addition to their antimicrobial properties, the macrolide antibiotics are known to have immunomodulatory activity. The mechanism of immunomodulation includes decreased production of pro-inflammatory cytokines and inhibition of neutrophil activation.[28-30] Macrolides are widely used both in infectious pneumonia due to their antimicrobial activity and in chronic inflammatory lung disease due to the immunomodulatory effects.[31] Azithromycin is preferred over other macrolides because data suggest it has stronger immunomodulatory effects than other macrolides.[30]

The use of macrolides in influenza-associated pneumonia has been associated with a faster reduction in inflammatory cytokines and, in combination with naproxen, decreased mortality.[32-34] Observational studies in MERS-CoV have not demonstrated a mortality benefit of macrolide use.[35] Macrolides have not been evaluated in severe betacoronavirus infections in randomised controlled trials. The safety of macrolides is well established.

##### 9.1.2 Appendix 2: Drug specific contraindications and cautions

**Lopinavir/ritonavir**

- Severe hepatic insufficiency*
- Co-administration with medicinal products that are highly dependent on CYP3A for clearance and for which elevated plasma concentrations are associated with serious and/or life-threatening events. This includes alfuzosin, ranolazine, amiodarone, dronaderone, fusidic acid, neratinib, venetoclax, colchicine, astemizole, terfenadine, lurasidone, pimozide, quetiapine, dihydroergotamine, ergonovine, ergotamine, methylergonovine, cisapride, elbasvir/grazoprevir, ombitasvir/paritaprevir/ritonavir, lovastatin, simvastatin, lomitapide, avanafil, sildenafil, vardenafil, midazolam, triazolam (See Summary of Product Characteristics for more detail). It may be appropriate to temporarily withhold such concomitant medication while the patient is receiving lopinavir/ritonavir.

**Dexamethasone**

- Known contra-indication to short-term dexamethasone.

**Hydroxychloroquine**

- Known prolonged QTc interval
- Caution: Co-administration with medications that prolong the QT interval (e.g. macrolides, quinolones) is not an absolute contraindication, but it may be appropriate to check the QT interval by performing an ECG.

**Azithromycin**

- Known prolonged QTc interval
- Co-administration with chloroquine or hydroxychloroquine
- Known hypersensitivity to macrolide antibiotic

* If these conditions are recorded on the baseline case report form, patients will be ineligible for randomisation to that arm of the study.

Note: This study is being conducted within hospitals. Therefore use of medication will be subject to standard medication reviews (typically within 48 hours of enrolment) which will guide modifications to both the study treatment and use of concomitant medication (e.g. in the case of potential drug interactions). The doctor may decide whether it is appropriate to stop such medications temporarily to allow the patient to complete the course of their assigned intervention.

##### 9.1.3 Appendix 3: Organisational Structure and Responsibilities

**Chief Investigator**
The Chief Investigator has overall responsibility for:

(i) Design and conduct of the Study in collaboration with the Steering Committee;
(ii) Preparation of the Protocol and subsequent revisions;

**Steering Committee**
The Steering Committee (see Section 9.1.4 for list of members) is responsible for:

(i) Agreement of the final Protocol and the Data Analysis Plans;
(ii) Reviewing progress of the study and, if necessary, deciding on Protocol changes;
(iii) Review and approval of study publications and substudy proposals;
(iv) Reviewing new studies that may be of relevance.

**Data Monitoring Committee**
The independent Data Monitoring Committee is responsible for:

(i) Reviewing unblinded interim analyses according to the Protocol;
(ii) Advising the Steering Committee if, in their view, the randomised data provide evidence that may warrant a change in the protocol (e.g. modification or cessation of one or more of the treatment arms).

**Central Coordinating Office (CCO)**
The CCO is responsible for the overall coordination of the Study, including:

(i) Study planning and organisation of Steering Committee meetings;
(ii) Ensuring necessary regulatory and ethics committee approvals;
(iii) Development of Standard Operating Procedures and computer systems
(iv) Monitoring overall progress of the study;
(v) Provision of study materials to LCCs;
(vi) Monitoring and reporting safety information in line with the protocol and regulatory requirements;
(vii) Dealing with technical, medical and administrative queries from LCCs.

**Local Clinical Centres (LCC)**
The LCC lead investigator and LCC clinic staff are responsible for:

(i) Obtaining all relevant local permissions (assisted by the CCO)
(ii) All trial activities at the LCC, including appropriate training and supervision for clinical staff
(iii) Conducting trial procedures at the LCC in line with all relevant local policies and procedures;
(iv) Dealing with enquiries from participants and others.

##### 9.1.4 Appendix 4: Organisational Details

**STEERING COMMITTEE**
(Major organisational and policy decisions, and scientific advice; blinded to treatment allocation)

| | |
|---|---|
| Chief Investigator | Peter Horby |
| Deputy Chief Investigator | Martin Landray |
| Clinical Trial Unit Leads | Richard Haynes, Edmund Juszczak |
| Co-investigators | Kenneth Baillie (Scotland Lead), Thomas Jaki, Katie Jeffery, Wei Shen Lim, Alan Montgomery, Kathy Rowan |

**DATA MONITORING COMMITTEE**
(Interim analyses and response to specific concerns)

| | |
|---|---|
| Chair | Peter Sandercock |
| Members | Janet Darbyshire, David DeMets, Robert Fowler, David Kalloo, Ian Roberts, Janet Wittes |
| Statistician (non-voting) | Jonathan Emberson, Natalie Staplin |

**To enquire about the trial, contact the RECOVERY Central Coordinating Office**

RECOVERY Central Coordinating Office:
Richard Doll Building
Old Road Campus
Roosevelt Drive
Oxford OX3 7LF
United Kingdom

Website: www.recoverytrial.net
(copies of this protocol and related forms and information can be downloaded)

**To RANDOMISE a patient, visit:**

RECOVERY
Randomised Evaluation of COVID-19 Therapy

Website: www.recoverytrial.net

### RANDOMISED EVALUATION OF COVID-19 THERAPY (RECOVERY)

**Background:** In early 2020, as this protocol was being developed, there were no approved treatments for COVID-19, a disease induced by the novel coronavirus SARS-CoV-2 that emerged in China in late 2019. The UK New and Emerging Respiratory Virus Threats Advisory Group (NERVTAG) advised that several possible treatments should be evaluated, including Lopinavir-Ritonavir, low-dose corticosteroids, and Hydroxychloroquine (which has now been done). A World Health Organization (WHO) expert group issued broadly similar advice. These groups also advised that other treatments will soon emerge that require evaluation.

**Eligibility and randomisation:** This protocol describes a randomised trial among patients hospitalised for COVID-19. All eligible patients are randomly allocated between several treatment arms, each to be given in addition to the usual standard of care in the participating hospital: No additional treatment *vs* azithromycin *vs* corticosteroids (children only) *vs* intravenous immunoglobulin (children only). In a factorial design (in the UK alone), eligible patients are allocated simultaneously to no additional treatment *vs* convalescent plasma *vs* synthetic neutralising antibodies (REGN-COV2). Separately, all participants aged 18 years or older will be allocated to either aspirin *vs* control. The study allows a subsequent randomisation for patients with progressive COVID-19 (evidence of hyper-inflammatory state): No additional treatment *vs* tocilizumab. For patients for whom not all the trial arms are appropriate or at locations where not all are available, randomisation will be between fewer arms.

**Adaptive design:** The interim trial results will be monitored by an independent Data Monitoring Committee (DMC). The most important task for the DMC will be to assess whether the randomised comparisons in the study have provided evidence on mortality that is strong enough (with a range of uncertainty around the results that is narrow enough) to affect national and global treatment strategies. In such a circumstance, the DMC will inform the Trial Steering Committee who will make the results available to the public and amend the trial arms accordingly. Regardless, follow-up will continue for all randomised participants, including those previously assigned to trial arms that are modified or ceased. New trial arms can be added as evidence emerges that other candidate therapeutics should be evaluated.

**Outcomes:** The main outcomes will be death, discharge, need for ventilation and need for renal replacement therapy. For the main analyses, follow-up will be censored at 28 days after randomisation. Additional information on longer term outcomes may be collected through review of medical records or linkage to medical databases where available (such as those managed by NHS Digital and equivalent organisations in the devolved nations).

**Simplicity of procedures:** To facilitate collaboration, even in hospitals that suddenly become overloaded, patient enrolment (via the internet) and all other trial procedures are greatly streamlined. Informed consent is simple and data entry is minimal. Randomisation via the internet is simple and quick, at the end of which the allocated treatment is displayed on the screen and can be printed or downloaded. Follow-up information is recorded at a single timepoint and may be ascertained by contacting participants in person, by phone or electronically, or by review of medical records and databases.

**Data to be recorded:** At randomisation, information will be collected on the identity of the randomising clinician and of the patient, age, sex, major co-morbidity, pregnancy, COVID-19 onset date and severity, and any contraindications to the study treatments. The main outcomes will be death (with date and probable cause), discharge (with date), need for ventilation (with number of days recorded) and need for renal replacement therapy. Reminders will be sent if outcome data have not been recorded by 28 days after randomisation. Suspected Unexpected Serious Adverse Reactions (SUSARs) to one of the study medications (e.g., Stevens-Johnson syndrome, anaphylaxis, aplastic anaemia) will be collected and reported in an expedited fashion. Other adverse events will not be recorded but may be available through linkage to medical databases.

**Numbers to be randomised:** The larger the number randomised the more accurate the results will be, but the numbers that can be randomised will depend critically on how large the epidemic becomes. If substantial numbers are hospitalised in the participating centres then it may be possible to randomise several thousand with mild disease and a few thousand with severe disease, but realistic, appropriate sample sizes could not be estimated at the start of the trial.

**Heterogeneity between populations:** If sufficient numbers are studied, it may be possible to generate reliable evidence in certain patient groups (e.g. those with major co-morbidity or who are older). To this end, data from this study may be combined with data from other trials of treatments for COVID-19, such as those being planned by the WHO.

**Add-on studies:** Particular countries or groups of hospitals, may well want to collaborate in adding further measurements or observations, such as serial virology, serial blood gases or chemistry, serial lung imaging, or serial documentation of other aspects of disease status. While well-organised additional research studies of the natural history of the disease or of the effects of the trial treatments could well be valuable (although the lack of placebo control may bias the assessment of subjective side-effects, such as gastro-intestinal problems), they are not core requirements.

**To enquire about the trial, contact the RECOVERY Central Coordinating Office**

Nuffield Department of Population Health, Richard Doll Building, Old Road Campus, Roosevelt Drive, Oxford OX3 7LF, United Kingdom

Website: www.recoverytrial.net
**To enquire about the trial outside of the UK, contact the relevant Clinical Trial Units** (see section 10)

**To RANDOMISE a patient, visit:**

Website: www.recoverytrial.net

### 1 BACKGROUND AND RATIONALE

#### 1.1 Setting

In 2019 a novel coronavirus-induced disease (COVID-19) emerged in Wuhan, China. A month later the Chinese Center for Disease Control and Prevention identified a new beta-coronavirus (SARS coronavirus 2, or SARS-CoV-2) as the aetiological agent.[1] The clinical manifestations of COVID-19 range from asymptomatic infection or mild, transient symptoms to severe viral pneumonia with respiratory failure. As many patients do not progress to severe disease the overall case fatality rate per infected individual is low, but hospitals in areas with significant community transmission have experienced a major increase in the number of hospitalised pneumonia patients, and the frequency of severe disease in hospitalised patients can be as high as 30%.[2-4] The progression from prodrome (usually fever, fatigue and cough) to severe pneumonia requiring oxygen support or mechanical ventilation often takes one to two weeks after the onset of symptoms.[2] The kinetics of viral replication in the respiratory tract are not well characterized, but this relatively slow progression provides a potential time window in which antiviral therapies could influence the course of disease. In May 2020 a new COVID-associated inflammatory syndrome in children was identified, Paediatric Inflammatory Multisystem Syndrome - Temporally associated with SARS-CoV-2 (PIMS-TS).[5] A rapid NHS England-led consensus process identified the need to evaluate corticosteroids and intravenous immunoglobulin (IVIg) as initial therapies in PIMS-TS, and confirmed tocilizumab as one of the biological anti-inflammatory agents to be evaluated as a second line therapy.

#### 1.2 Treatment Options

##### 1.2.1 Main randomisation

This protocol allows reliable assessment of the effects of multiple different treatments (including re-purposed and novel drugs) on major outcomes in COVID-19. All patients will receive usual care for the participating hospital.

**Randomisation part A:** Eligible patients may be randomly allocated between the following treatment arms:

- **No additional treatment**
- **Azithromycin**
- **Corticosteroids (children ≤44 weeks gestational age, or >44 weeks gestational age with PIMS-TS only)**
- **Intravenous immunoglobulin (children >44 weeks gestational age with PIMS-TS only)**

**Randomisation part B [UK only]:** Simultaneously, eligible patients will be randomly allocated between the following treatment arms:

- **No additional treatment**
- **Convalescent plasma**
- **Synthetic neutralising antibodies (REGN-COV2) (adults and children ≥12 years old only)**

**Randomisation part C (adults ≥18 years old only):** Simultaneously, eligible patients will be randomly allocated between the following treatment arms:

- **No additional treatment**
- **Aspirin**

##### 1.2.2 Second randomisation for patients with progressive COVID-19

Severe COVID-19 is associated with release of pro-inflammatory cytokines, such as IL-1, IL-6 and TNFα, and other markers of systemic inflammation including ferritin and C-reactive protein.[3,6,7] There is a possibility that this response may cause or exacerbate lung injury, leading to life-threatening disease.

Participants with progressive COVID-19 (as evidenced by hypoxia and an inflammatory state) may undergo an optional second randomisation between the following treatment arms:

- **No additional treatment**
- **Tocilizumab**

##### 1.2.3 Modifications to the number of treatment arms

Other arms can be added to the first or second randomisation if evidence emerges that there are suitable candidate therapeutics. Conversely, in some patient populations, not all trial arms are appropriate (e.g. due to contraindications based on co-morbid conditions or concomitant medication); in some hospitals or countries, not all treatment arms will be available (e.g. due to manufacturing and supply shortages); and at some times, not all treatment arms will be active (e.g. due to lack of relevant approvals and contractual agreements). The Trial Steering Committee may elect to pause one or more of the arms in order to increase trial efficiency during a fluctuating epidemic. In any of these situations, randomisation will be between fewer arms. Depending on the availability and suitability of treatments, it may be allowed for participants to be randomised in only one or two parts (A, B, or C) of the main randomisation.

#### 1.3 Design Considerations

The RECOVERY Protocol describes an overarching trial design to provide reliable evidence on the efficacy of candidate therapies for suspected or confirmed COVID-19 infection in hospitalised patients receiving usual standard of care.

In early 2020, when the trial first started, there were no known treatments for COVID-19. The anticipated scale of the epidemic is such that hospitals, and particularly intensive care facilities, may be massively overstretched at some points in time, with around 10% requiring hospitalisation. In this situation, even treatments with only a moderate impact on survival or on hospital resources could be worthwhile. Therefore, the focus of RECOVERY is the impact of candidate treatments on mortality and on the need for hospitalisation or ventilation.

Critically, the trial is designed to minimise the burden on front-line hospital staff working within an overstretched care system during a major epidemic. Eligibility criteria are therefore simple and trial processes (including paperwork) are minimised.

The protocol is deliberately flexible so that it is suitable for a wide range of settings, allowing:

- a broad range of patients to be enrolled in large numbers;
- randomisation between only those treatment arms that are *both* available at the hospital *and* not believed by the enrolling doctor to be contraindicated (e.g. by particular co-morbid conditions or concomitant medications);
- treatment arms to be added or removed according to the emerging evidence; and
- additional substudies may be added to provide more detailed information on side effects or sub-categorisation of patient types but these are not the primary objective and are not required for participation.

In a cohort of 191 hospitalised COVID-19 patients with a completed outcome, the median time from illness onset to discharge was 22·0 days (IQR 18·0–25·0) and the median time to death was 18·5 days (15·0–22·0). Thirty-two patients (17%) required invasive mechanical ventilation and the median time from onset to mechanical ventilation was 14.5 days. Therefore, early endpoint assessment, such as 28 days after randomisation, is likely to provide largely complete outcome data and will permit early assessment of treatment efficacy and safety.[8]

#### 1.4 Potential for effective treatments to become available

In early 2020, when the trial first started, there were no known treatments for COVID-19. However, over time, effective treatments may become available, typically as the result of reliable information from randomised trials (including from this study). For example, in June 2020, results from the RECOVERY trial showed that dexamethasone reduces the mortality in COVID-19 patients requiring mechanical ventilation or oxygen. In response, many clinical guidelines now recommend the use of dexamethasone as standard of care for these types of patients.

The RECOVERY trial randomises eligible participant to usual standard of care for the local hospital alone vs usual standard of care plus one or more additional study treatments. Over time, it is expected that usual standard of care alone will evolve. Thus randomisation will always be relevant to the current clinical situation and the incremental effects of the study treatments will be appropriately assessed.

### 2 DESIGN AND PROCEDURES

#### 2.1 Eligibility

Patients are eligible for the study if all of the following are true:

**(i) Hospitalised**

**(ii) SARS-CoV-2 infection associated disease (clinically suspected or laboratory confirmed)**

In general, SARS-CoV-2 infection should be suspected when a patient presents with:

a) typical symptoms (e.g. influenza-like illness with fever and muscle pain, or respiratory illness with cough and shortness of breath); and
b) compatible chest X-ray findings (consolidation or ground-glass shadowing); and
c) alternative causes have been considered unlikely or excluded (e.g. heart failure, influenza).

However, the diagnosis remains a clinical one based on the opinion of the managing doctor.

A small number of children (aged <18 years) present with atypical features, including a hyperinflammatory state and evidence of single or multi-organ dysfunction (called Paediatric Multisystem Inflammatory Syndrome temporally associated with COVID-19 [PIMS-TS]). Some do not have significant lung involvement.[a]

**(iii) No medical history that might, in the opinion of the attending clinician, put the patient at significant risk if he/she were to participate in the trial**

In addition, if the attending clinician believes that there is a specific contra-indication to one of the active drug treatment arms (see Appendix 2; section 8.2 and Appendix 3; section 8.3 for children) or that the patient should definitely be receiving one of the active drug treatment arms then that arm will not be available for randomisation for that patient. For patients who lack capacity, an advanced directive or behaviour that clearly indicates that they would not wish to participate in the trial would be considered sufficient reason to exclude them from the trial.

In some locations, children (aged <18 years) will not be recruited, to comply with local and national regulatory approvals (see Section 8.3).

#### 2.2 Consent

Informed consent should be obtained from each patient 16 years and over before enrolment into the study. However, if the patient lacks capacity to give consent due to the severity of their medical condition (e.g. acute respiratory failure or need for immediate ventilation) or

[a] https://www.rcpch.ac.uk/sites/default/files/2020-05/COVID-19-Paediatric-multisystem-%20inflammatory%20syndrome-20200501.pdf

prior disease, then consent may be obtained from a relative acting as the patient's legally designated representative or independent doctor. Further consent will then be sought with the patient if they recover sufficiently. For children aged <16 years old consent will be sought from their parents or legal guardian. Where possible, children aged between 10-15 years old will also be asked for assent. Children aged ≥16 years old will asked for consent as for adults. Witnessed consent may be obtained over the telephone or web video link if hospital visiting rules or parental infection mean a parent/guardian cannot be physically present.

Due to the poor outcomes in COVID-19 patients who require ventilation (>90% mortality in one cohort[8]), patients who lack capacity to consent due to severe disease (e.g. needs ventilation), and for whom a relative to act as the legally designated representative is not immediately available, randomisation and consequent treatment will proceed with consent provided by a treating clinician (independent of the clinician seeking to enrol the patient) who will act as the legally designated representative (if allowed by local regulations). Consent will then be obtained from the patient's personal legally designated representative (or directly from the patient if they recover promptly) at the earliest opportunity.

#### 2.3 Baseline information

The following information will be recorded on the web-based form by the attending clinician or delegate:

- Patient details (e.g. name or initials [depending on privacy requirements], NHS/CHI number [UK only] or medical records number, date of birth, sex)
- Clinician details (e.g. name)
- COVID-19 symptom onset date
- COVID-19 severity as assessed by need for supplemental oxygen, non-invasive ventilation or invasive mechanical ventilation/extracorporeal membrane oxygenation (ECMO)
- Oxygen saturations on air (if available)
- Latest routine measurement of creatinine, C-reactive protein, and D-dimer (if available)
- SARS-CoV-2 PCR test result (if available)
- Major co-morbidity (e.g. heart disease, diabetes, chronic lung disease) and pregnancy
- Use of relevant medications (corticosteroids, remdesivir, antiplatelet and anticoagulant therapy)
- Date of hospitalisation
- Contraindication to the study treatment regimens (in the opinion of the attending clinician)
- Willingness to receive a blood product [UK only]
- Name of person completing the form

The person completing the form will then be asked to confirm that they wish to randomise the patient and will then be required to enter their name and e-mail address.

#### 2.4 Main randomisation

In addition to receiving usual care, eligible patients will be allocated using a central web-based randomisation service (without stratification or minimisation). From version 6.0 of the protocol, a factorial design will be used such that eligible patients may be randomised to one of the treatment arms in Randomisation A and, simultaneously, to one of the treatment arms in Randomisation B. From version 10.0 of the protocol, a further factorial randomisation was added (Main Randomisation part C).

##### 2.4.1 Main randomisation part A:

Eligible patients may be randomised to one of the arms listed below. The doses in this section are for adults. Please see Appendix 3 for paediatric dosing. Study treatments do not need to be continued after discharge from hospital.

- **No additional treatment**

- **Azithromycin 500mg** by mouth (or nasogastric tube) or intravenously once daily for 10 days.

- **Corticosteroid (in children ≤44 weeks gestational age, or >44 weeks gestational age with PIMS-TS only)**: see Appendix 3.

- **Intravenous immunoglobulin (in children >44 weeks gestational age with PIMS-TS only)**: see Appendices 2 and 3 for dose, contraindications and monitoring information.

For randomisation part A, the randomisation program will allocate patients in a ratio of 2:1 between the no additional treatment arm and each of the other arms available. Hence if 5 arms are available, then the randomisation will be in the ratio 2:1:1:1:1. If one or more of the active drug treatments is not available at the hospital or is believed, by the attending clinician, to be contraindicated (or definitely indicated) for the specific patient, then this fact will be recorded via the web-based form prior to randomisation; random allocation will then be between the remaining arms (i.e. in a 2:1:1:1, 2:1:1 or 2:1 ratio). If no treatments are both available and suitable, then it may be possible to only be randomised in part B (UK only) and/or part C.

##### 2.4.2 Main randomisation part B [UK only]:

Eligible patients may be randomised to one of the arms listed below. The doses in this section are for adults. Please see Appendix 3 for paediatric dosing. **Participants in this randomisation should have a serum sample sent to their transfusion laboratory prior to randomisation in which presence of antibodies against SARS-CoV-2 may be tested.**

- **No additional treatment**

- **Convalescent plasma** Single unit of ABO compatible convalescent plasma (275mls +/- 75 mls) intravenous per day on study days 1 (as soon as possible after randomisation) and 2 (with a minimum of 12 hour interval between 1st and 2nd units). ABO identical plasma is preferred if available. The second transfusion should not be given if patient has a suspected serious adverse reaction during or after the first transfusion.

- **Synthetic neutralising antibodies (REGN-COV2; adults and children aged ≥12 years[b] only).** A single dose of REGN10933 + REGN10987 8 g (4 g of each monoclonal antibody) in 250ml 0.9% saline infused intravenously over 60 minutes +/- 15 minutes as soon as possible after randomisation

For randomisation part B, the randomisation program will allocate patients in a ratio of 1:1:1 between each of the arms. If the active treatment is not available at the hospital, the patient does not consent to receive convalescent plasma, or is believed, by the attending clinician, to be contraindicated for the specific patient, then this fact will be recorded via the web-based form and the patient will be excluded from the relevant arm in Randomisation part B.

##### 2.4.3 Main randomisation part C [adults aged ≥18 years only]:

Eligible patients may be randomised to one of the arms listed below.

- **No additional treatment**

- **Aspirin** 150 mg by mouth (or nasogastric tube) or per rectum once daily until discharge.

Note: The allocation in this randomisation should not influence the use of standard thromboprophylaxis care.

#### 2.5 Second randomisation for patients with progressive COVID-19

Patients enrolled in the RECOVERY trial and with clinical evidence of a hyper-inflammatory state may be considered for a second randomisation if they meet the following criteria:

(i) Randomised into the RECOVERY trial no more than 21 days ago
(ii) Clinical evidence of progressive COVID-19:
  a. oxygen saturation <92% on room air or requiring oxygen (or in children (age <18 years), significant systemic disease with persistent pyrexia, with or without evidence of respiratory involvement)[c]; and
  b. C-reactive protein ≥75 mg/L
(iii) No medical history that might, in the opinion of the attending clinician, put the patient at significant risk if he/she were to participate in this aspect of the RECOVERY trial. (Note: Pregnancy and breastfeeding are not specific exclusion criteria.)

Note: Participants may undergo this second randomisation at any point after being first randomised, provided they meet the above criteria, and thus may receive up to four study treatments (one each from Main randomisation parts A, B and C, plus one from the second randomisation). For some participants the second randomisation may be immediately after the first but for others it may occur a few hours or days later, if and when they deteriorate.

---

[b] Older children who weigh <40kg will also not be eligible for this treatment.
[c] A small number of children (age <18 years) present with atypical features, including a hyperinflammatory state and evidence of single or multi-organ dysfunction. Some do not have significant lung involvement.
(see: https://www.rcpch.ac.uk/sites/default/files/2020-05/COVID-19-Paediatric-multisystem-%20inflammatory%20syndrome-20200501.pdf)

The following information will be recorded (on the web-based form) by the attending clinician or delegate:

- Patient details (e.g. name or initials, NHS/CHI number [UK only] or medical records number, date of birth, sex)
- Clinician details (e.g. name)
- COVID-19 severity as assessed by need for supplemental oxygen or ventilation/ECMO
- Markers of progressive COVID-19 (including oxygen saturation, C-reactive protein)
- Contraindication to the study drug treatments (in the opinion of the attending clinician)
- Name of person completing the form

The person completing the form will then be asked to confirm that they wish to randomise the patient and will then be required to enter their own name and e-mail address.

Eligible participants may be randomised between the following treatment arms:

- **No additional treatment**

- **Tocilizumab** by intravenous infusion with the dose determined by body weight:

| Weight* | Dose |
|---|---|
| >40 and ≤65 kg | 400 mg |
| >65 and ≤90 kg | 600 mg |
| >90 kg | 800 mg |

* for lower weights, dosing should be 8 mg/kg (see Appendix 3 for paediatric dosing)
(Note: body weight may be estimated if it is impractical to weigh the patient)

Tocilizumab should be given as a single intravenous infusion over 60 minutes in 100ml sodium chloride 0.9%. A second dose may be given ≥12 and <24 hours later if, in the opinion of the attending clinician, the patient's condition has not improved.

The randomisation program will allocate patients in a ratio of 1:1 between the arms being evaluated in the second randomisation. Participants should receive standard management (including blood tests such as liver function tests and full blood count) according to their clinical need.

#### 2.6 Administration of allocated treatment

The details of the allocated study treatments will be displayed on the screen and can be printed or downloaded. The hospital clinicians are responsible for prescription and administration of the allocated treatments. The patient's own doctors are free to modify or stop study treatments if they feel it is in the best interests of the patient without the need for the patient to withdraw from the study (see section 2.9). This study is being conducted within hospitals. Therefore use of medication will be subject to standard medication reviews (typically within 48 hours of enrolment) which will guide modifications to both the study treatment and use of concomitant medication (e.g. in the case of potential drug interactions).

Note: [UK only] NHS guidelines require patients to have **two** separate blood samples taken for Group and Screen prior to administration of blood products. Each sample is approximately 5 mL and both need to be taken at any time between admission to hospital and receipt of the first plasma transfusion (as the laboratory will not issue plasma without both samples), although if a valid historical sample exists this can be used for one of the samples. The participant's blood group is identified to ensure that blood group-compatible plasma is given and this information would be available to the participant if they wish. Such tests may be required as part of the routine care of the participant if the managing team wish to consider using blood products and samples will be stored, retained and destroyed as per trust standard procedures and protocols. The extra serum sample collected for measurement of coronavirus and antibodies against it will be prepared in the local transfusion laboratory (including removing any identifiers and labelling with the participant's study ID) and sent to a central laboratory for analysis. Once testing is complete these samples will be destroyed.

#### 2.7 Collecting follow-up information

The following information will be ascertained at the time of death or discharge or at 28 days after first randomisation (whichever is sooner):

- Vital status (alive / dead, with date and presumed cause of death, if appropriate)
- Hospitalisation status (inpatient / discharged, with date of discharge, if appropriate)
- SARS-CoV-2 test result
- Use of ventilation (with days of use and type, if appropriate)
- Use of renal dialysis or haemofiltration
- Documented new major cardiac arrhythmia (including atrial and ventricular arrhythmias)
- Major bleeding (defined as intracranial bleeding or bleeding requiring transfusion, endoscopy, surgery, or vasoactive drugs)
- Thrombotic event, defined as either (i) acute pulmonary embolism; (ii) deep vein thrombosis; (iii) ischaemic stroke; (iv) myocardial infarction; or (v) systemic arterial embolism.
- Use of any medications included in the RECOVERY trial protocol (including drugs in the same class) or other purported COVID-19 treatments (e.g. remdesivir, favipiravir)
- Participation in other randomised trials of interventions (vaccines or treatments) for COVID-19.

- Additional information including results of routine tests (including full blood count, coagulation and inflammatory markers, cardiac biomarkers, electro- and echo-cardiograms) and other treatments given will be collected for children in the UK. This information will be obtained and entered into the web-based IT system by a member of the hospital clinical or research staff. At some locations, electrocardiograms done as part of routine care of adult participants will also be collected.

Follow-up information is to be collected on all study participants, irrespective of whether or not they complete the scheduled course of allocated study treatment. Study staff will seek follow-up information through various means including medical staff, reviewing information from medical notes, routine healthcare systems, and registries.

For all randomised participants, vital status (alive / dead, with date and presumed cause of death, if appropriate) is to be ascertained at 28 days after first randomisation. This may be achieved through linkage to routine death registration data (e.g. in the UK) or through direct contact with the participant, their relatives, or medical staff and completion of an additional follow-up form.

##### 2.7.1 Additional assessment of safety of antibody-based therapy [UK only]

For at least the first 200 participants in each comparison in Main Randomisation part B (no additional treatment vs. convalescent plasma and no additional treatment vs. synthetic neutralising antibody), the following information will be collected on the following events occurring within the first 72 hours after randomisation:

- Sudden worsening in respiratory status
- Severe allergic reaction or other infusion reaction
- Temperature >39ºC or ≥2ºC rise above baseline
- Sudden hypotension, defined as either (i) sudden drop in systolic blood pressure of ≥30 mmHg with systolic blood pressure ≤80 mmHg; or (ii) requiring urgent medical attention
- Clinical haemolysis, defined as fall in haemoglobin plus one or more of the following: rise in lactate dehydrogenase (LDH), rise in bilirubin, positive direct antiglobulin test (DAT), or positive crossmatch
- Thrombotic event, defined as either (i) acute pulmonary embolism; or (ii) deep-vein thrombosis; or (iii) ischaemic stroke; or (iv) myocardial infarction; or (v) systemic arterial embolism.

The Data Monitoring Committee will review unblinded information on these outcomes and advise if, in their view, the collection of such information should be extended to more participants.

In addition, Serious Hazards Of Transfusion (SHOT) reporting will be conducted for all patients receiving convalescent plasma for the full duration of the study (see section 4.1).

#### 2.8 Duration of follow-up

All randomised participants are to be followed up until death, discharge from hospital or 28 days after randomisation (whichever is sooner). It is recognised that in the setting of this trial, there may be some variability in exactly how many days after randomisation, information on disease status is collected. This is acceptable and will be taken account of in the analyses and interpretation of results, the principle being that some information about post-randomisation disease status is better than none.

In the UK, longer term (up to 10 years) follow-up will be sought through linkage to electronic healthcare records and medical databases including those held by NHS Digital, Public Health England and equivalent bodies, and to relevant research databases (e.g. UK Biobank, Genomics England).

#### 2.9 Withdrawal of consent

A decision by a participant (or their parent/guardian) that they no longer wish to continue receiving study treatment should **not** be considered to be a withdrawal of consent for follow-up. However, participants (or their parent/guardian) are free to withdraw consent for some or all aspects of the study at any time if they wish to do so. In accordance with regulatory guidance, de-identified data that have already been collected and incorporated in the study database will continue to be used (and any identifiable data will be destroyed).

For participants who lack capacity, if their legal representative withdraws consent for treatment or methods of follow-up then these activities would cease.

### 3 STATISTICAL ANALYSIS

All analyses for reports, presentations and publications will be prepared by the coordinating centre at the Nuffield Department of Population Health, University of Oxford. A more detailed statistical analysis plan will be developed by the investigators and published on the study website whilst still blind to any analyses of aggregated data on study outcomes by treatment allocation.

#### 3.1 Outcomes

For each pairwise comparison with the ‘no additional treatment’ arm, the **primary objective** is to provide reliable estimates of the effect of study treatments on all-cause mortality at 28 days after randomisation (with subsidiary analyses of cause of death and of death at various timepoints following discharge).

The **secondary objectives** are to assess the effects of study treatments on duration of hospital stay; and, among patients not on invasive mechanical ventilation at baseline, the composite endpoint of death or need for invasive mechanical ventilation or ECMO.

Other objectives include the assessment of the effects of study treatments on the need for any ventilation (and duration), renal replacement therapy and new major cardiac arrhythmias.

Study outcomes will be assessed based on data recorded up to 28 days and up to 6 months after randomisation.

Where available, data from routine healthcare records (including linkage to medical databases held by organisations such as NHS Digital in the UK) and from relevant research studies (such as UK Biobank, Genomics England, ISARIC-4C and PHOSP-COVID) will allow subsidiary analyses of the effect of the study treatments on particular non-fatal events (e.g. ascertained through linkage to Hospital Episode Statistics), the influence of pre-existing major co-morbidity (e.g. diabetes, heart disease, lung disease, hepatic insufficiency, severe depression, severe kidney impairment, immunosuppression), and longer-term outcomes as well as in particular sub-categories of patient (e.g. by genotype, pregnancy).

#### 3.2 Methods of analysis

For all outcomes, comparisons will be made between all participants randomised to the different treatment arms, irrespective of whether they received their allocated treatment (“intention-to-treat” analyses).

For time-to-event analyses, each treatment group will be compared with the no additional treatment group using the log-rank test. Kaplan-Meier estimates for the time to event will also be plotted (with associated log-rank p-values). The log-rank 'observed minus expected' statistic (and its variance) will also be used to estimate the average event rate ratio (and its confidence interval) for those allocated to each treatment group versus the no additional treatment group. For binary outcomes where the timing is unknown, the risk ratio and absolute risk difference will be calculated with confidence intervals and p-value reported. For the primary outcome (death within 28 days of randomisation), discharge alive before 28 days will assume safety from the event (unless there is additional data confirming otherwise).

Pairwise comparisons within each randomisation will be made between each treatment arm and the no additional treatment arm (reference group) in that particular randomisation (main randomisation part A, B or C, and second randomisation). However, since not all treatments may be available or suitable for all patients, those in the no additional treatment arm will only be included in a given comparison if, at the point of their randomisation, they *could* alternatively have been randomised to the active treatment of interest. Allowance for multiple treatment comparisons due to the multi-arm design will be made. All p-values will be 2-sided.

Pre-specified subgroup analysis (e.g., level of respiratory support, time since onset of symptoms; sex; age group; ethnicity; use of corticosteroids) will be conducted for the primary outcome using the statistical test for interaction (or test for trend where appropriate). Sensitivity analyses will be conducted among those patients with laboratory confirmed SARS-CoV-2.

Further details will be fully described in the Statistical Analysis Plan.

### 4 DATA AND SAFETY MONITORING

#### 4.1 Recording Suspected Serious Adverse Reactions

The focus is on those events that, based on a single case, are highly likely to be related to the study medication. Examples include anaphylaxis, Stevens Johnson Syndrome, or bone marrow failure, where there is no other plausible explanation.

Any Serious Adverse Event[d] that is believed with a reasonable probability to be due to one of the study treatments will be considered a Suspected Serious Adverse Reaction (SSAR). In making this assessment, there should be consideration of the probability of an alternative cause (for example, COVID-19 itself or some other condition preceding randomisation), the timing of the event with respect to study treatment, the response to withdrawal of the study treatment, and (where appropriate) the response to subsequent re-challenge.

All SSARs should be reported by telephone to the Central Coordinating Office and recorded on the study IT system immediately.

---

[d] Serious Adverse Events are defined as those adverse events that result in death; are life-threatening; require in-patient hospitalisation or prolongation of existing hospitalisation; result in persistent or significant disability or incapacity; result in congenital anomaly or birth defect; or are important medical events in the opinion of the responsible investigator (that is, not life-threatening or resulting in hospitalisation, but may jeopardise the participant or require intervention to prevent one or other of the outcomes listed above).

[UK only] Suspected serious transfusion reactions in patients who receive convalescent plasma should additionally be reported to Serious Hazards of Transfusions (SHOT) and through the MHRA Serious Adverse Blood Reactions and Events (SABRE) system.[e]

##### 4.2 Central assessment and onward reporting of SUSARs

Clinicians at the Central Coordinating Office are responsible for expedited review of reports of SSARs received. Additional information (including the reason for considering it both serious and related, and relevant medical and medication history) will be sought.

The focus of Suspected Unexpected Serious Adverse Reaction (SUSAR) reporting will be on those events that, based on a single case, are highly likely to be related to the study medication. To this end, anticipated events that are either efficacy endpoints, consequences of the underlying disease, or common in the study population will be exempted from expedited reporting. Thus the following events will be exempted from expedited reporting:

(i) Events which are the consequence of COVID-19; and
(ii) Common events which are the consequence of conditions preceding randomisation.

Any SSARs that are not exempt will be reviewed by a Central Coordinating Office clinician and an assessment made of whether the event is “expected” or not (assessed against the relevant Summary of Product Characteristics or Investigator Brochure). Any SSARs that are not expected would be considered a Suspected Unexpected Serious Adverse Reaction (SUSAR).

All confirmed SUSARs will be reported to the Chair of the DMC and to relevant regulatory authorities, ethics committees, and investigators in an expedited manner in accordance with regulatory requirements.

##### 4.3 Recording other Adverse Events

In addition to recording Suspected Serious Adverse Reactions (see section 4.1), information will be collected on all deaths and efforts will be made to ascertain the underlying cause. Other serious or non-serious adverse events will not be recorded unless specified in section 2.7. It is anticipated that for some substudies, more detailed information on adverse events (e.g. through linkage to medical databases) or on other effects of the treatment (e.g. laboratory or radiological features) will be recorded and analysed but this is not a requirement of the core protocol.

##### 4.4 Role of the Data Monitoring Committee (DMC)

During the study, interim analyses of all study data will be supplied in strict confidence to the independent DMC. The DMC will request such analyses at a frequency relevant to the emerging data from this and other studies.

---

[e] https://www.shotuk.org/reporting/

The DMC will independently evaluate these analyses and any other information considered relevant. The DMC will determine if, in their view, the randomised comparisons in the study have provided evidence on mortality that is strong enough (with a range of uncertainty around the results that is narrow enough) to affect national and global treatment strategies. In such a circumstance, the DMC will inform the Trial Steering Committee who will make the results available to the public and amend the trial arms accordingly. Unless this happens, the Trial Steering Committee, Chief Investigator, study staff, investigators, study participants, funders and other partners will remain blind to the interim results until 28 days after the last patient has been randomised for a particular intervention arm (at which point analyses may be conducted comparing that arm with the no additional treatment arm).

The DMC will review the safety and efficacy analyses among children (age <18 years) both separately and combined with the adult data. As described in section 2.7.1, the DMC will advise if collection of information relating to the safety of convalescent plasma should be extended beyond the first 200 patients enrolled to each comparison in Main Randomisation part B.

##### 4.5 Blinding

This is an open-label study. However, while the study is in progress, access to tabular results of study outcomes by allocated treatment allocation will not be available to the research team, patients, or members of the Trial Steering Committee (unless the DMC advises otherwise).

#### 5 QUALITY MANAGEMENT

##### 5.1 Quality By Design Principles

In accordance with the principles of Good Clinical Practice and the recommendations and guidelines issued by regulatory agencies, the design, conduct and analysis of this trial is focussed on issues that might have a material impact on the wellbeing and safety of study participants (hospitalised patients with suspected or confirmed SARS-CoV-2 infection) and the reliability of the results that would inform the care for future patients.

The critical factors that influence the ability to deliver these quality objectives are:

- to minimise the burden on busy clinicians working in an overstretched hospital during a major epidemic
- to ensure that suitable patients have access to the trial medication without impacting or delaying other aspects of their emergency care
- to provide information on the study to patients and clinicians in a timely and readily digestible fashion but without impacting adversely on other aspects of the trial or the patient's care
- to allow individual clinicians to use their judgement about whether any of the treatment arms are not suitable for the patient
- to collect comprehensive information on the mortality and disease status

In assessing any risks to patient safety and well-being, a key principle is that of proportionality. Risks associated with participation in the trial must be considered in the context of usual care. At present, there are no proven treatments for COVID-19, basic hospital care (staffing, beds, ventilatory support) may well be overstretched, and mortality for hospitalised patients may be around 10% (or more in those who are older or have significant co-morbidity).

#### 5.2 Training and monitoring

The focus will be on those factors that are critical to quality (i.e. the safety of the participants and the reliability of the trial results). Remedial actions would focus on issues with the potential to have a substantial impact on the safety of the study participants or the reliability of the results.

The study will be conducted in accordance with the principles of International Conference on Harmonisation Guidelines for Good Clinical Research Practice (ICH-GCP) and relevant local, national and international regulations. Any serious breach of GCP in the conduct of the clinical trial will be handled in accordance with regulatory requirements. Prior to initiation of the study at each Local Clinical Centre (LCC), the Central Coordinating Office (CCO) or relevant Regional Coordinating Centre (RCC) will confirm that the LCC has adequate facilities and resources to carry out the study. LCC lead investigators and study staff will be provided with training materials.

In the context of this epidemic, visits to hospital sites is generally not appropriate as they could increase the risks of spreading infection, and in the context of this trial they generally would not influence the reliability of the trial results or the well-being of the participants. In exceptional circumstances, the CCO or RCC may arrange monitoring visits to LCCs as considered appropriate based on perceived training needs and the results of central statistical monitoring of study data.[9,10] The purpose of such visits will be to ensure that the study is being conducted in accordance with the protocol, to help LCC staff to resolve any local problems, and to provide extra training focussed on specific needs. No routine source data verification will take place.

In the UK, training of laboratory and transfusion staff and initiation of convalescent plasma delivery will be performed by NHS Blood and Transplant Clinical Trials Unit.

#### 5.3 Data management

LCC clinic staff will use the bespoke study web-based applications for study management and to record participant data (including case report forms) in accordance with the protocol. Data will be held in central databases located at the CCO or on secure cloud servers. In some circumstances (e.g. where there is difficulty accessing the internet or necessary IT equipment), paper case report forms may be required with subsequent data entry by either LCC or CCO staff. Although data entry should be mindful of the desire to maintain integrity and audit trails, in the circumstances of this epidemic, the priority is on the timely entry of data that is sufficient to support reliable analysis and interpretation about treatment effects. CCO staff will be responsible for provision of the relevant web-based applications and for generation of data extracts for analyses.

All data access will be controlled by unique usernames and passwords, and any changes to data will require the user to enter their username and password as an electronic signature in accordance with regulatory requirements.[11] Staff will have access restricted to the functionality and data that are appropriate for their role in the study.

##### 5.4 Source documents and archiving

Source documents for the study constitute the records held in the study main database. These will be retained for at least 25 years from the completion of the study. Identifiable data will be retained only for so long as it is required to maintain linkage with routine data sources (see section 2.8), with the exception of children for whom such data must be stored until they reach 21 years old (due to the statute of limitations). The sponsor and regulatory agencies will have the right to conduct confidential audits of such records in the CCO and LCCs (but should mindful of the workload facing participating hospitals and the infection control requirements during this epidemic).

#### 6 OPERATIONAL AND ADMINISTRATIVE DETAILS

##### 6.1 Sponsor and coordination

The University of Oxford will act as the trial Sponsor. The trial will be coordinated by a Central Coordinating Office (CCO) within the Nuffield Department of Population Health staffed by members of the two registered clinical trials units – the Clinical Trial Service Unit and the National Perinatal Epidemiology Unit Clinical Trials Unit. The CCO will oversee Regional Coordinating Centres which will assist with selection of Local Clinical Centres (LCCs) within their region and for the administrative support and monitoring of those LCCs. The data will be collected, analysed and published independently of the source of funding.

##### 6.2 Funding

This study is supported by a grant to the University of Oxford from UK Research and Innovation/National Institute for Health Research (NIHR) and by core funding provided by NIHR Oxford Biomedical Research Centre, the Wellcome Trust, the Bill and Melinda Gates Foundation, Department for International Development, Health Data Research UK, NIHR Health Protection Unit in Emerging and Zoonotic Infections and the Medical Research Council Population Health Research Unit, and NIHR Clinical Trials Unit Support Funding.

##### 6.3 Indemnity

The University has a specialist insurance policy in place which would operate in the event of any participant suffering harm as a result of their involvement in the research (Newline Underwriting Management Ltd, at Lloyd's of London). In the UK, NHS indemnity operates in respect of the clinical treatment that is provided.

##### 6.4 Local Clinical Centres

The study will be conducted at multiple hospitals (LCCs) within each region. At each LCC, a lead investigator will be responsible for trial activities but much of the work will be carried out by medical staff attending patients with COVID-19 within the hospital and by hospital research nurses, medical students and other staff with appropriate education, training, and experience. Where LCCs plan to recruit children the principal investigator will co-opt support from a local paediatrician and/or neonatologists to oversee the management of children and infants in the trial.

#### 6.5 Supply of study treatments

For licensed treatments (e.g. lopinavir-ritonavir, corticosteroids, tocilizumab) all aspects of treatment supply, storage, and management will be in accordance with standard local policy and practice for prescription medications. Treatment issue to randomised participants will be by prescription. Such study treatments will not be labelled other than as required for routine clinical use. They will be stored alongside other routine medications with no additional monitoring. No accountability records will be kept beyond those used for routine prescriptions.

For unlicensed treatments, manufacture, packaging, labelling and delivery will be the responsibility of the pharmaceutical company and, in the UK, the Department of Health and Social Care. Each LCC will maintain an accountability log and will be responsible for the storage and issue of study treatment. If treatments require storage at a specific temperature, LCCs can use existing temperature-controlled facilities and associated monitoring. Treatment issue to randomised participants will be in accordance with local practice (and may be in line with the processes required for routine prescriptions or compassionate use).

For convalescent plasma in the UK, manufacture, packaging, and delivery will be the responsibility of the relevant UK Blood Service (NHS Blood and Transplant for England, Welsh Blood Service for Wales, Scottish National Blood Transfusion Service for Scotland, and the Northern Ireland Blood Transfusion Service for Northern Ireland). Convalescent plasma will be labelled in accordance with regulatory requirements and the unit will be issued to the ward for a named patient in a bag marked for clinical trial use only.

Treatment will be issued to randomised participants by prescription.

#### 6.6 End of trial

The end of the scheduled treatment phase is defined as the date of the last follow-up visit of the last participant. In the UK, it is intended to extend follow-up for a year or more beyond the final study visit through linkage to routine medical records and central medical databases. The end of the study is the date of the final data extraction from NHS Digital (anticipated to be 10 years after the last patient is enrolled).

#### 6.7 Publications and reports

The Trial Steering Committee will be responsible for drafting the main reports from the study and for review of any other reports. In general, papers initiated by the Trial Steering Committee (including the primary manuscript) will be written in the name of the RECOVERY Collaborative Group, with individual investigators named personally at the end of the report (or, to comply with journal requirements, in web-based material posted with the report).

The Trial Steering Committee will also establish a process by which proposals for additional publications (including from independent external researchers) are considered by the Trial Steering Committee. The Trial Steering Committee will facilitate the use of the study data and approval will not be unreasonably withheld. However, the Trial Steering Committee will need to be satisfied that any proposed publication is of high quality, honours the commitments made to the study participants in the consent documentation and ethical approvals, and is compliant with relevant legal and regulatory requirements (e.g. relating to data protection and privacy). The Trial Steering Committee will have the right to review and comment on any draft manuscripts prior to publication.

#### 6.8 Substudies

Proposals for substudies must be approved by the Trial Steering Committee and by the relevant ethics committee and competent authorities (where required) as a substantial amendment or separate study before they begin. In considering such proposals, the Trial Steering Committee will need to be satisfied that the proposed substudy is worthwhile and will not compromise the main study in any way (e.g. by impairing recruitment or the ability of the participating hospitals to provide care to all patients under their care).

### 7 VERSION HISTORY

| Version number | Date | Brief Description of Changes |
|---|---|---|
| 1.0 | 13-Mar-2020 | Initial version |
| 2.0 | 21-Mar-2020 | Addition of hydroxychloroquine. Administrative changes and other clarifications. |
| 3.0 | 07-Apr-2020 | Extension of eligibility to those with suspected COVID-19<br>Addition of azithromycin arm.<br>Addition of inclusion of adults who lack permanently lack capacity.<br>Change to primary outcome from in-hospital death to death within 28 days of randomisation. |
| 4.0 | 14-Apr-2020 | Addition of second randomisation to tocilizumab vs. standard of care among patients with progressive COVID-19. |
| 5.0 | 24-Apr-2020 | Addition of children to study population. |
| 6.0 | 14-May-2020 | Addition of convalescent plasma |
| 7.0 | 18-Jun-2020 | Allowance of randomisation in part B of main randomisation without part A.<br>Removal of hydroxychloroquine and dexamethasone treatment arms. |
| 8.0 | 03-Jul-2020 | Removal of lopinavir-ritonavir<br>Addition of intravenous immunoglobulin arm for children<br>Changes to corticosteroid dosing for children.<br>Addition of baseline serum sample in convalescent plasma randomisation |
| 9.0 | 10-Sep-2020 | Addition of synthetic neutralizing antibodies<br>Additional baseline data collection<br>Addition of countries outside UK |
| 9.1 | 18-Sep-2020 | Addition of information about vaccination of children of pregnant mothers receiving REGN10933+REGN10987 |
| 9.2 [not submitted in UK] | 15-Oct-2020 | Additional information for countries outside UK |
| 10.0 | 26-Oct-2020 | Addition of main randomisation part C<br>General updates to avoid duplication and improve clarity |
| 10.1 | 01-Nov-2020 | Additional information for pregnant women |

### 8 APPENDICES

#### 8.1 Appendix 1: Information about the treatment arms

All patients will receive usual care in the participating hospital.

**[UK only] Corticosteroids:** Favourable modulation of the immune response is considered one of the possible mechanisms by which corticosteroids might be beneficial in the treatment of severe acute respiratory coronavirus infections, including COVID-19, SARS and MERS. Common to severe cases of these infections is the presence of hypercytokinemia (a cytokine 'storm') and development of acute lung injury or acute respiratory distress syndrome (ARDS).[12-15] Pathologically, diffuse alveolar damage is found in patients who die from these infections.[16] A growing volume of clinical trial data from patients with severe community acquired pneumonia, ARDS and septic shock suggest benefit from low-to-moderate dose corticosteroids in relation to mortality and length of stay.[17-19]

In trials of low-to-moderate doses of corticosteroids, the main adverse effect has been hyperglycaemia.[18,20] A systematic review of (mainly low-dose) corticosteroid trials in severe sepsis and septic shock did not identify any increased risk of gastroduodenal bleeding, superinfection or neuromuscular weakness; an association with an increased risk of hyperglycaemia (RR 1.16, 95% CI 1.07 to 1.25) and hypernatraemia (RR 1.61, 95% CI 1.26 to 2.06) was noted.[21]

Methylprednisolone is a corticosteroid with mainly glucocorticoid activity. It is used in the treatment of conditions in which rapid and intense corticosteroid effect is required. Its licensed indications for paediatrics include a wide range of conditions including inflammatory disorders, allergic disorders, draft rejection reactions, severe erythema multiforme, juvenile idiopathic arthritis, and many others. In the paediatric population, a dosage of 10 mg/kg/day to a maximum of 1 g/day for up to 3 days is recommended in the treatment of graft rejection reactions following transplantation. A higher dosage of 30 mg/kg/day to a maximum of 1 g/day for up to 3 days is recommended for the treatment of haematological, rheumatic, renal and dermatological conditions (Source: British National Formulary for Children). Storage should be as per conditions in the Summary of Product Characteristics.

PIMS-TS is associated with a hyper-inflammatory state with elevated ESR, C-reactive protein, D-dimers, lactate dehydrogenase, ferritin, and increased levels of pro-inflammatory cytokines including as IL-1 and IL-6. While there is a pharmacological basis for using high dose methylprednisolone, the Delphi consensus process conducted by NHS England identified equipoise for its use in the treatment of PIMS-TS.

**Azithromycin:** Azithromycin is a macrolide antibiotic. In addition to their antimicrobial properties, the macrolide antibiotics are known to have immunomodulatory activity. The mechanism of immunomodulation includes decreased production of pro-inflammatory cytokines and inhibition of neutrophil activation.[22-24] Macrolides are widely used both in infectious pneumonia due to their antimicrobial activity and in chronic inflammatory lung disease due to the immunomodulatory effects.[25] Azithromycin is preferred over other macrolides because data suggest it has stronger immunomodulatory effects than other macrolides.[24]

The use of macrolides in influenza-associated pneumonia has been associated with a faster reduction in inflammatory cytokines and, in combination with naproxen, decreased mortality.[26-28] Observational studies in MERS-CoV have not demonstrated a mortality benefit of macrolide use.[29] Macrolides have not been evaluated in severe betacoronavirus infections in randomised controlled trials. The safety of macrolides is well established.

**[UK only] Intravenous immunoglobulin (IVIg):** IVIg is human normal immunoglobulin, available in a number of different preparations in routine NHS practice. The NHS England consensus process has established intravenous immunoglobulin as the interim first line treatment in non-shocked COVID-associated PIMS-TS and also that there is need for evaluation of intravenous immunoglobulin and corticosteroid in the initial management of PIMS-TS. In the similar but different disease process known as Kawasaki Diseases, randomised controlled trials and meta-analyses have demonstrated that early recognition and treatment of KD with IVIg (and aspirin) reduces the occurrence of coronary artery aneurysms. Current published guidelines recommend a dose of 2 g/kg IVIg given as a single infusion, as this has been shown to reduce the coronary artery aneurysm rate compared to a lower divided dose regimen.[30]

IVIg is licensed for immunomodulation in adults, children and adolescents (0-18 years) in a number of clinical conditions including but not limited to primary immune thrombocytopenia, Guillain Barré syndrome, Kawasaki disease (in association with aspirin), chronic inflammatory demyelinating polyradiculoneuropathy and multifocal motor neuropathy.

**Tocilizumab** is a monoclonal antibody that binds to the receptor for IL-6, blocking IL-6 signalling and reduces inflammation. Tocilizumab is licensed for use in patients with rheumatoid arthritis and for use in people aged at least 2 years with chimeric antigen receptor (CAR) T cell-induced severe or life-threatening cytokine release syndrome.

Severe COVID-19 is associated with a hyper-inflammatory state with elevated ESR, C-reactive protein, D-dimers, lactate dehydrogenase, ferritin, and increased levels of pro-inflammatory cytokines including as IL-1 and IL-6.[4,31,32] There have been published and unpublished (pre-print) case series reports of the successful treatment of COVID-19 patients with IL-6 inhibitors.[31,33] IL-6 inhibitors have not been evaluated for the treatment of COVID-19 in randomised controlled trials.

**[UK only] Convalescent plasma:** Convalescent plasma treatment, containing high titres of polyclonal antibody, has been used to treat severe viral pneumonias. Many studies have been small or poorly controlled but have reported beneficial effects in avian influenza[34-36], influenza A (H1N1) infections in 1915-1917[37] and 2009/2010[38,39], and seasonal influenza B[40]. More relevant to SARS-CoV-2, a systematic review of convalescent plasma treatment in SARS-CoV infections in 2003 identified eight observational studies that all reported improved mortality associated with the use of convalescent plasma – infected patients received various amounts of convalescent plasma.[41] Recent studies in seasonal influenza A and in MERS-CoV highlight the importance of high avidity and high titre antibodies respectively.[42,43]

Convalescent plasma therapy had been given to at least 245 COVID-19 patients by the end of February 2020, and, according to a Chinese health official, 91 cases had shown improvement in clinical indicators and symptoms. Five small case series (26 patients in total) have been published that report the use of convalescent plasma in people with COVID-19 infection.[44-48] These studies have reported clinical and radiological improvements after treatment with convalescent plasma. However, these small uncontrolled studies have significant flaws and the reported effects are unreliable. Convalescent plasma is currently being tested in the REMAP-CAP trial among patients on intensive care units.

**[UK only] Synthetic neutralising antibodies (REGN-COV2):** Synthetic monoclonal antibodies (mAbs) have been demonstrated to be safe and effective in viral disease when used as prophylaxis (respiratory syncytial virus and rabies) and treatment (Ebola virus disease).[49,50] Anti-SARS-CoV-2 mAbs are designed to bind to and neutralise the virus. In addition, mAbs may have additional effector functions (antibody dependent phagocytosis and cytotoxicity) through binding to SARS-CoV-2 spike protein expressed on the surface of cells. Anti-SARS-CoV-2 spike protein neutralizing mAbs have demonstrated in vivo efficacy in both therapeutic and prophylactic settings in mouse, and non-human primates models, with decreases in viral load and lung pathology.[51-53]

Regeneron has developed 2 non-competing, high-affinity human IgG1 anti-SARS-CoV-2 mAbs, REGN10933 and REGN10987 that bind specifically to the receptor binding domain of the spike glycoprotein of SARS-CoV-2, blocking viral entry into host cells.[54,55] REGN10933 and REGN10987 are both potent neutralizing antibodies that block the interaction between the spike protein and its canonical receptor angiotensin-converting enzyme 2. REGN10933 and REGN10987 are intended to be utilized as a combination treatment, known as REGN-COV2, and should not be used individually as monotherapy. A combination of antibodies that bind to non-overlapping epitopes may minimize the likelihood of loss of antiviral activity due to naturally circulating viral variants or development of escape mutants under drug pressure. In animal studies (rhesus macaques and hamsters) the antibody cocktail (REGN10933+REGN10987) reduced virus load in lower and upper airway and decreased virus induced pathological sequelae when administered prophylactically or therapeutically.[56]

**Aspirin:** Patients with COVID-19 appear to be at high risk of thromboembolism.[57] Classical risk factors for thromboembolism are common in the COVID-19 hospitalised population, but the relatively low incidence of deep vein thrombosis compared to the incidence of pulmonary embolism (and the often peripheral location of the pulmonary emboli observed) suggests that inflammation and associated endothelial injury and platelet activation may be an important cause of thromboembolism in this patient population.[57,58] Therefore antiplatelet therapy is a potential thromboprophylactic therapy in COVID-19. It is also being tested in the REMAP-CAP trial.

#### 8.2 Appendix 2: Drug specific contraindications and cautions

**Corticosteroid (children only)**
- Known contra-indication to short-term corticosteroid.

**Azithromycin**
- Known prolonged QTc interval*
- Co-administration with chloroquine or hydroxychloroquine
- Known hypersensitivity to macrolide antibiotic

**Intravenous Immunoglobulin (children only)**
- Hypersensitivity to the active substance (human immunoglobulins) or to any of the excipients
- Patients with selective IgA deficiency who developed antibodies to IgA, as administering an IgA-containing product can result in anaphylaxis
- Hyperprolinaemia type I or II.

Potential complications can often be avoided by ensuring that participants:
i. are carefully monitored for any symptoms throughout the infusion period;
ii. have urine output and serum creatinine levels monitored; and
iii. avoid concomitant use of loop diuretics.

Such monitoring should occur regularly during the admission, at a frequency appropriate to the illness of the child.

**Aspirin**
- Age <18 years old
- Known hypersensitivity to aspirin
- Recent major bleeding that precludes use of aspirin in opinion of managing physician
- Current use of aspirin, clopidogrel or other antiplatelet therapy

**Tocilizumab**
- Known hypersensitivity to tocilizumab.
- Evidence of active TB infection[f]
- Clear evidence of active bacterial, fungal, viral, or other infection (besides COVID-19)

(Note: Pregnancy and breastfeeding are not exclusion criteria.)

**Convalescent plasma**
- Known moderate or severe allergy to blood components*
- Not willing to receive a blood product*

**Synthetic neutralising antibodies (REGN-COV2)**

[f] Note: The risk of reactivation of latent tuberculosis with tocilizumab is considered to be extremely small.

- Intravenous immunoglobulin treatment during current admission*
- Age <12 years old or child with weight <40kg*

(Note: Pregnancy and breastfeeding are not exclusion criteria.)

The infusion of synthetic neutralising antibodies should be interrupted if any of the following are observed (or worsen during the infusion): sustained/severe cough, rigors/chills, rash, pruritus, urticaria, diaphoresis, hypotension, dyspnoea, vomiting, or flushing. The reactions should be treated symptomatically, and the infusion may be restarted at 50% of the original rate once all symptoms have ceased (or returned to baseline) and at the discretion of the managing physician. If the managing physician feels there is medical need for treatment or discontinuation of the infusion other than described above, they should use clinical judgement to provide appropriate response according to typical clinical practice.

Pregnant women that are administered REGN10933 and REGN10987 must be advised that live vaccines should be avoided in children with *in utero* exposure to biologics for at least the first 6 months of life.

* If these conditions are recorded on the baseline case report form, patients will be ineligible for randomisation to that arm of the study.

Note: This study is being conducted within hospitals. Therefore use of medication will be subject to standard medication reviews (typically within 48 hours of enrolment) and clinical assessments (including appropriate blood tests) which will guide modifications to both the study treatment and use of concomitant medication (e.g. in the case of potential drug interactions). The doctor may decide whether it is appropriate to stop such medications temporarily to allow the patient to complete the course of their assigned intervention.

Although all available data on use in pregnancy are reassuring, since the effect of some of the treatments on unborn babies is uncertain, female participants who are not already pregnant will be advised that they should not get pregnant within 3 months of the completion of trial treatment(s).

#### 8.3 Appendix 3: Paediatric dosing information

Children (aged <18 years old) will be recruited in the UK only.

##### Main Randomisation Part A

| Arm | Route | Weight # | Dose (Duration for all arms = 10 days or until discharge from hospital) |
|---|---|---|---|
| **No additional treatment** | - | - | - |
| **Corticosteroid**<br>- Solution for injection*<br>- Powder for solution for injection*<br>-<br>*various strengths available | Intravenous | All Including pre-term neonates | Neonates/infants with a corrected gestational age of ≤44 weeks<br><br>**Hydrocortisone** (IV):<br>0.5 mg/kg every 12 hours for 7 days and then 0.5mg/kg once daily for 3 days |
| | Intravenous | All | For all other children (with PIMS-TS):<br><br>**Methylprednisolone sodium succinate**<br>10 mg/kg (as base) once daily for 3 days (max 1 gram)<br><br>No additional oral corticosteroid should be prescribed to follow the 3 day treatment course. |
| **Human normal immunoglobulin (IVIg)**<br><br>- solution for infusion<br><br>*various strengths available | Intravenous | All | For children with corrected gestational age >44 weeks and <18 years with PIMS-TS phenotype:<br><br>2 g/kg as a single dose. (Dose should be based on ideal body weight in line with NHS England guidance.) |
| **Azithromycin**<br><br>- 40mg in 1mL oral suspension<br>- 250mg tablet/capsule<br>- 500mg tablet/capsule<br>- 500mg powder for solution for infusion | Oral<br>or<br>Nasogastric<br>or<br>Intravenous | ≤ 16 kg Including preterm neonates | 10 mg/kg once daily§ |
| | | 17 - 25 kg | 200 mg once daily§ |
| | | 26 - 35 kg | 300 mg once daily§ |
| | | 36 - 45 kg | 400 mg once daily§ |
| | | ≥ 46 kg | 500 mg once daily§ |

### Weight to be rounded to the nearest kg unless dosage expressed as mg/kg or mL/kg.
§ for 10 days (or until discharge from hospital)

#### Main Randomisation Part B

| | | | |
|---|---|---|---|
| **Convalescent Plasma** | Intravenous | | 5 mL/kg of ABO compatible convalescent plasma intravenous up to standard adult dose of 275 mLs per day on study days 1 and 2.<br><br>Minimum of 12 hour interval between 1st and 2nd units.<br><br>Convalescent plasma for neonates and infants up to one year of age needs to be ordered on a named patient basis from the relevant National Blood Service to ensure the unit meets neonatal requirements. Data transfer storage and retention will be in line with NHSBT standard procedures and protocols. |
| **Synthetic neutralising antibodies** (REGN10933 + REGN10987) | Intravenous | ≥12 years And ≥40 kg | 8 g (4 g of each monoclonal antibody) |

#### Second stage randomisation (Patients < 1 year of age will NOT be eligible)

| Arm | Route | Weight | Dose |
|---|---|---|---|
| **No additional treatment** | - | - | - |
| **Tocilizumab** | Intravenous | Infants < 1 year excluded | |
| | | < 30 kg | 12 mg/kg<br>A second dose may be given ≥12 and ≤24 hours later if, in the opinion of the attending clinicians, the patient’s condition has not improved. |
| | | ≥ 30 kg | 8 mg/kg (max 800 mg)<br>A second dose may be given ≥12 and ≤24 hours later if, in the opinion of the attending clinicians, the patient’s condition has not improved. |

#### 8.4 Appendix 4: Use of IMPs in pregnant and breastfeeding women

All trial drugs (except REGN-COV2) have been used in pregnant women with pre-existing medical disorders where benefits outweigh the risks to fetus or woman, including in the first trimester. The existing data related to each drug is summarized below.

**Azithromycin**
Azithromycin is used in pregnancy to treat genital Chlamydia trachomatis infection, with a Cochrane systematic review and meta-analysis reporting fewer gastrointestinal side-effects compared to erythromycin, and inconsistent results on risk of preterm birth, preterm rupture of membranes, perinatal mortality and low birthweight, confounded by the indication for treatment.[59] A recent systematic review and meta-analysis of all macrolide antibiotics acknowledges potential bias in child outcome reports due to treatment indication.[60] The UK Teratology Information Service monograph concludes that there is no definitive evidence linking azithromycin with increased risk of miscarriage or congenital malformations.[61] Azithromycin is detected in only low levels in breastmilk and is not expected to cause adverse events in breastfed infants (reviewed in Lactmed database: www.ncbi.nlm.nih.gov/books/NBK501200/) _Azithromycin has also been used in several trials in preterm infants as a prophylactic treatment to prevent bronchopulmonary dysplasia.[62]

**Convalescent plasma**
Convalescent plasma is plasma from people who had confirmed COVID-19 (SARS-Cov-2) infection, and have now recovered and been free of the infection for 28 days. The plasma contains antibodies that their immune systems have produced in fighting the virus. It is hoped that giving this plasma will help speed up recovery of a patient with active infection and improve their chances of survival. Plasma is already used as a treatment in pregnant patients who are bleeding,[63] or have particular blood conditions.[64,65] The plasma being used in this trial is from a selected donor and hopefully contains anti-SARS-Cov-2 antibodies, but is otherwise no different. Plasma infusions can occasionally cause side effects. Mostly this is a rise in temperature, itching or a rash, and in very extreme cases, anaphylaxis. Other potential complications include breathlessness and changes in blood pressure. Monitoring of pulse and blood pressure takes place before and after the infusion. There is no risk of miscarriage or fetal loss, preterm birth, preterm rupture of membranes, perinatal mortality or low birthweight, from plasma transfusions and there are no concerns with breast feeding.

**REGN-COV2 Monoclonal antibodies**
Monoclonal antibodies have been used as therapeutic agents in pregnancy over recent years, for a variety of conditions. Human monoclonal antibodies in use in pregnancy include anti-TNF agents, such as adalimumab, indicated for a variety of chronic inflammatory diseases such as rheumatoid arthritis and inflammatory bowel disease. Data have recently accumulated from a variety of cohort and registry studies indicating that such exposure in pregnancy was not associated with an increased risk for adverse pregnancy outcomes, when compared to unexposed pregnancies with the same underlying medical diseases.[66] This is supported by a consensus report on immunosuppressives and biologics during pregnancy and lactation, confirming no evidence of elevated adverse pregnancy outcomes or malformation risks.[67] Some monoclonal antibodies are transported across the placenta (and may also enter breast milk) but as REGN10933 and REGN10987 do not have any human targets such exposure should not be associated with risk of harm. Pregnant women, just like other patients with COVID-19, are at significant risk from the infection itself

(particularly those in the third trimester.[68,69] All pregnant women in RECOVERY are entered into the UK Obstetric Surveillance System which follows all pregnancies to their conclusion.[69] Given the early safety experience with REGN10933+REGN10987 it would appear appropriate not to exclude pregnant women from this aspect of the trial (as such exclusion would inhibit the development of treatments for this population).[70]

**Aspirin**

Aspirin is widely used for the prevention of pre-eclampsia in pregnant women at increased risk of the disease. A recent Cochrane meta-analysis on this topic included seventy-seven trials (40,249 women) taking aspirin at doses between 60 and 150mg daily.[71] In most trials, aspirin was started from 12 weeks' gestation, although a more recent meta-analysis has reported eight trials (1426 women) in which aspirin was initiated in the first trimester.[72] In light of the clear evidence of effectiveness, 75-150mg aspirin is recommended for pre-eclampsia prophylaxis in NICE guidelines for management of hypertension in pregnancy (NG133), and in the NHS England document 'Saving Babies' Lives for women at increased risk of placental dysfunction disorders.[73,74] There is some ongoing uncertainty as to the optimal dose (75mg vs. 150mg) for pre-eclampsia prophylaxis, but both doses are in widespread clinical use in pregnancy in the UK for these indications and in other conditions (e.g. in pregnant women with antiphospholipid syndrome).

**Tocilizumab**

Two pharmaceutical global safety registry database studies have reported on tocilizumab use in pregnancy, including outcomes from 288 pregnancies [75] and 61 pregnancies,[76] typically for rheumatoid or other arthritides, and with the majority having received the drug in the first trimester. These data suggest that the rates of congenital abnormality, spontaneous pregnancy loss and other adverse outcomes were not higher than in the general population.[76] Small studies have shown that tocilizumab is transferred to the fetus with serum concentrations approximately 7-fold lower than those observed in maternal serum at the time of birth.[77] Very low concentrations of tocilizumab are identified in breast milk and no drug is transferred into the serum of breast fed infants.[77,78] Women should be advised that if treated after 20 weeks' gestation, their infant should not be immunised with live vaccines (rotavirus and BCG) for the first 6 months of life. All non-live vaccinations are safe and should be undertaken.[79]

#### 8.5 Appendix 5: Organisational Structure and Responsibilities

##### Chief Investigator

The Chief Investigator has overall responsibility for:

(i) Design and conduct of the Study in collaboration with the Trial Steering Committee;
(ii) Preparation of the Protocol and subsequent revisions;

##### Trial Steering Committee

The Trial Steering Committee (see Section 8.6 for list of members) is responsible for:

(i) Agreement of the final Protocol and the Data Analysis Plans;
(ii) Reviewing progress of the study and, if necessary, deciding on Protocol changes;
(iii) Review and approval of study publications and substudy proposals;
(iv) Reviewing new studies that may be of relevance.

##### Regional (South East Asia) Steering Committee

The regional SEA Steering Committee (see Section 8.6 for list of members) is responsible for:

(i) Reviewing progress of the study in South East Asia;
(ii) Review of study publications and substudy proposals;
(iii) Considering potential new therapies to be included in South East Asia;
(iv) Assisting RCC in selection of LCCs
(v) Reviewing new studies that may be of relevance.

##### Data Monitoring Committee

The independent Data Monitoring Committee is responsible for:

(i) Reviewing unblinded interim analyses according to the Protocol;
(ii) Advising the Steering Committee if, in their view, the randomised data provide evidence that may warrant a change in the protocol (e.g. modification or cessation of one or more of the treatment arms).

##### Central Coordinating Office (CCO)

The CCO is responsible for the overall coordination of the Study, including:

(i) Study planning and organisation of Steering Committee meetings;
(ii) Ensuring necessary regulatory and ethics committee approvals;
(iii) Development of Standard Operating Procedures and computer systems
(iv) Monitoring overall progress of the study;
(v) Provision of study materials to RCCs/LCCs;
(vi) Monitoring and reporting safety information in line with the protocol and regulatory requirements;
(vii) Dealing with technical, medical and administrative queries from LCCs.

##### Regional Coordinating Centre (RCC)

The RCCs are responsible for:

(i) Ensuring necessary regulatory and ethics committee approvals;
(ii) Provision of study materials to LCCs;
(iii) Dealing with technical, medical and administrative queries from LCCs.

##### Local Clinical Centres (LCC)

The LCC lead investigator and LCC clinic staff are responsible for:

(i) Obtaining all relevant local permissions (assisted by the CCO)
(ii) All trial activities at the LCC, including appropriate training and supervision for clinical staff
(iii) Conducting trial procedures at the LCC in line with all relevant local policies and procedures;
(iv) Dealing with enquiries from participants and others.

#### 8.6 Appendix 5: Organisational Details

##### STEERING COMMITTEE

(Major organisational and policy decisions, and scientific advice; blinded to treatment allocation)

| | |
|---|---|
| Chief Investigator | Peter Horby |
| Deputy Chief Investigator | Martin Landray |
| Clinical Trial Unit Lead | Richard Haynes |
| Co-investigators | Kenneth Baillie (Scotland Lead), Lucy Chappell, Saul Faust, Thomas Jaki, Katie Jeffery, Edmund Juszczak, Wei Shen Lim, Marion Mafham, Alan Montgomery, Kathy Rowan, Guy Thwaites, Jeremy Day (South East Asia Leads) |

##### South East Asia Steering Committee (Members TBD)

| | |
|---|---|
| Regional Lead Investigators | Guy Thwaites, Jeremy Day |
| Country Lead Investigators: | TBD (Nepal), TBD (VietNam), TBD (Indonesia) |
| MOH or local country representatives: | TBD (Nepal), TBD (VietNam), TBD (Indonesia) |
| Independent members: | TBD (3 members) |

##### DATA MONITORING COMMITTEE

(Interim analyses and response to specific concerns)

| | |
|---|---|
| Chair | Peter Sandercock |
| Members | Janet Darbyshire, David DeMets, Robert Fowler, David Lalloo, Ian Roberts, Janet Wittes |
| Statisticians (non-voting) | Jonathan Emberson, Natalie Staplin |

### 10 CONTACT DETAILS

Website: www.recoverytrial.net
(copies of this protocol and related forms and information can be downloaded)

**RECOVERY Central Coordinating Office:**
Richard Doll Building, Old Road Campus, Roosevelt Drive, Oxford OX3 7LF
United Kingdom

**RECOVERY Vietnam:**
Oxford University Clinical Research Unit, Centre for Tropical Medicine, 764 Vo Van Kiet, District 5, Ho Chi Minh City, Vietnam


**RECOVERY Indonesia:**
Eijkman Oxford Clinical Research Unit (EOCRU), Eijkman Institute for Molecular Biology
Jl. P. Diponegoro No. 69, Jakarta-Indonesia 10430

**RECOVERY Nepal:**
Clinical Trial Unit, Oxford University Clinical Research Unit-Nepal, Patan Academy of Health Sciences, Kathmandu, Nepal

**To RANDOMISE a patient, visit:**

RECOVERY
Randomised Evaluation of COVID-19 Therapy

Website: www.recoverytrial.net

#### Substantial amendments to RECOVERY trial protocol

| Protocol version | Date | Randomisation | Treatment arms |
|---|---|---|---|
| 1.0 | 13-Mar-2020 | Main (part A) | No additional treatment<br>Lopinavir-ritonavir[a]<br>Low-dose corticosteroid[b]<br>Nebulised Interferon-ß-1a (never activated) |
| 2.0 | 23-Mar-2020 | Main (part A) | No additional treatment<br>Lopinavir-ritonavir[a]<br>Low-dose corticosteroid[b]<br>Hydroxychloroquine |
| 3.0 | 07-Apr-2020 | Main (part A) | No additional treatment<br>Lopinavir-ritonavir[a]<br>Low-dose corticosteroid[b]<br>Hydroxychloroquine[c]<br>Azithromycin |
| 4.0 | 14-Apr-2020 | Main (part A)<br><br><br><br><br><br>Second[d] | No additional treatment<br>Lopinavir-ritonavir[a]<br>Low-dose corticosteroid[b]<br>Hydroxychloroquine[c]<br>Azithromycin<br><br>No additional treatment<br>Tocilizumab |
| 5.0 | 24-Apr-2020 | - | (no change – extension to children <18 years old) |
| 6.0 | 14-May-2020 | Main (part A)<br><br><br><br><br><br>Main (part B factorial)<br><br><br>Second[d] | No additional treatment<br>Lopinavir-ritonavir[a]<br>Low-dose corticosteroid[b]<br>Hydroxychloroquine[c]<br>Azithromycin<br><br>No additional treatment<br>Convalescent plasma<br><br>No additional treatment<br>Tocilizumab |
| 7.0 | 18-Jun-2020 | Main (part A)<br><br><br><br><br>Main (part B factorial)<br><br><br>Second[d] | No additional treatment<br>Lopinavir-ritonavir[a]<br>Low-dose corticosteroid[b]<br>Azithromycin<br><br>No additional treatment<br>Convalescent plasma<br><br>No additional treatment<br>Tocilizumab |

| Protocol version | Date | Randomisation | Treatment arms |
|---|---|---|---|
| 8.0 | 03-Jul-2020 | Main (part A)<br><br><br><br><br><br>Main (part B factorial)<br><br><br>Second[d] | No additional treatment<br>Low-dose corticosteroid[b]<br>Intravenous immunoglobulin[e]<br>High-dose corticosteroid[e]<br>Azithromycin<br><br>No additional treatment<br>Convalescent plasma<br><br>No additional treatment<br>Tocilizumab |
| 9.1 | 18-Sep-2020 | Main (part A)<br><br><br><br><br><br>Main (part B factorial)<br><br><br><br>Second[d] | No additional treatment<br>Low-dose corticosteroid[b]<br>Intravenous immunoglobulin[e]<br>High-dose corticosteroid[e]<br>Azithromycin<br><br>No additional treatment<br>Convalescent plasma<br>REGEN-COV2<br><br>No additional treatment<br>Tocilizumab |
| 10.1 | 01-Nov-2020 | Main (part A)<br><br><br><br><br><br>Main (part B factorial)<br><br><br><br>Main (part C factorial)<br><br><br>Second[d] | No additional treatment<br>Low-dose corticosteroid[b]<br>Intravenous immunoglobulin[e]<br>High-dose corticosteroid[e]<br>Azithromycin[f]<br><br>No additional treatment<br>Convalescent plasma<br>REGEN-COV2<br><br>No additional treatment<br>Aspirin<br><br>No additional treatment<br>Tocilizumab |

[a] enrolment ceased 29 June 2020 when the Data Monitoring Committee advised that the Chief Investigators review the unblinded data.
[b] enrolment of adults ceased 8 June 2020 as more than 2,000 patients had been recruited to the active arm
[c] enrolment ceased 5 June 2020 when the Data Monitoring Committee advised that the Chief Investigators review the unblinded data.
[d] for patients with (a) oxygen saturation <92% on air or requiring oxygen or children with significant systemic disease with persistent pyrexia; and (b) C-reactive protein ≥75 md/dL)
[e] for children only
[f] enrolment of adults ceased 27 November 2020 as more than 2,500 patients had been recruited to the active arm

### Statistical Analysis Plans

### Statistical Analysis Plan V1.0

RECOVERY

Randomised Evaluation of COVID-19 Therapy

### Statistical Analysis Plan

**Version 1.0**

**Date: 09 June 2020**

Protocol version: 6.0, 14 May 2020

IRAS no: 281712
REC ref: EE/20/0101
ISRCTN: 50189673
EudraCT: 2020-001113-21

Nuffield Department of
POPULATION HEALTH

UNIVERSITY OF OXFORD

#### Abbreviations

| | |
|---|---|
| ADaM | Analysis Data Model |
| AE | adverse event |
| CDISC | The Clinical Data Interchange Standards Consortium |
| CI | confidence interval |
| CoV | Coronavirus |
| COVID | coronavirus-induced disease |
| CPAP | Continuous Positive Airway Pressure |
| CRP | C-reactive protein |
| CTU | clinical trials unit |
| CTSU | Clinical Trials Service Unit |
| DMC | Data Monitoring Committee |
| ECMO | Extra Corporeal Membrane Oxygenation |
| eCRF | Electronic case report form |
| $FiO_2$ | fraction of inspired oxygen |
| ICD | International Classification of Diseases |
| IFN | interferon |
| ICNARC | Intensive Care National Audit and Research Centre |
| IQR | interquartile range |
| ITT | intention to treat |
| MedDRA | Medical Dictionary for Regulatory Activities |
| MERS | Middle East Respiratory Syndrome |
| NPEU | National Perinatal Epidemiology Unit |
| OPCS-4 | NHS Classification of Interventions and Procedures |
| $PaO_2$ | partial pressure of oxygen |
| RR | risk ratio |
| SAE | serious adverse event |
| SARS | severe acute respiratory syndrome |
| SARS-CoV-2 | virus causing COVID-19 |
| SSAR | Suspected serious adverse reaction |
| SUSAR | Suspected unexpected serious adverse reaction |
| SD | standard deviation |
| SC | Steering Committee |

#### List of authors and reviewers

##### *Authors*

Dr Louise Linsell, Lead Trial Statistician, Nuffield Department of Population Health (NDPH), University of Oxford

Jennifer Bell, Trial Statistician, NDPH, University of Oxford

##### *Reviewers*

Professor Jonathan Emberson, Data Monitoring Committee (DMC) Statistician, NDPH, University of Oxford (prior to unblinded interim analysis of trial outcomes)

Professor Richard Haynes, Steering Committee (SC) Member, NDPH, University of Oxford

Professor Peter Horby, Chief Investigator (CI), Epidemic Diseases Research Group, University of Oxford

Professor Thomas Jaki, Steering Committee Member, Department of Mathematics and Statistics, Lancaster University

Associate Professor Edmund Juszczak, SC Member, NDPH, University of Oxford

Professor Martin Landray, Deputy CI, NDPH, University of Oxford

Professor Alan Montgomery, SC Member, Nottingham Clinical Trials Unit, University of Nottingham

Dr Natalie Staplin, DMC Statistician, NDPH, University of Oxford (prior to unblinded interim analysis of trial outcomes)

#### Roles and responsibilities

##### *Trial Statisticians*

Dr Louise Linsell and Jennifer Bell

NDPH, University of Oxford

Role: To develop the statistical analysis plan and conduct the final comparative analysis. Blinded to trial allocation.

##### *Data Monitoring Committee (DMC) Statisticians*

Professor Jonathan Emberson and Dr Natalie Staplin

NDPH, University of Oxford

Role: To conduct regular interim analyses for the DMC. Contribution restricted up until unblinded to trial allocation.

##### *Trial IT systems & Programmers*

Andy King, David Murray, Richard Welsh

NDPH, University of Oxford

Role: To generate and prepare reports monitoring the randomisation schedule. To supply data snapshots for interim and final analysis. Responsibility for randomisation system, clinical databases and related activities.

Bob Goodenough

NDPH, University of Oxford

Role: Validation of IT systems

Dr Will Stevens

NDPH, University of Oxford

Role: To produce analysis-ready datasets according to CDISC standards.

### 1 INTRODUCTION

This document details the proposed presentation and analysis for the main paper(s) reporting results from the multicentre randomised controlled trial RECOVERY (ISRCTN50189673) to investigate multiple treatments on major outcomes in inpatients for COVID-19 (clinically suspected or laboratory confirmed).

The results reported in these papers will follow the strategy set out here, which adheres to the guidelines for the content of a statistical analysis plan.[1] Any subsequent analyses of a more exploratory nature will not be bound by this strategy, and will be detailed in a separate analysis plan.

Suggestions for subsequent analyses by oversight committees, journal editors or referees, will be considered carefully in line with the principles of this analysis plan.

Any deviations from the statistical analysis plan will be described and justified in the final report to the funder. The analysis will be carried out by an identified, appropriately qualified and experienced statisticians, who will ensure the integrity of the data during their processing e.g. by parallel programming.

This statistical analysis plan is based on the latest version of the protocol. A record of amendments to the protocol can be found in the RECOVERY trial directory: https://www.recoverytrial.net/for-site-staff/site-set-up-1.

### 2 BACKGROUND INFORMATION

#### 2.1 Rationale

In early 2020, as the protocol was being developed, there were no approved treatments for COVID-19. The aim of the trial is to provide reliable evidence on the efficacy of candidate therapies (including re-purposed and novel drugs) for suspected or confirmed COVID-19 infection on major outcomes in hospitalised adult patients receiving standard care.

#### 2.2 Objectives of the trial

##### 2.2.1 *Primary objective*

To provide reliable estimates of the effect of study treatments on all-cause mortality within 28 days of randomisation.

##### 2.2.2 *Secondary objectives*

To investigate the effect of study treatments on the duration of hospital stay, the need for (and duration of) ventilation, and the need for renal replacement therapy.

#### 2.3 Trial design

This is a multi-centre, multi-arm, adaptive, open-label, randomised controlled trial with three possible stages of randomisation. In the main randomisation patients are allocated to no additional treatment or one of 4 anti-viral or host-directed treatments. In addition, in a factorial design, eligible patients can also be allocated simultaneously to no additional treatment or convalescent plasma. Patients who deteriorate according to predefined criteria can be further randomised to no additional treatment or an immunomodulatory treatment. The trial is designed with streamlined processes in order to facilitate rapid large-scale recruitment with minimal data collection.

#### 2.4 Eligibility

##### 2.4.1 *Inclusion criteria*

Patients are eligible for the trial if all of the following are true:

- Hospitalised
- SARS-Cov-2 infection (clinically suspected or laboratory confirmed)
- No medical history that might, in the opinion of the attending clinician, put the patient at significant risk if they were to participate in the trial.

##### 2.4.2 *Exclusion criteria*

If one or more of the active drug treatments is not available at the hospital or is believed, by the attending clinician, to be contraindicated (or definitely indicated) for the specific patient, then this fact will be recorded via the web-based form prior to randomisation; random allocation will then be between the remaining (or indicated) arms.

#### 2.5 Treatments

All patients will receive standard management for the participating hospital. The main randomisation will be between the following treatment arms (although not all arms may be available at any one time). The doses listed are for adults; paediatric dosing is described in the protocol.

##### 2.5.1 *Main randomisation part A:*

- **No additional treatment**
- **Lopinavir 400mg-Ritonavir 100mg** by mouth (or nasogastric tube) every 12 hours for 10 days.
- **Corticosteroid** in the form of dexamethasone**,** administered as an oral liquid or intravenous preparation 6 mg once daily for 10 days. In pregnancy, prednisolone 40 mg administered by mouth (or intravenous hydrocortisone 80 mg twice daily) should be used instead.
- **Hydroxychloroquine** by mouth for 10 days (4 doses in first 24 hours and 1 dose every 12 hours for 9 days).
- **Azithromycin 500mg** by mouth (or nasogastric tube) or intravenously once daily for a total of 10 days.

##### 2.5.2 *Main randomisation part B:*

In a factorial design, eligible patients may be randomised to the arms below. The doses listed are for adults; paediatric dosing is described in the protocol.

- **No additional treatment**
- **Convalescent plasma** Single unit of ABO compatible convalescent plasma (275mls +/- 75 mls) intravenous per day on study days 1 (as soon as possible after randomisation) and 2 (with a minimum of 12 hour interval between 1st and 2nd units). ABO identical plasma is preferred if available. The second transfusion should not be given if patient has a suspected serious adverse reaction during or after the first transfusion.

##### 2.5.3 *Second randomisation for patients with progressive COVID-19*

Patients enrolled in the main RECOVERY trial and with clinical evidence of a hyper-inflammatory state may be considered for a second randomisation if they meet the following criteria:

- Randomised into the main RECOVERY trial no more than 21 days ago
- Clinical evidence of progressive COVID-19:
  - oxygen saturation <92% on room air or requiring oxygen (or in children, significant systemic disease with persistent pyrexia, with or without evidence of respiratory involvement); and
  - C-reactive protein (CRP) ≥75 mg/L
- No medical history that might, in the opinion of the attending clinician, put the patient at significant risk if they were to participate in this aspect of the RECOVERY trial

Eligible participants may be randomised between the following treatment arms:

- **No additional treatment**
- **Tocilizumab** by intravenous infusion with the dose determined by body weight.

#### 2.6 Definitions of primary and secondary outcomes

Outcomes will be assessed at 28 days and then 6 months after randomisation. Analysis of longer-term outcomes collected beyond this will be described in a separate Statistical Analysis Plan.

##### 2.6.1 *Primary outcome*

Mortality (all-cause)

##### 2.6.2 *Secondary clinical outcomes*

- Time to discharge from hospital
- Use of mechanical ventilation/Extra Corporal Membrane Oxygenation (ECMO) or death (among patients not on ventilation or ECMO at baseline)

##### 2.6.3 *Subsidiary clinical outcomes*

- Cause-specific mortality (COVID-19; cardiovascular; non-vascular; other)
- Use of renal dialysis or haemofiltration
- Serious cardiac arrhythmia (recorded in a subset)

- Use of ventilation (overall and by type)
- Duration of ventilation (overall and by type)

###### 2.6.4 *Detailed derivation of outcomes*

The detailed derivation of outcomes included in statistical analysis will be described separately in a data derivation document and included in the Study Data Reviewer's Guide.

##### 2.7 Hypothesis framework

For each of the primary, secondary and subsidiary outcomes, the null hypothesis will be that there is no true difference in effect between any of the treatment arms.

##### 2.8 Sample size

The larger the number randomised, the more accurate the results will be, but the numbers that can be randomised will depend critically on how large the epidemic becomes. If substantial numbers are hospitalised in the participating centres then it may be possible to randomise several thousand with moderate disease and a few thousand with severe disease. Some indicative sample sizes and projected recruitment will be estimated using emerging data for several different scenarios. Sample size and recruitment will be monitored by the Steering Committee (SC) throughout the trial.

##### 2.9 Randomisation

Eligible patients will be randomised using a 24/7 secure central web-based randomisation system, developed and hosted within NDPH, University of Oxford. Users of the system will have no insight into the next allocation, given that simple randomisation is being used. In the event that a patient is randomised inadvertently more than once during the same hospital admission, the first allocation will be used.

The implementation of the randomisation procedure will be monitored by the Senior Trials Programmer, and the SC notified if an error in the randomisation process is identified.

###### 2.9.1 *Main randomisation part A*

Simple randomisation will be used with a 2:1:1:1:1 allocation ratio to one of the following treatment arms (in addition to usual care), which is subject to change:

- No additional treatment
- Lopinavir-Ritonavir
- Corticosteroid
- Hydroxychloroquine
- Azithromycin

The randomisation programme will allocate patients in a ratio of 2:1 between the no additional treatment arm and each of the other arms that are not contra-indicated and available. Hence if all 4 active treatment arms are available, then the randomisation will be in the ratio 2:1:1:1:1. If one or more of the active drug treatments is not available at the hospital or is believed, by the attending clinician, to be contraindicated (or definitely indicated) for the specific patient, then this fact will be recorded via the web-based form prior to randomisation; random allocation will then be between the remaining arms (in a 2:1:1:1, 2:1:1 or 2:1 ratio).

##### 2.9.2 *Main randomisation part B*

In a factorial design, eligible patients will be randomised simultaneously using simple randomisation with allocation ratio 1:1 to one of the following arms:

- No additional treatment
- Convalescent plasma

##### 2.9.3 *Second randomisation for patients with progressive COVID-19*

Eligible participants may be randomised using simple randomisation with an allocation ratio 1:1 between the following arms:

- No additional treatment
- Tocilizumab

#### 2.10 Blinding

This is an open-label study. However, while the study is in progress, access to tabular results of study outcomes by treatment allocation will not be available to the research team, CIs, trial statisticians, clinical teams, or members of the SC (unless the DMC advises otherwise). The DMC and DMC statisticians will be unblinded.

#### 2.11 Data collection schedule

Baseline and outcome information will be collected on trial-specific electronic case report forms (eCRFs) and entered into a web-based IT system by a member of the hospital or research staff. Follow-up information will be collected on all study participants, irrespective of whether or not they complete the scheduled course of allocated study treatment. Study staff will seek follow-up information through various means, including routine healthcare systems and registries.

All randomised participants will be followed up until death or 6 months post-randomisation to the main trial (whichever is sooner). NHS Digital and equivalent organisations in the devolved nations will supply data fields relevant to trial baseline and outcome measures to NDPH, University of Oxford on a regular basis, for participants enrolled into the trial. This will be combined with the trial-specific data collected via the web-based IT system and adjudicated internally.

Longer term (up to 10 years) follow-up will be sought through linkage to electronic healthcare records and medical databases including those held by NHS Digital, Public Health England and equivalent bodies, and to relevant research databases (e.g. UK Biobank, Genomics England).

#### 2.12 Data monitoring

During the study all study data will be supplied in strict confidence to the independent DMC for independent assessment and evaluation. The DMC will request such analyses at a frequency relevant to the emerging data from this and other studies.

The DMC has been requested to determine if, in their view, the randomised comparisons in the study have provided evidence on mortality that is strong enough (with a range of uncertainty around the results that is narrow enough) to affect national and global treatment strategies. Hence, multiple reviews by the Data Monitoring Committee have no material impact on the final analysis. In such a circumstance, the DMC will inform the SC who will make the results available to the public and amend the trial arms accordingly.

#### 2.13 Trial reporting

The trial will be reported according to the principles of the CONSORT statements.[2, 3, 4] The exact composition of the trial publication(s) depends on the size of the epidemic, the availability of drugs, and the findings from the various pairwise comparative analyses (with the no additional treatment arm) in the main trial.

### 3 ANALYSIS POPULATIONS

#### 3.1 Population definitions

The intention to treat (ITT) population will be all participants randomised, irrespective of treatment received. This ITT population will be used for analysis of efficacy and safety data.

For interim analyses, baseline data will be reported for all participants with data available and outcome data will be reported for all participants who have died, been discharged from hospital, or reached day 28 after the first randomisation.

### 4 DESCRIPTIVE ANALYSES

#### 4.1 Participant throughput

The flow of participants through the trial will be summarised for each separate pairwise comparison using a CONSORT diagram, for the main and second randomisation separately. The flow diagram for the nested factorial design (main randomisation part B to convalescent plasma) will be stratified by the 5 arms included in the main randomisation part A. The flow diagrams will describe the numbers of participants randomly allocated, who received allocation, withdrew consent, and included in the ITT analysis population. The flow diagrams for arms in the main randomisation will also report the number of participants who underwent the second randomisation.

#### 4.2 Baseline comparability of randomised groups

The following characteristics will be described separately for patients randomised to each main comparison (for each separate pairwise comparison of active treatment with the no additional treatment arm), and separately for the first and second randomisation.

###### 4.2.1 Main randomisation (part A and B)

- Age at randomisation
- Sex
- Ethnicity
- Time since COVID-19 symptoms onset
- Time since hospitalisation
- Current respiratory support requirement
- Currently requiring renal dialysis or haemofiltration
- Comorbidities (diabetes, heart disease, chronic lung disease, tuberculosis, human immunodeficiency virus, severe liver disease, severe kidney impairment)
- If female, known to be pregnant

###### 4.2.2 Second randomisation

In addition to the above:

- Type of ventilation support currently required (none, CPAP alone, non-invasive ventilation, high-flow nasal oxygen, mechanical ventilation, ECMO)
- Latest oxygen saturation measurement (%)
- Latest CRP measurement (mg/L)
- Latest ferritin measurement (ng/mL)
- Latest creatinine measurement (µmol/L)
- Allocation in first randomisation
- Interval between first and second randomisation

The number and percentage will be presented for binary and categorical variables. The mean and standard deviation or the median and the interquartile range will be presented for continuous variables, or the range if appropriate. There will be no tests of statistical significance performed nor confidence intervals calculated for differences between randomised groups on any baseline variable.

##### 4.3 Completeness of follow-up

All reasonable efforts will be taken to minimise loss to follow-up, which is expected to be minimal as data collection for primary and secondary outcomes using trial-specific eCRFs is combined with linkage to routine clinical data on study outcomes from NHS Digital, ICNARC, and similar organisations in the devolved nations.

The number and percentage of participants with follow-up information at day 28 and at 6 months after the main randomisation will be reported. Data will be shown for each of the following: all-cause mortality, hospital discharge status, ventilation status, and will be shown for each randomised group for the main and second randomisation separately.

##### 4.4 Adherence to treatment

The number and proportion of patients who did not receive the treatment they were allocated to will be reported. If any other trial treatment options were known to be received, instead of or in addition to, the allocated treatment during the 28 day follow-up period after the first randomisation, these will be collected and reported. Details on the number of days (or doses) of treatment received will be reported for all trial treatments received where available.

### 5 COMPARATIVE ANALYSES

For all outcomes, the primary analysis will be performed on the intention to treat (ITT) population at 28 days after the main randomisation. An ITT analysis of all outcomes at 6 months post-randomisation will also be conducted.

Pairwise comparisons will be made between each treatment arm and the no additional treatment arm (reference group) in that particular randomisation (main randomisation part A, main randomisation part B, and second randomisation). Since not all treatments may be available or suitable for all patients, those in the no additional treatment arm will only be included in a given comparison if, at the point of their randomisation, they *could* alternatively have been randomised to the active treatment of interest (i.e. the active treatment was available at the time and it was not contra-indicated). The same applies to treatment arms added at a later stage; they will only be compared to those patients recruited concurrently.

#### 5.1 Main randomisation part A

##### 5.1.1 *Primary outcome*

Mortality (all-cause) will be summarised with counts and percentages by randomised comparison group. A time-to-event analysis will be conducted using the log-rank test, with the p-value reported. Kaplan-Meier estimates for the time to event will also be plotted (with associated log-rank p-values). The log-rank 'observed minus expected' statistic (and its variance) will be used to estimate the average event rate ratio and confidence interval for each treatment group versus the no additional treatment group.[5] For the primary outcome, discharge alive before the relevant time period (28 days) will be assumed as absence of the event (unless there is additional data confirming otherwise).

##### 5.1.2 *Secondary outcomes*

###### *5.1.2.1 Time to discharge from hospital*

A time-to-event analysis will be used to compare each treatment group with the no additional treatment group using Kaplan-Meier and the log-rank test, as described above. Patients who die in hospital will be censored after 28 days. This gives an unbiased estimate of the recovery rate and comparable estimates to the competing risks approach in the absence of other censoring (which is expected to be very minimal).[6]

###### *5.1.2.2 Use of mechanical ventilation/ECMO or death (among those not on ventilation or ECMO at randomisation)*

Counts and percentages will be presented by randomised group and the risk ratio will be calculated for each pairwise comparison with the no additional treatment arm, with confidence intervals and p-values reported. The absolute risk difference will also be presented with confidence intervals. Patients who were already on ventilation at randomisation will be excluded from the denominator.

###### 5.1.3 *Subsidiary clinical outcomes*

###### *5.1.3.1 Cause-specific mortality*

Cause-specific mortality will be analysed in a similar manner to the primary outcome. Deaths from other causes will be censored at the date of death and a separate survival curve will be presented for each cause of death (COVID-19, other infection, cardiovascular and other).

###### *5.1.3.2 Use of renal dialysis or haemofiltration*

Counts and percentages will be presented by randomised group and the risk ratio will be calculated for each pairwise comparison with the no additional treatment arm, with confidence intervals and p-values reported. The absolute risk difference will also be presented with confidence intervals. Patients who were already on renal dialysis or haemofiltration at randomisation will be excluded from the denominator.

###### *5.1.3.3 Major cardiac arrhythmia*

Counts and percentages will be presented by randomised group and the risk ratio for any major cardiac arrhythmia will be calculated for each pairwise comparison with the no additional treatment arm, with confidence intervals and p-values reported. The absolute risk difference will also be presented with confidence intervals. Type of arrhythmia will also be described: (i) atrial flutter or fibrillation; (ii) supraventricular tachycardia; (iii) ventricular tachycardia; (iv) ventricular fibrillation; (v) atrioventricular block requiring intervention, with subtotals for (i)-(ii) and (iii)-(iv).

###### *5.1.3.4 Use of ventilation (overall and by type)*

Counts and percentages will be presented by randomised group for patients who received any assisted ventilation. Patients who were already on assisted ventilation at randomisation will be excluded from the denominator. The number of patients receiving the different types of ventilation will also be reported: (i) CPAP; (ii) other non-invasive; (iii) high-flow nasal oxygen; (iv) mechanical; (v) ECMO, with subtotals for (i)-(iii) (non-invasive) and (iv)-(v) (invasive).

###### *5.1.3.5 Duration of ventilation (overall and by type)*

The mean (SD) duration of ventilation will be calculated in days from the main randomisation for each randomised group in those who received ventilation, separately for survivors and non-survivors. This will be reported overall for any assisted ventilation and separately for mechanical ventilation or ECMO. The mean difference and confidence intervals will be presented for each pairwise comparison with the no additional treatment arm.

##### 5.2 Main randomisation part B

For the evaluation of treatment effect in the factorial design, the main effect of convalescent plasma across all arms in main randomisation part A combined, will be presented and tested, as described in 5.1. Data stratified by allocation in part A will also be reported to aid interpretation, but no tests for statistical interaction will be performed.

Additional safety data will be collected in a subset of patients randomised to part B. These will be tabulated separately by allocation (convalescent plasma versus no additional treatment): (i) sudden worsening in respiratory status; (ii) severe allergic reaction; (iii) temperature >39°C or ≥2°C rise since randomisation; (iv) sudden hypotension, clinical haemolysis and thrombotic event.

#### 5.3 Second randomisation

Evaluation of treatment effects in the main randomisation and the second randomisation will be conducted independently, as described in 5.1. In addition to the overall comparison for Tocilizumab vs no additional treatment, results will be stratified according to allocation in the main randomisation (part A and part B), however no interaction tests will be performed between the allocations in the two stages.

#### 5.4 Pre-specified subgroup analyses

Pre-specified subgroup analyses will be conducted for the main randomisation (part A and part B) and the second randomisation, for the following outcomes:

- Mortality (all-cause)
- Time to discharge from hospital
- Use of mechanical ventilation/ECMO or death

The analyses will be conducted using a test for heterogeneity (or test for trend for 3 or more ordered groups). Results will be presented on forest plots as event rate ratios (or risk ratios) with confidence intervals. The following subgroups will be examined:

- Risk group (three risk groups with approximately equal number of deaths based on factors recorded at randomisation)
- Requirement for respiratory support at randomisation (None; Oxygen only; Ventilation or ECMO)
- Time since illness onset (≤7 days; >7 days)
- Age (<70; 70-79; 80+ years)
- Sex (Male; Female)
- Ethnicity (White; Black, Asian or Minority Ethnic; Unknown)

Additional analyses will set the results for children (<18 years) and pregnant women in the context of the overall results.

#### 5.5 Significance levels and adjustment of p-values for multiplicity

Evaluation of the primary trial (main randomisation) and secondary randomisation will be conducted independently and no adjustment be made for these. Formal adjustment will not be made for multiple treatment comparisons, the testing of secondary and subsidiary outcomes, or subgroup analyses. 95% confidence intervals will be presented for estimates of between-group effects throughout.

#### 5.6 Statistical software employed

The statistical software SAS version 9.4, R Studio 3.6.2 and Stata/SE version 15 (or later) for Windows will be used for the interim and final analyses.

#### 5.7 Data standards and coding terminology

Datasets for analysis will be prepared using CDISC standards for SDTM and ADaM. Wherever possible, clinical outcomes (which may be obtained in a variety of standards, including ICD10 and OPCS-4) will be coded using MedDRA version 20.1.

### 6 SAFETY DATA

Suspected serious adverse reactions (SSARs) and suspected unexpected serious adverse reactions (SUSARs) will be listed by trial allocation.

### 7 ADDITIONAL EXPLORATORY ANALYSIS

Any post-hoc analysis requested by the oversight committees, a journal editor or referees will be labelled explicitly as such. Any further future analyses not specified in the analysis protocol will be exploratory in nature and will be documented in a separate statistical analysis plan.

### 8 DIFFERENCES FROM PROTOCOL V6.0

Use and duration of ventilation are described as secondary objectives in the protocol, and listed as subsidiary outcomes in the statistical analysis plan. The testing of multiple treatment arms will not formally be adjusted for, but given the number of comparisons, due allowance will be made in their interpretation. Formal methods of adjustment for multiplicity were not adopted because of treatment arms being added over time (including the factorial convalescent plasma comparison), unequal recruitment into each arm, and the ultimate number of treatments under evaluation not known in advance. While methods for these situations exist it was felt that the resulting change in level of significance was not appropriate.

### 9 REFERENCES

#### 9.1 Trial documents

Dummy tables and the data derivation document can be found in the RECOVERY trial directory and will be published with this SAP on the trial website.

#### 9.2 Other references

### 10 APPROVAL

| | | |
|---|---|---|
| Lead Trial Statistician | Name: Dr Louise Linsell | |
| | Signature: | Date: |
| Chief Investigator | Name: Professor Peter Horby | |
| | Signature: | Date: |
| Deputy Chief Investigator | Name: Professor Martin Landray | |
| | Signature: | Date: |
| Steering Committee Statistician | Name: Associate Professor Ed Juszczak | |
| | Signature: | Date: |
| Steering Committee Statistician | Name: Professor Alan Montgomery | |
| | Signature: | Date: |
| Steering Committee Statistician | Name: Professor Thomas Jaki | |
| | Signature: | Date: |

#### 11 DOCUMENT HISTORY

| Version | Date | Edited by | Comments/Justification | Timing in relation to unblinded interim monitoring | Timing in relation to unblinding of Trial Statisticians |
|---|---|---|---|---|---|
| 0.1 | 20/03/20 | LL/JB | First draft. | Prior | Prior |
| 0.2 | 01/04/20 | LL/JB | Comments and amendments from Martin Landray, Jonathan Emberson & Natalie Staplin. Also aligned with updated protocol and CRFs. | Prior | Prior |
| 0.3 | 01/04/20 | EJ/LL | Further edits and comments. | Prior | Prior |
| 0.4 | 07/04/20 | JB/EJ/LL | Following statistics group meeting on 02/04/20. | Prior | Prior |
| 0.5 | 22/04/20 | JB/LL/EJ | Following statistics group meeting on 09/04/20 and further protocol update. | After | Prior |
| 0.6 | 24/04/20 | LL | Following statistics group meeting on 23/04/20. | After | Prior |
| 0.7 | 10/05/20 | LL | Protocol update. | After | Prior |
| 0.8 | 15/05/20 | LL | Following statistics group meeting on 15/05/20. | After | Prior |
| 0.9 | 27/05/20 | LL | Further comments from SC members prior to interim analysis on 28/05/20. | After | Prior |
| 1.0 | 09/06/20 | LL | Revised following the stopping of the hydrochloroquine arm, and prior to the trial statisticians receiving unblinded data for this arm. | After | Prior |

#### Statistical Analysis Plan V2.1 (including summary of changes)

RECOVERY

Randomised Evaluation of COVID-19 Therapy

### Statistical Analysis Plan

**Version 2.1**

**Date: 02 December 2020**

Aligned with protocol version: 11.1, 21 November 2020

IRAS no: 281712
REC ref: EE/20/0101
ISRCTN: 50189673
EudraCT: 2020-001113-21

Nuffield Department of
POPULATION HEALTH

UNIVERSITY OF
OXFORD

#### Abbreviations

| | |
|---|---|
| ADaM | Analysis Data Model |
| AE | Adverse event |
| CDISC | The Clinical Data Interchange Standards Consortium |
| CI | Confidence interval |
| COVID | Coronavirus-induced disease |
| CPAP | Continuous Positive Airway Pressure |
| CRP | C-reactive protein |
| DMC | Data Monitoring Committee |
| ECMO | Extra Corporeal Membrane Oxygenation |
| eCRF | Electronic case report form |
| ICD | International Classification of Diseases |
| ICNARC | Intensive Care National Audit and Research Centre |
| ITT | Intention to treat |
| MedDRA | Medical Dictionary for Regulatory Activities |
| OPCS-4 | National Health Service OPCS Classification of Interventions and Procedures version 4 |
| SARS | Severe acute respiratory syndrome |
| SARS-CoV-2 | Severe acute respiratory syndrome coronavirus 2 |
| SSAR | Suspected serious adverse reaction |
| SUSAR | Suspected unexpected serious adverse reaction |
| TSC | Trial Steering Committee |

#### List of authors and reviewers (up to and including SAP version 1.1)

##### *Authors*

Dr Louise Linsell, Lead Trial Statistician, Nuffield Department of Population Health (NDPH), University of Oxford

Jennifer Bell, Trial Statistician, NDPH, University of Oxford

##### *Reviewers*

Professor Jonathan Emberson, Data Monitoring Committee (DMC) Statistician, NDPH, University of Oxford (prior to unblinded interim analysis of trial outcomes)

Professor Richard Haynes, Clinical Coordinator, NDPH, University of Oxford

Professor Peter Horby, Chief Investigator (CI), Nuffield Department of Medicine, University of Oxford

Professor Thomas Jaki, TSC Member, Department of Mathematics and Statistics, Lancaster University

Associate Professor Edmund Juszczak, TSC Member, NDPH, University of Oxford (until 6 July 2020)

Professor Martin Landray, Deputy CI, NDPH, University of Oxford

Professor Alan Montgomery, TSC Member, Nottingham Clinical Trials Unit, University of Nottingham

Dr Natalie Staplin, DMC Statistician, NDPH, University of Oxford (prior to unblinded interim analysis of trial outcomes)

#### List of authors and reviewers (version 2.0 onwards)

##### *Authors*

Professor Edmund Juszczak, TSC Member (University of Nottingham from 6 July 2020)

Professor Alan Montgomery (University of Nottingham), TSC Member

Professor Thomas Jaki (University of Cambridge) co-investigator and TSC Member

##### *Reviewers*

Enti Spata, Jennifer Bell, Dr Louise Linsell, Trial Statisticians, NDPH, University of Oxford

Professor Richard Haynes, Clinical Coordinator, NDPH, University of Oxford

Professor Martin Landray, Deputy CI, NDPH, University of Oxford

Professor Peter Horby, CI, Nuffield Department of Medicine, University of Oxford

#### Roles and responsibilities

##### *Trial Statisticians*

*Until 30th September 2020:* Dr Louise Linsell and Jennifer Bell (NDPH, University of Oxford)

Role: To develop the statistical analysis plan (blinded to trial allocation) and conduct the final comparative analyses for Lopinavir-Ritonavir¸ Corticosteroid (dexamethasone) and Hydroxychloroquine (main randomisation part A).

*From 1st October 2020:* Enti Spata (NDPH, University of Oxford)

Role: To develop the statistical analysis plan (blinded to trial allocation) and conduct the final comparative analyses for all other treatment arms.

##### *Data Monitoring Committee (DMC) Statisticians*

Professor Jonathan Emberson and Dr Natalie Staplin (NDPH, University of Oxford)

Role: To conduct regular interim analyses for the DMC. Contribution restricted up until unblinded to trial allocation.

##### *Statisticians on the Trial Steering Committee (TSC)*

Professor Edmund Juszczak (University of Nottingham), Professor Alan Montgomery (University of Nottingham), and Professor Thomas Jaki (University of Cambridge)

Role: Major organisational and policy decisions, and scientific advice; blinded to treatment allocation.

##### *Trial IT systems & Programmers*

Andy King, David Murray, Richard Welsh (NDPH, University of Oxford)

Role: To generate and prepare reports monitoring the randomisation schedule. To supply data snapshots for interim and final analysis. Responsibility for randomisation system, clinical databases and related activities.

Bob Goodenough (NDPH, University of Oxford)

Role: Validation of IT systems

Dr Will Stevens, Karl Wallendszuz (NDPH, University of Oxford)

Role: To produce analysis-ready datasets according to CDISC standards.

### 1 INTRODUCTION

This document details the proposed presentation and analysis for the main paper(s) reporting results from the multicentre randomised controlled trial RECOVERY (ISRCTN50189673) to investigate multiple treatments on major outcomes in inpatients for COVID-19 (clinically suspected or laboratory confirmed).

The results reported in these papers will follow the strategy set out here, which adheres to the guidelines for the content of a statistical analysis plan (SAP).[1] Any subsequent analyses of a more exploratory nature will not be bound by this strategy.

Suggestions for subsequent analyses by oversight committees, journal editors or referees, will be considered carefully in line with the principles of this analysis plan.

Any deviations from the statistical analysis plan will be described and justified in the final report. The analysis will be carried out by identified, appropriately qualified and experienced statisticians, who will ensure the integrity of the data during their processing.

This SAP is based on multiple versions of the protocol. All regulatory documents can be found in the RECOVERY trial directory: https://www.recoverytrial.net/for-site-staff/site-set-up-1/regulatory-documents.

SAP versions 1.0 & 1.1 applied to the first three principal comparisons (hydroxychloroquine, dexamethasone, and lopinavir-ritonavir versus no additional treatment respectively), for which data matured in the first UK wave of the pandemic. However, due to its later introduction, enrolment of patients in the azithromycin arm was much slower. Over time, factorial randomisations and a second randomisation have been added, introducing new treatment arms including convalescent plasma, tocilizumab, synthetic neutralizing antibodies, and aspirin. These changes, combined with the fact that use of corticosteroids (one of the original treatment arms) is now the usual standard of care for many patients, makes this a sensible juncture to re-evaluate the SAP and produce version 2.0.

### 2 BACKGROUND INFORMATION

#### 2.1 Rationale

In early 2020, as the protocol was being developed, there were no approved treatments for COVID-19. The aim of the trial is to provide reliable evidence on the efficacy of candidate therapies (including re-purposed and novel drugs) for suspected or confirmed COVID-19 infection on major outcomes in hospitalised adult patients receiving standard care.

#### 2.2 Objectives of the trial

##### 2.2.1 *Primary objective*

To provide reliable estimates of the effect of study treatments on all-cause mortality within 28 days of the relevant randomisation.

##### 2.2.2 *Secondary objectives*

To investigate the effect of study treatments on the duration of hospital stay and on the combined endpoint of use of invasive mechanical ventilation (including Extra Corporal Membrane Oxygenation [ECMO]) or death.

#### 2.3 Trial design

This is a multi-centre, multi-arm, adaptive, open label, randomised controlled trial with three possible stages of randomisation, as described below. The trial is designed with streamlined processes in order to facilitate rapid large-scale recruitment with minimal data collection.

#### 2.4 Eligibility

##### 2.4.1 *Inclusion criteria*

Patients are eligible for the trial if all of the following are true:

- Hospitalised
- SARS-Cov-2 infection (clinically suspected or laboratory confirmed)
- No medical history that might, in the opinion of the attending clinician, put the patient at significant risk if they were to participate in the trial.

##### 2.4.2 *Exclusion criteria*

If one or more of the active drug treatments is not available at the hospital or is believed, by the attending clinician, to be contraindicated (or definitely indicated) for the specific patient, then this fact will be recorded via the web-based form prior to randomisation; random allocation will then be between the remaining arms.

#### 2.5 Treatments

All patients will receive standard management for the participating hospital. The main randomisation will be between the following treatment arms (although not all arms may be available at any one time). The doses listed are for adults; paediatric dosing is described in the protocol.

##### 2.5.1 *Main randomisation part A:*

- **No additional treatment**
- **Lopinavir 400mg-Ritonavir 100mg** by mouth (or nasogastric tube) every 12 hours for 10 days. [Introduced in protocol version 1.0; **enrolment closed** 29 June 2020]

- **Corticosteroid** in the form of dexamethasone**,** administered as an oral liquid or intravenous preparation 6 mg once daily for 10 days. In pregnancy, prednisolone 40 mg administered by mouth (or intravenous hydrocortisone 80 mg twice daily) should be used instead. [Introduced in protocol version 1.0; **enrolment closed to adults** 8 June 2020]
- **Hydroxychloroquine** by mouth for 10 days (4 doses in first 24 hours and 1 dose every 12 hours for 9 days). [Introduced in protocol version 2.0; **enrolment closed** 5 June 2020]
- **Azithromycin 500mg** by mouth (or nasogastric tube) or intravenously once daily for a total of 10 days. [Introduced in protocol version 3.0; enrolment closed 27 November 2020]
- **Colchicine** by mouth for 10 days (1.5 mg in first 12 hours then 0.5 mg twice daily)

##### 2.5.2 *Main randomisation part B:*

In a factorial design, eligible patients may be randomised to the arms below. The doses listed are for adults; paediatric dosing is described in the protocol.

- **No additional treatment**
- **Convalescent plasma** Single unit of ABO compatible convalescent plasma (275mls ± 75 mls) intravenous per day on study days 1 (as soon as possible after randomisation) and 2 (with a minimum of 12-hour interval between 1st and 2nd units). ABO identical plasma is preferred if available. The second transfusion should not be given if patient has a suspected serious adverse reaction during or after the first transfusion. [Introduced in protocol version 6.0; enrolment ongoing]
- **Synthetic neutralising antibodies** (REGN-COV2; adults and children aged ≥12 years only - children who weigh <40kg will also not be eligible for this treatment). A single dose of REGN10933 + REGN10987 8 g (4 g of each monoclonal antibody) in 250ml 0.9% saline infused intravenously over 60 minutes ± 15 minutes as soon as possible after randomisation. [Introduced in protocol version 9.1; enrolment ongoing]

##### 2.5.3 *Main randomisation part C:*

In a factorial design, eligible patients may be randomised to the arms below. The dose listed is for adults; children are excluded from this comparison.

- **No additional treatment**
- **Aspirin** 150 mg by mouth (or nasogastric tube) or per rectum once daily until discharge. [Introduced in protocol version 10.1; enrolment ongoing]

##### 2.5.4 *Second randomisation for patients with progressive COVID-19*

Patients enrolled in the main RECOVERY trial and with clinical evidence of a hyper-inflammatory state may be considered for a second randomisation if they meet the following criteria:

- Randomised into the main RECOVERY trial no more than 21 days ago
- Clinical evidence of progressive COVID-19:
    - oxygen saturation <92% on room air or requiring oxygen (or in children, significant systemic disease with persistent pyrexia, with or without evidence of respiratory involvement); and
    - C-reactive protein (CRP) ≥75 mg/L
- No medical history that might, in the opinion of the attending clinician, put the patient at significant risk if they were to participate in this aspect of the RECOVERY trial

Eligible participants may be randomised between the following treatment arms:

- **No additional treatment**
- **Tocilizumab** by intravenous infusion with the dose determined by body weight. [Introduced in protocol version 4.0; enrolment ongoing]

#### 2.6 Definitions of primary and secondary outcomes

Outcomes will be assessed at 28 days and then 6 months after the relevant randomisation. Analysis of longer-term outcomes collected beyond this will be described in a separate Statistical Analysis Plan.

##### 2.6.1 *Primary outcome*

Mortality (all-cause)

##### 2.6.2 *Secondary clinical outcomes*

- Time to discharge from hospital
- Use of invasive mechanical ventilation (including Extra Corporal Membrane Oxygenation [ECMO]) or death (among patients not on invasive mechanical ventilation or ECMO at time of randomisation)

##### 2.6.3 *Subsidiary clinical outcomes*

- Use of ventilation (overall and by type) among patients not on ventilation (of any type) at time of randomisation
- Duration of invasive mechanical ventilation among patients on invasive mechanical ventilation at time of randomisation (defined as time to successful cessation of invasive mechanical ventilation: see section 5.1.3.2)
- Use of renal dialysis or haemofiltration (among patients not on renal dialysis or haemofiltration at time of randomisation)
- Thrombotic events (overall and by type; introduced in Protocol version 10.1)

##### 2.6.4 *Safety outcomes*

- Cause-specific mortality (COVID-19, other infection, cardiac, stroke, other vascular, cancer, other medical, external, unknown cause)

- Major cardiac arrhythmia (recorded on follow-up forms completed from 12 May 2020 onwards)
- Major bleeding (overall and by type; introduced in Protocol version 10.1)
- Early safety of antibody-based therapy (sudden worsening in respiratory status; severe allergic reaction; temperature >39°C or ≥2°C rise since randomisation; sudden hypotension; clinical haemolysis; and thrombotic events within the first 72 hours; Main randomization phase B only)

###### 2.6.5 *Detailed derivation of outcomes*

The detailed derivation of outcomes included in statistical analysis will be described separately in a data derivation document and included in the Study Data Reviewer's Guide.

##### 2.7 Hypothesis framework

For each of the primary, secondary and subsidiary outcomes, the null hypothesis will be that there is no true difference in effect between any of the treatment arms.

##### 2.8 Sample size

The larger the number randomised, the more accurate the results will be, but the numbers that can be randomised will depend critically on how large the epidemic becomes. If substantial numbers are hospitalised in the participating centres then it may be possible to randomise several thousand with moderate disease and a few thousand with severe disease. Some indicative sample sizes and projected recruitment will be estimated using emerging data for several different scenarios. Sample size and recruitment will be monitored by the TSC throughout the trial.

##### 2.9 Randomisation

Eligible patients will be randomised using a 24/7 secure central web-based randomisation system, developed and hosted within NDPH, University of Oxford. Users of the system will have no insight into the next allocation, given that simple randomisation is being used. If a patient is randomised inadvertently more than once during the same hospital admission, the first allocation will be used.

The implementation of the randomisation procedure will be monitored by the Senior Trials Programmer, and the TSC notified if an error in the randomisation process is identified.

###### 2.9.1 *Main randomisation part A*

Simple randomisation will be used to allocate participants to one of the following treatment arms (in addition to usual care), which is subject to change:

- No additional treatment
- Lopinavir-Ritonavir [Introduced in protocol version 1.0; **enrolment closed** 29 June 2020]
- Corticosteroid [Introduced in protocol version 1.0; **enrolment closed to adults** 8 June 2020]
- Hydroxychloroquine [Introduced in protocol version 2.0; **enrolment closed** 5 June 2020]

- Azithromycin [Introduced in protocol version 3.0; **enrolment closed 27 November 2020**]
- Colchicine [Introduced in protocol version 11.1]

The randomisation programme will allocate patients in a ratio of 2:1 between the no additional treatment arm and each of the other arms that are not contra-indicated and are available. Hence if all 4 active treatment arms are available, then the randomisation will be in the ratio 2:1:1:1:1. If one or more of the active drug treatments is not available at the hospital or is believed, by the attending clinician, to be contraindicated (or definitely indicated) for the specific patient, then this fact will be recorded via the web-based form prior to randomisation; random allocation will then be between the remaining arms (in a 2:1:1:1, 2:1:1 or 2:1 ratio).

##### 2.9.2 *Main randomisation part B*

In a factorial design, eligible patients will be randomised simultaneously using simple randomisation with allocation ratio 1:1:1 to one of the following arms, which is subject to change:

- No additional treatment
- Convalescent plasma [Introduced in protocol version 6.0; enrolment ongoing]
- Synthetic neutralising antibodies [Introduced in protocol version 9.1; enrolment ongoing]

If the active treatment is not available at the hospital, the patient does not consent to receive convalescent plasma, or is believed, by the attending clinician, to be contraindicated for the specific patient, then this fact will be recorded via the web-based form and the patient will be excluded from the relevant arm in Randomisation part B.

##### 2.9.3 *Main randomisation part C*

In a factorial design, eligible patients will be randomised simultaneously using simple randomisation with allocation ratio 1:1 to one of the following arms, which is subject to change:

- No additional treatment
- Aspirin [Introduced in protocol version 10.1; enrolment ongoing]

Note: From protocol version 7.0 onwards, randomisation is permitted in part B of main randomisation without randomisation in part A. From protocol version 10.1 onwards, randomisation is permitted in any combination of parts A, B, and C.

##### 2.9.4 *Second randomisation for patients with progressive COVID-19*

Eligible participants will be randomised using simple randomisation with an allocation ratio 1:1 between the following arms, which is subject to change:

- No additional treatment
- Tocilizumab [Introduced in protocol version 4.0; enrolment ongoing]

#### 2.10 Blinding

This is an open-label study. However, while the study is in progress, access to tabular results of study outcomes by treatment allocation will not be available to the research team, CIs, trial statisticians, clinical teams, or members of the TSC (unless the DMC advises otherwise). The DMC and DMC statisticians will be unblinded.

#### 2.11 Data collection schedule

Baseline and outcome information will be collected on trial-specific electronic case report forms (eCRFs) and entered into a web-based IT system by a member of the hospital or research staff. Follow-up information will be collected on all study participants, irrespective of whether they complete the scheduled course of allocated study treatment. Study staff will seek follow-up information through various means, including routine healthcare systems and registries.

All randomised participants will be followed up until death or 6 months post-randomisation to the main trial (whichever is sooner). NHS Digital and equivalent organisations in the devolved nations will supply data fields relevant to trial baseline and outcome measures to NDPH, University of Oxford on a regular basis, for participants enrolled into the trial. This will be combined with the trial-specific data collected via the web-based IT system and adjudicated internally.

Longer term (up to 10 years) follow-up will be sought through linkage to electronic healthcare records and medical databases including those held by NHS Digital, Public Health England and equivalent bodies, and to relevant research databases (e.g. UK Biobank, Genomics England).

#### 2.12 Data monitoring

During the study all study data will be supplied in strict confidence to the independent DMC for independent assessment and evaluation. The DMC will request such analyses at a frequency relevant to the emerging data from this and other studies.

The DMC has been requested to determine if, in their view, the randomised comparisons in the study have provided evidence on mortality that is strong enough (with a range of uncertainty around the results that is narrow enough) to affect national and global treatment strategies. Hence, multiple reviews by the Data Monitoring Committee have no material impact on the final analysis. In such a circumstance, the DMC will inform the TSC who will make the results available to the public and amend the trial arms accordingly.

#### 2.13 Trial reporting

The trial will be reported according to the principles of the CONSORT statements.[2, 3, 4] The exact composition of the trial publication(s) depends on the size of the epidemic, the availability of drugs, and the findings from the various pairwise comparative analyses (with the no additional treatment arm) in the main trial.

### 3 ANALYSIS POPULATIONS

#### 3.1 Population definitions

The intention to treat (ITT) population will be all participants randomised, irrespective of treatment received. This ITT population will be used for analysis of efficacy and safety data. For interim analyses, baseline data will be reported for all participants with data available and outcome data will be reported for all participants who have died, been discharged from hospital, or reached day 28 after the first randomisation.

### 4 DESCRIPTIVE ANALYSES

#### 4.1 Participant throughput

The flow of participants through the trial will be summarised for each separate pairwise comparison using a CONSORT diagram. The flow diagram will show the contribution of participants from each of the paths (from each of the parts of the main randomisation and from the second randomisation), where applicable. The flow diagrams will describe the numbers of participants randomly allocated, who received allocation, withdrew consent, and included in the ITT analysis population. The flow diagrams for arms in the main randomisation will also report the number of participants who underwent the second randomisation.

#### 4.2 Baseline comparability of randomised groups

The following characteristics will be described separately for patients randomised to each main comparison (for each separate pairwise comparison of active treatment with the no additional treatment arm), and separately for the first and second randomisation.

##### 4.2.1 Main randomisation (parts A, B and C)

- Age at randomisation
- Sex
- Ethnicity
- Region (UK, South East Asia)
- Time since COVID-19 symptoms onset
- Time since hospitalisation
- Current respiratory support
- Comorbidities (diabetes, heart disease, chronic lung disease, tuberculosis, human immunodeficiency virus, severe liver disease, severe kidney impairment)
- SARS-Cov-2 test result
- If female, known to be pregnant
- Use of systemic corticosteroid (including those allocated to corticosteroid in part A)
- Use of other relevant treatments (e.g. remdesivir, antiplatelet treatment, anticoagulant treatment)
- For part B only, anti-SARS-CoV-2 antibody concentration
- For treatment comparisons introduced in protocol v9.1 onwards:
    - C-reactive protein
    - Estimated glomerular filtration rate (calculated using the CKD-EPI formula)
    - D-dimer

###### 4.2.2 Second randomisation

In addition to the above:

- Current respiratory support
- Latest oxygen saturation measurement
- Latest C-reactive protein
- Latest ferritin
- Latest estimated glomerular filtration rate (calculated using the CKD-EPI formula)
- Allocation in main randomisation parts A, B, and C
- Interval between first and second randomisation

The number and percentage will be presented for binary and categorical variables. The mean and standard deviation or the median and the interquartile range will be presented for continuous variables.

##### 4.3 Completeness of follow-up

All reasonable efforts will be taken to minimise loss to follow-up, which is expected to be minimal as data collection for primary and secondary outcomes using trial-specific eCRFs is combined with linkage to routine clinical data on study outcomes from NHS Digital, ICNARC, and similar organisations in the devolved nations.

The number and percentage of participants with follow-up information at day 28 and at 6 months after the relevant randomisation will be reported. Data will be shown for each of the following: all-cause mortality, hospital discharge status, ventilation status, and will be shown for each randomised group for the main and second randomisation separately.

##### 4.4 Adherence to treatment

The number and proportion of patients who did not receive the treatment they were allocated to will be reported. If any other trial treatment options were known to be received, instead of or in addition to, the allocated treatment during the 28-day follow-up period after the first randomisation, these will be collected and reported. Details on the number of days (or doses) of treatment received will be reported for all trial treatments received where available.

#### 5 COMPARATIVE ANALYSES

For all outcomes, the primary analysis will be performed on the intention to treat (ITT) population at 28 days after randomisation. An ITT analysis of all outcomes at 6 months post-randomisation will also be conducted.

Pairwise comparisons will be made between each treatment arm and the no additional treatment arm (reference group) in that particular randomisation (main randomisation part A, main randomisation part B, main randomisation part C, and second randomisation). Since not all treatments may be available or suitable for all patients, those in the no additional treatment arm will only be included in a given comparison if, at the point of their randomisation, they *could* alternatively have been randomised to the active treatment of interest (i.e. the active treatment was available at the time and it was not contra-indicated). The same applies to treatment arms added at a later stage; they will only be compared to those patients recruited concurrently.

#### 5.1 Main randomisation part A

##### 5.1.1 *Primary outcome*

Mortality (all-cause) will be summarised with counts and percentages by randomised comparison group. A time-to-event analysis will be conducted using the log-rank test, with the p-value reported. Kaplan-Meier estimates for the time to event will also be plotted (with associated log-rank p-values). The log-rank 'observed minus expected' statistic (and its variance) will be used to calculate the one-step estimate of the event rate ratio and confidence interval for each treatment group versus the no additional treatment group.[5] For the primary outcome, discharge alive before the relevant time period (28 days after randomisation) will be assumed as absence of the event (unless there is additional data confirming otherwise).

##### 5.1.2 *Secondary outcomes*

###### *5.1.2.1 Time to discharge alive from hospital*

A time-to-event analysis will be used to compare each treatment group with the no additional treatment group using the log-rank test. As described for the primary outcome, the rate ratio and its confidence interval will be estimated from the log-rank observed minus expected statistic and its variance, and Kaplan-Meier curves will be drawn. Patients who die in hospital will be censored after 28 days after randomisation. This gives an unbiased estimate of the recovery rate and comparable estimates to the competing risks approach in the absence of other censoring (which is expected to be very minimal).[6]

###### *5.1.2.2 Use of invasive mechanical ventilation (including ECMO) or death*

Counts and percentages will be presented by randomised group and the risk ratio will be calculated for each pairwise comparison with the no additional treatment arm, with confidence intervals and p-values reported. The absolute risk difference will also be presented with confidence intervals. Each component of this composite outcome will also be summarised. Patients who were already on invasive mechanical ventilation or ECMO at randomisation will be excluded from these analyses.

##### 5.1.3 *Subsidiary clinical outcomes*

###### *5.1.3.1 Use of ventilation (overall and by type)*

Counts and percentages will be presented by randomised group for patients who received any assisted ventilation, together with risk ratios and confidence intervals for each pairwise comparison with the no additional treatment arm. The number of patients receiving the two main types of ventilation will also be reported: non-invasive ventilation (including CPAP, other non-invasive ventilation or high-flow nasal oxygen), and invasive mechanical ventilation (including ECMO). Patients who were already receiving ventilation[a] at randomisation will be excluded from these analyses.

###### *5.1.3.2 Duration of invasive mechanical ventilation (time to successful cessation of invasive mechanical ventilation)*

Successful cessation of invasive mechanical ventilation will be defined as removal of invasive mechanical ventilation within (and survival to) 28 days after randomisation. A time-to-event analysis will be used to compare each treatment group with the no additional treatment group using the log-rank test, as described above. The rate ratio and its confidence interval will be estimated from the log-rank observed minus expected statistic and its variance, and Kaplan-Meier curves will be drawn. Patients who die within 28 days of randomisation will be censored *after* 28 days after randomisation. Patients who were not already on invasive mechanical ventilation or ECMO at randomisation will be excluded from these analyses.

###### *5.1.3.3 Use of renal dialysis or haemofiltration*

Counts and percentages will be presented by randomised group and the risk ratio will be calculated for each pairwise comparison with the no additional treatment arm, with confidence intervals and p-values reported. The absolute risk difference will also be presented with confidence intervals. Patients who were already on renal dialysis or haemofiltration at randomisation will be excluded from these analyses.

###### *5.1.3.4 Thrombotic event*

Counts and percentages will be presented by randomised group. The absolute risk differences will also be presented with confidence intervals. Type of thrombotic event will also be described: (i) acute pulmonary embolism; (ii) deep vein thrombosis; (iii) ischaemic stroke, (iv) myocardial infarction; (v) systemic arterial embolism; and (vi) all sites combined.

#### 5.2 Main randomisation part B

In the factorial design, the main effects of treatments evaluated in part B will be presented and tested across all arms in main randomisation parts A and C combined, as described in 5.1. (Assessments of whether the effects of treatments in part B vary depending on other randomised treatments are described in section 5.7).

#### 5.3 Main randomisation part C

In the factorial design, the main effects of treatments evaluated in part C will be presented and tested across all arms in main randomisation parts A and B combined, as described in 5.1. (Assessments of whether the effects of treatments in part C vary depending on other randomised treatments are described in section 5.7).

#### 5.4 Second randomisation

[a] For comparisons introduced to the main randomisation prior to protocol version 9.1, patients who were already receiving oxygen at randomisation will also be excluded from these analyses (since it is not possible to distinguish those who were already receiving non-invasive ventilation).

Evaluation of treatment effects in the main randomisation and the second randomisation will be conducted independently, as described in 5.1.

#### 5.5 Pre-specified subgroup analyses

Pre-specified subgroup analyses will be conducted for the main randomisation (parts A, B and C) and the second randomisation, for the following outcomes:

- Mortality (all-cause)
- Time to discharge from hospital
- Use of invasive mechanical ventilation (including ECMO) or death

Tests for heterogeneity (or tests for trend for 3 or more ordered groups) will be conducted to assess whether there is any good evidence that the effects in particular subgroups differ materially from the overall effect seen in all patients combined. Results will be presented on forest plots as event rate ratios, or risk ratios, with confidence intervals. The following subgroups will be examined based on information at randomisation:

- Age (<70; 70-79; 80+ years)
- Sex (Male; Female)
- Ethnicity (White; Black, Asian or Minority Ethnic)
- Region (UK, South East Asia)
- Time since illness onset (≤7 days; >7 days)
- Requirement for respiratory support
  - For main randomisation: None; Oxygen only (with or without non-invasive ventilation); Invasive mechanical ventilation (including ECMO)
  - For second randomisation: No ventilator support (including no or low-flow oxygen); Non-invasive ventilation (including CPAP, other non-invasive ventilation, or high-flow nasal oxygen), Invasive mechanical ventilation (including ECMO)
- Use of systemic corticosteroid (including dexamethasone)
- For part B only: Recipient anti-SARS-CoV-2 antibody concentration at randomisation (<8 $x10^6$ units; ≥8 $x10^6$ units[b])

#### 5.6 Sensitivity analyses

Sensitivity analyses of the primary and secondary outcomes will be conducted among those patients with a positive test for SARS-COV-2 (i.e. confirmed cases).

#### 5.7 Other exploratory analyses

In addition, exploratory analyses will be conducted to test for interactions between treatments allocated in each of the different randomisations, provided that doing so does not lead to premature unblinding of results for ongoing comparators.

[b] Measured by Oxford immunoassay. 8 $x10^6$ represents the threshold shown to distinguish evidence of prior infection from no evidence of prior infection with sensitivity and specificity >98%.[7] Exploratory analyses using other assays may determine appropriate cut-offs based on blinded inspection of associations between assay results and baseline characteristics and mortality.

Non-randomised exploratory analyses will be used to explore the likely influence of different levels of convalescent plasma antibody concentration on the efficacy of convalescent plasma.

Additional analyses will set the results for children (<18 years) and pregnant women in the context of the overall results.

#### 5.8 Adjustment for baseline characteristics

The main analyses described above will be unadjusted for baseline characteristics. However, if there are any important imbalances between the randomised groups in key baseline pre-specified subgroups (see section 5.4) or allocation in the orthogonal components of the main randomisation, where applicable, emphasis will be placed on analyses that are adjusted for the relevant baseline characteristic(s). This will be done using Cox regression for the estimation of adjusted hazard ratios and a log-binomial regression model for the estimation of adjusted risk ratios.

#### 5.9 Significance levels and adjustment of p-values for multiplicity

Evaluation of the primary trial (main randomisation) and secondary randomisation will be conducted independently, and no adjustment be made for these. Formal adjustment will not be made for multiple treatment comparisons, the testing of secondary and subsidiary outcomes, or subgroup analyses. However, due allowance for multiple testing will be made in the interpretation of the results: the larger the number of events on which a comparison is based and the more extreme the P-value after any allowance has been made for the nature of the particular comparison (i.e. primary or secondary; pre-specified or exploratory), the more reliable the comparison and, hence, the more definite any finding will be considered. 95% confidence intervals will be presented for the main comparisons.

#### 5.10 Statistical software employed

The statistical software SAS version 9.4 and R Studio 3.6.2 (or later) for Windows will be used for the interim and final analyses.

#### 5.11 Data standards and coding terminology

Datasets for analysis will be prepared using CDISC standards for SDTM and ADaM. Wherever possible, clinical outcomes (which may be obtained in a variety of standards, including ICD10 and OPCS-4) will be coded using MedDRA version 20.1.

### 6 SAFETY DATA

Suspected serious adverse reactions (SSARs) and suspected unexpected serious adverse reactions (SUSARs) will be listed by trial allocation.

For each of the following, counts and percentages will be presented by randomised group. Where possible, the absolute risk differences will also be presented with confidence intervals:

###### 6.1.1.1 Cause-specific mortality

Cause-specific mortality (COVID-19, other infection, cardiac, stroke, other vascular, cancer, other medical, external, unknown cause) will be analysed in a similar manner to the primary outcome.

###### 6.1.1.2 Major cardiac arrhythmia

Type of arrhythmia will also be described: (i) atrial flutter or fibrillation; (ii) supraventricular tachycardia; (iii) ventricular tachycardia; (iv) ventricular fibrillation; (v) atrioventricular block requiring intervention, with subtotals for (i)-(ii) and (iii)-(iv).

###### 6.1.1.3 Major bleeding

Type of bleeding will also be described: (i) intracranial bleeding; (ii) gastro-intestinal bleeding; (iii) other bleeding site, and (iv) all sites combined.

###### 6.1.1.4 Early safety of antibody-based therapy

Additional safety data will be collected in a subset of patients randomised to part B: (i) sudden worsening in respiratory status; (ii) severe allergic reaction; (iii) temperature >39°C or ≥2°C rise since randomisation; (iv) sudden hypotension; (v) clinical haemolysis; and (vi) thrombotic event.

### 7 ADDITIONAL POST-HOC EXPLORATORY ANALYSIS

Any post-hoc analysis requested by the oversight committees, a journal editor or referees will be labelled explicitly as such. Any further future analyses not specified in the analysis protocol will be exploratory in nature and will be documented in a separate statistical analysis plan.

### 8 DIFFERENCES FROM PROTOCOL

The testing of multiple treatment arms will not formally be adjusted for, but given the number of comparisons, due allowance will be made in their interpretation. Formal methods of adjustment for multiplicity were not adopted because of treatment arms being added over time (including the factorial convalescent plasma comparison), unequal recruitment into each arm, and the ultimate number of treatments under evaluation not known in advance.

This analysis plan will be updated prior to unblinding of the 6-month follow-up results. Additional analyses may be specified, e.g. to explore the impact of randomised treatment allocation on hospital re-admission for COVID-19.

### 9 REFERENCES

#### 9.1 Trial documents

Study protocol, case report forms, training materials, and statistical analysis plan are published on the trial website.

#### 9.2 Other references

### 10 APPROVAL

| | | |
|---|---|---|
| Trial Statistician | Name: Mr Enti Spata | |
| | Signature: | Date: |
| Chief Investigator | Name: Professor Peter Horby | |
| | Signature: | Date: |
| Deputy Chief Investigator | Name: Professor Martin Landray | |
| | Signature: | Date: |
| Steering Committee Statistician | Name: Professor Edmund Juszczak | |
| | Signature: | Date: |
| Steering Committee Statistician | Name: Professor Alan Montgomery | |
| | Signature: | Date: |
| Steering Committee Statistician | Name: Professor Thomas Jaki | |
| | Signature: | Date: |

#### 11 DOCUMENT HISTORY

| Version | Date | Edited by | Comments/Justification | Timing in relation to unblinded interim monitoring | Timing in relation to unblinding of Trial Statisticians |
|---|---|---|---|---|---|
| 0.1 | 20/03/20 | LL/JB | First draft. | Prior | Prior |
| 0.2 | 01/04/20 | LL/JB | Comments and amendments from Martin Landray, Jonathan Emberson & Natalie Staplin. Also aligned with updated protocol and CRFs. | Prior | Prior |
| 0.3 | 01/04/20 | EJ/LL | Further edits and comments. | Prior | Prior |
| 0.4 | 07/04/20 | JB/EJ/LL | Following statistics group meeting on 02/04/20. | Prior | Prior |
| 0.5 | 22/04/20 | JB/LL/EJ | Following statistics group meeting on 09/04/20 and further protocol update. | After | Prior |
| 0.6 | 24/04/20 | LL | Following statistics group meeting on 23/04/20. | After | Prior |
| 0.7 | 10/05/20 | LL | Protocol update. | After | Prior |
| 0.8 | 15/05/20 | LL | Following statistics group meeting on 15/05/20. | After | Prior |
| 0.9 | 27/05/20 | LL | Further comments from TSC members prior to interim analysis on 28/05/20. | After | Prior |
| 1.0 | 09/06/20 | LL | Revised following the stopping of the hydroxychloroquine arm, and prior to the trial statisticians receiving unblinded data for this arm. | After | Prior |
| 1.1 | 21/06/20 | LL/JB/RH | Additional clarification of ventilation denominators. Adjustment for any imbalances of subgroup characteristics between treatment arms at randomisation. Clarification of analysis of composite outcome. Removal of 'Unknown' ethnicity subgroup. Addition of section 5.5 Adjustment for baseline characteristics. | After | After unblinding of hydroxychloroquine and dexamethasone arms. |

| Version | Date | Edited by | Comments/Justification | Timing in relation to unblinded interim monitoring | Timing in relation to unblinding of Trial Statisticians |
|---|---|---|---|---|---|
| 2.0 | 04/11/20 | EJ/ES | Revised to reflect changes in protocol, including introduction of factorial randomisations and new arms, including convalescent plasma, tocilizumab, synthetic neutralizing antibodies (REGN-COV2, and aspirin. | Prior to interim analysis of aspirin arm<br><br>After interim analyses of all other arms | After unblinding of 28-day results for hydroxychloroquine, lopinavir-ritonavir, and dexamethasone arms.<br><br>Prior to unblinding of any other arms |
| 2.1 | 02/12/20 | ES | Addition of colchicine. Modification of definition of recipient antibody concentration subgroup. | Prior to interim analyses including antibody results or of colchicine arm. | After unblinding of 28-day results for hydroxychloroquine, lopinavir-ritonavir, and dexamethasone arms.<br><br>Prior to unblinding of any other arms |
